# Population Pharmacokinetic Analysis of Dexmedetomidine in Children using Real World Data from Electronic Health Records and Remnant Specimens

**DOI:** 10.1101/2021.05.03.21256553

**Authors:** Nathan T. James, Joseph H. Breeyear, Richard Caprioli, Todd Edwards, Brian Hachey, Prince J. Kannankeril, Jacob M. Keaton, Matthew D. Marshall, Sara L. Van Driest, Leena Choi

**Affiliations:** Departments of Biostatistics, Vanderbilt University Medical Center, Nashville, TN; Departments of Medicine, Vanderbilt University Medical Center, Nashville, TN; Departments of Pediatrics, Vanderbilt University Medical Center, Nashville, TN; Departments of Pharmaceutical Services, Vanderbilt University Medical Center, Nashville, TN; Departments of Center for Pediatric Precision Medicine, Vanderbilt University Medical Center, Nashville, TN; Departments of Center for Precision Health Research, National Human Genome Research Institute, National Institutes of Health, Bethesda, MD

**Keywords:** Population Pharmacokinetics, Dexmedetomidine, Pragmatic Research, Opportunistic Sampling, Real-World Data

## Abstract

**Aim:** Our objectives were to perform a population pharmacokinetic analysis of dexmedetomidine in children using remnant specimens and data from electronic health records (EHRs) and explore the impact of patient’s characteristics and pharmacogenetics on dexmedetomidine clearance.

**Methods:** Dexmedetomidine dosing and patient data were gathered from EHRs and combined with opportunistically sampled remnant specimens. Population pharmacokinetic models were developed using nonlinear mixed-effects modeling. The first stage developed a model without genotype variables; the second stage added pharmacogenetic effects.

**Results:** Our final study population included 354 post-cardiac surgery patients age 0 to 22 years (median 16 months). The final two-compartment model included allometric weight scaling and age maturation. Population parameter estimates and 95% confidence intervals were 27.3 L/hr (24.0 – 31.1 L/hr) for total clearance (CL), 161 L (139 – 187 L) for central compartment volume of distribution (V_1_), 26.0 L/hr (22.5 – 30.0 L/hr) for intercompartmental clearance (Q), and 7903 L (5617 – 11119 L) for peripheral compartment volume of distribution (V_2_). The estimate for postmenstrual age when 50% of adult clearance is achieved was 42.0 weeks (41.5 – 42.5 weeks) and the Hill coefficient estimate was 7.04 (6.99 – 7.08). Genotype was not statistically or clinically significant.

**Conclusion:** Our study demonstrates the use of real-world EHR data and remnant specimens to perform a population PK analysis and investigate covariate effects in a large pediatric population. Weight and age were important predictors of clearance. We did not find evidence for pharmacogenetic effects of *UGT1A4* or *UGT2B10* genotype or *CYP2A6* risk score.

**What is already known about this subject:**

∘ Previous dexmedetomidine pharmacokinetic (PK) studies in pediatric populations have limited sample size.
∘ Smaller studies present a challenge for identifying covariates that may impact individual PK profiles.

**What this study adds:**

∘ We performed a dexmedetomidine population PK study with a large pediatric cohort using data obtained from electronic health records and remnant plasma specimens to enable increased sample size.
∘ xsDifferences in PK due to *UGT1A4* or *UGT2B10* variants or *CYP2A6* risk score are not clinically impactful for this population.

## Introduction

Dexmedetomidine is an alpha2-agonist with anxiolytic, sedative, and analgesic properties with minimal effects on respiratory depression.^1,2^ It is routinely used as part of intraoperative anesthetic management during surgical repairs of congenital heart disease (CHD) and in the postoperative period in the intensive care unit (ICU)^3,4^ and is commonly dosed as a continuous intravenous (IV) infusion using a fixed weight-based rate (e.g., starting at 0.3 mcg/kg/h). This dosing regimen will be adequate for some, but necessarily results in inappropriately low or high dosing for others. The proper dose for these latter individuals is not achieved until the initial sedation effect is observed, recognized as inadequate or excessive by the clinical team, and the dose adjusted accordingly. These patients are at risk for dose-related dexmedetomidine side effects, including bradycardia and hypotension, or use of additional sedative agents, including opioid analgesics. Accurate prediction of an individual’s dexmedetomidine requirement (precision dosing) could help minimize titration time to achieve sedation and analgesia goals without toxicity.

Many population pharmacokinetic (PK) studies of dexmedetomidine in pediatric populations have been reported.^5–17^ For example, Potts et al.^17^ report on dexmedetomidine use in 95 pediatric ICU patients using data pooled from several previous studies, Su et al.^15^ studied 59 children on mechanical ventilation after open heart surgery, Pérez-Guillé et al.^12^ assessed 30 children undergoing ambulatory surgery, and Zuppa et al.^9^ examined dexmedetomidine PK among 119 children undergoing cardiac surgery. Most have a small number of individuals and frequent specimen collection. For pediatric ICU populations, the median sample size is 29.5 (range 18-119), and the median number of total drug levels collected is 236.5 (range 89 – 1967) with a median of 9 per subject (range 2 – 16).^5,7,9–12,14,15^ Some studies have addressed small sample size with methods that combine information from multiple populations including pooled pediatric analyses,^17^ creating “universal” models for children and adults,^6,8^ and Bayesian analyses with informative priors;^16^ however, even these models only include information from at most around 130 children.

Previously, studies have identified weight^5–12,14–17^ and age^5,6,8–10,14–17^, along with cardiac bypass^9,10,15^, as important factors to explain inter-individual variability. However, lower sample size may limit identification of additional covariates impacting inter-individual variability. For example, although it is known that dexmedetomidine is rapidly distributed and metabolized in the liver by two pathways – direct glucuronidation by uridine 5′-diphosphate-glucuronosyltransferase (UGT) 1A4 and 2B10 and cytochrome P450 (CYP) 2A6 mediated aliphatic hydroxylation^2,18^ – small studies of the impact of genetic variation or expression levels of these enzymes have failed to demonstrate pharmacogenetic associations.^19,20^ A study including 260 children demonstrated that carriers of the T allele of *CYP2A6* rs835309 had significantly lower concentrations of dexmedetomidine (TT + TG vs. GG, *p* – value = .025).^21^ A newly developed weighted genetic risk score to predict *CYP2A6* activity raises the possibility of better capturing the impact of variants across this gene for pharmacogenetic analysis.^22^ Study of a larger cohort may allow the identification of genetic biomarkers affecting dexmedetomidine PK, facilitating precision dosing based on genotype.

We combined data from electronic health records (EHRs) and remnant specimens collected during usual clinical care to perform a population PK analysis, similar to two previous pediatric fentanyl studies, and employing a system for constructing PK analysis datasets in R.^23–26^ The major goals of this study were to develop a dexmedetomidine population PK model for children with data obtained from EHRs and remnant specimens and quantify genetic effects that were selected *a priori* based on previous studies and known metabolic pathways.

## Methods

### Study Design

This study was approved by the Vanderbilt University Medical Center (VUMC) Institutional Review Board and has been previously described.^23^ In brief, pediatric patients undergoing surgery for CHD are offered enrollment in this observational study. Parents provide written consent for their child’s participation, and informed assent is obtained when appropriate. Drug selection and dosing are determined by the primary clinical team; over the course of study enrollment, clinical leadership provided recommended protocols to guide clinicians in drug and dose selection for analgesia and sedation (included in supplemental material); however, final regimens were always at the discretion of the treating clinicians. Remnant specimens from clinical testing are obtained for drug concentration measurements, which are not disclosed to the clinical teams. Specimens were not collected in connection with dose administration or to monitor PK characteristics such as trough concentration or C_max_. Enrollment with remnant specimen collection began in July 2012 and is ongoing. Data analyzed for this study were collected prior to October 2017. All study participants were admitted to the pediatric cardiac ICU after surgery. Enrolled participants were excluded from the analysis if their surgery was cancelled, if there was missing genotype data, if extracorporeal membrane oxygenation (ECMO) treatment was required, or if they did not survive to hospital discharge. For those with multiple surgeries, data from the one procedure with the highest number of measured serum drug concentrations were used, excluding all others. Drug concentrations were excluded if inadequate internal standard concentrations were detected and insufficient volume remained to repeat analysis, or if they were obtained before any documented dexmedetomidine dosing.

### Data Collection

Demographic data (including parent-reported race) and medical history were documented at the time of study enrollment. Surgical and clinical data were extracted from the EHR prospectively. Dexmedetomidine dosing, including scheduled boluses, as-needed intermittent boluses, and continuous infusions after the postoperative admission to the ICU were determined from the EHR and the Vanderbilt Enterprise Data Warehouse. The Enterprise Data Warehouse contains nurse administration, nurse flowsheets, and pharmacy dispense data, enabling the computation of administered drug amounts over specific time periods. Study data were collected and managed using REDCap electronic data capture tools, a secure, web-based application hosted at Vanderbilt University.^27^

### Drug Concentration Measurement

For the purposes of drug concentration analysis, all remnant plasma specimens ≥100 μL from blood obtained for clinical testing of electrolyte or basic metabolic panels in study subjects were obtained from the Vanderbilt Clinical Chemistry Laboratory and stored at −20ºC until processing for drug concentration analysis in the Vanderbilt Mass Spectrometry Research Core. Specimen processing and mass spectrometry analysis have been previously described in detail.^23^ Briefly, acetonitrile precipitation was followed by tandem mass spectrometry using a 16-drug assay. Dexmedetomidine assay accuracy is 89 – 112%, and the lower and upper limits of quantification (LLOQ and ULOQ) are 0.005 and 5 ng/mL.

### Genotyping and CYP2A6 Activity Score Prediction

Study participants provided a peripheral blood sample for genetic analysis. Genomic DNA was extracted through the Vanderbilt Technologies for Advanced Genomics (VANTAGE) Core laboratory and study participants were genotyped using either the Axiom™ Precision Medicine Research Array or the Precision Medicine Diversity Array according to manufacturer protocols at the Children’s Hospital of Philadelphia DNA core. As part of genotype data quality control, variants were removed if genotype call rate was <98%, if minor allele frequency was >20% different from 1000 Genomes phase 3 European reference populations, or for deviation from Hardy-Weinberg Equilibrium (*p* – value < 1×10^−10^, results shown in **Supplemental Table S3**). Individuals were removed if their genotype call rate was <98%, the genetically estimated sex differed from parental-reported sex, or for relatedness (2nd degree or closer). Genotype data were imputed to the 1000 Genomes phase 3 reference panel. For this study, we extracted data for specific variants in *UGT2B10* (rs2942857; rs112561475; rs61750900), *UGT1A4* (rs2011425; rs3892221; rs6755571) and *CYP2A6* (rs56113850; rs2316204; rs113288603; rs28399442; rs1801272; rs28399433) from the study database.

### Data Processing

Data was processed using the modularized “*EHRtoPKPD*” system for postmarketing population PK studies with real-world data from EHRs.^25^ This system has been implemented in the R software^28^ package EHR^26^ which includes interactive checks for data quality to reconcile missing, duplicate, and other erroneous concentration or dosing information (for details, see https://choileena.github.io/). Output from the EHR package was further cleaned by removing: (i) concentration measurements more than 150 hrs (approximately 50 times dexmedetomidine half-life) after the end of the final bolus or infusion dose, (ii) concentration measurements below the LLOQ if they are after the final bolus or infusion dose, except for the first such measurement, (iii) concentration measurements above the ULOQ, (iv) subjects whose only concentration measurements are below the LLOQ after applying criteria (i)-(iii), and (v) subjects with missing dose information indicated by increases in concentration without an accompanying dose and confirmed by manual chart review.

Serum creatinine concentration was a time-varying covariate typically measured concurrently with dexmedetomidine concentration. If serum creatinine was not available when dexmedetomidine concentration was measured, we selected the serum creatinine concentration measured closest to the dexmedetomidine concentration data within 7 days. For each subject, weight varied little within the timeframe of available concentration data, so most weight data were the same as the baseline demographic measurements. When additional weight measurements were available, usually during infusion, weight measurements obtained at the same time as the dosing event were used. Measures of albumin concentration were available within a 7-day window for only 48 subjects, precluding use of albumin concentration as a covariate.

### Population PK Analysis

We performed population PK analysis of dexmedetomidine using nonlinear mixed-effects models implemented by Monolix version 2020R1^29^ and estimated the parameters with the stochastic approximation expectation-maximization (SAEM) algorithm. Observed concentrations below the LLOQ were considered to be censored between 0 and 0.005 ng/mL and were handled in the modeling using the likelihood (M3) method for interval censoring.^30,31^ After the model parameters were estimated with SAEM, the objective function value (OFV) was calculated using Monte Carlo importance sampling with 10,000 samples from a Student-t proposal distribution and degrees of freedom chosen by testing a sequence of values (ν =1, 2, 5, 10, 15). Because the SAEM estimation method includes stochastic variability and can sometimes fail to converge in a setting with sparse sampling,^32^ we performed 5 runs with different random seeds for each model and selected the run with median OFV for model comparison.

For model selection we used a likelihood ratio test to compare differences in estimated OFV for nested models and corrected Bayesian Information Criteria (BICc) to compare non-nested models; relative standard errors, parameter estimate values, magnitude of random effects and change in CV% were also considered. In addition, we used several graphical methods for model evaluation including observed vs. population and individual predictions, individual weighted residuals vs. predicted concentration and time, correlations between samples from the conditional random effects distributions, samples from the conditional random effects distributions vs. covariates, and prediction corrected visual predictive checks.^33,34^

All covariates were chosen *a priori* based on previous research and biological plausibility, including *UGT1A4, UGT2B10*, and *CYP2A6* variants, age, sex, Society of Thoracic Surgery–European Association for Cardio-Thoracic Surgery (STAT) Congenital Heart Surgery Mortality score,^35^ cardiac bypass time, length of ICU stay, and serum creatinine. We focused on modeling the effects of covariates on total clearance, although the graphical checks were examined for possible relationships between covariates and other PK parameters.

Model building proceeded in two stages; we first considered all covariates except *UGT1A4, UGT2B10*, and *CYP2A6* to build an adequate model for dexmedetomidine PK and then examined the hypothesized association between the genotype variables and total clearance by adding these effects individually to the stage one model. For stage one we explored models with various structural, residual error, and inter-individual variance components and adjusted for non-genotype covariates. Following a strategy outlined by Bonate, we began with richly parameterized inter-individual variability and residual error models including all random effects, all correlations between random effects, and combined additive and proportional residual error, and then simplified this structure.^36^ We examined the structural model by comparing one- and two-compartment models without covariates. Following this we considered size and age maturation; these two covariates have been shown to be important factors in pediatric PK models with a large age range and in previous dexmedetomidine studies.^6,8,16,17,37–39^ For size, we employed an allometric weight model with fixed or estimated scaling parameters. For maturation, we considered an exponential age model, a sigmoid Hill maturation model, a body-weight dependent exponent model, and an age-dependent exponent model.^40^ Next, we investigated whether other non-genotype covariates improved the model with size and maturation factors and refined residual and inter-individual variance structure. Each covariate was considered as an exponentially linear or categorical term.

In the second stage we tested for the association between genotype and total clearance by including these effects in the model found in stage one. For *UGT1A4* and *UGT2B10*, dichotomous models (coding individuals as having a loss-of-function variant or not) and additive models (counting the number of variants) were considered. For *CYP2A6*, enzyme activity was predicted using a polygenic score and included as an exponential term.^22^ Details of all models explored along with specific mathematical relationships, estimated OFV, and BICc are shown in **Supplemental Tables S3 – S10**. Graphical checks for the model selection process are shown in **Supplemental Figures S3 – S27**.

## Results

### Study Population and Specimens

We collected 4,369 residual plasma specimens from 620 subjects. After removing 89 subjects with unknown sample collection time, 108 subjects with no dosing information within 7 days of the first concentration measurement, and 14 subjects due to in hospital mortality or ECMO, the output of the EHR package pipeline contained 411 subjects with 2,172 dexmedetomidine concentration measurements. The further cleaning steps described above removed 14 subjects and 43 more subjects without genotype information were also removed. The study cohort flow diagram of data processing is shown in **Figure 1**, and the final study population of 354 subjects with 1,400 specimens is described in **Table 1**. The median postnatal age was 16 months (interquartile range [IQR] 5 – 62), median postmenstrual age was 105 weeks (IQR 62 - 304) and median weight was 9.4 kg (IQR 6.0 – 18.2). The age and weight distributions are shown in **Supplemental Figures S1** and **S2** and **Supplemental Table S1** shows postnatal age categories.

**Table 1.** Study Cohort. Summary of demographic, genotype, clinical, dosing, and specimen sampling characteristics

|  | Entire cohort |
| --- | --- |
| n | 354 |
| Postnatal Age (months) | 15.7 (5.3 – 61.8)<br>[0.03, 270.9] |
| Postmenstrual Age (weeks) | 104.6 (61.7 – 303.7)<br>[39.1, 1200.0] |
| Median weight (kg) | 9.4 (6.0 – 18.2)<br>[2.0, 138.0] |
| Male sex | 183 (52%) |
| Race |  |
| White | 293 (83%) |
| Black | 40 (11%) |
| American Indian or Alaska Native | 2 (1%) |
| Asian | 6 (2%) |
| Other | 5 (1%) |
| Unknown | 8 (2%) |
| Median serum creatinine (mg/dL) | 0.49 (0.44 – 0.56)<br>[0.24, 1.12] |
| STAT score |  |
| 1 | 154 (43%) |
| 2 | 108 (31%) |
| 3 | 41 (12%) |
| 4 | 46 (13%) |
| 5 | 5 (1%) |
| *UGT1A4* variants |  |
| 0 | 262 (74%) |
| 1 | 87 (25%) |
| 2 | 5 (1%) |
| *UGT2B10* variants |  |
| 0 | 186 (53%) |
| 1 | 117 (33%) |
| 2 or 3 | 51 (14%) |
| *CYP2A6* score <sup>a</sup> | 2.04 (2.00 – 2.21)<br>[1.58, 2.43] |
| Cardiac bypass time (hr) | 1.66 (1.2 – 2.4)<br>[0, 7.1] |
| Length of ICU hospitalization (days) | 4 (2 – 6)<br>[1, 120] |
| Total dosing events <sup>b</sup> | 2386 |
| Total IV infusion doses | 2351 |
| Infusion duration (hr) | 2.0 (1.0 – 5.1) [0.02, 125.6] |
| Infusion rate (mcg/kg/hr) | 0.6 (0.5 – 1.0) [0.03, 2.0] |
| Total IV bolus doses | 35 |
| Bolus dose amount (mcg/kg) | 1.00 (0.96 – 1.01) [0.06, 4.21] |
| Dosing events per subject | 5 (3 – 8)<br>[1, 37] |
| Total dexmedetomidine concentration measurements | 1400 |
| Dexmedetomidine measurements below lower limit of quantification | 120 |
| Dexmedetomidine measurements per subject | 3 (2 – 5)<br>[1, 18] |
| First dexmedetomidine measurement time after dose start (hr) | 5.3 (3.7 – 11.3)<br>[0, 178.5] |
| Final dexmedetomidine measurement time after dose start (hr) | 67.7 (39.3 – 130.5)<br>[2.0, 659.8] |

Continuous variable summary statistics: median (interquartile range) [minimum, maximum]; Categorical variable summary statistics: number (%); <sup>a</sup>Among n=350 subjects with available score; <sup>b</sup> A dosing event is defined as a bolus administration or an infusion interval with constant administration rate for a specific duration.

**Figure 1.**
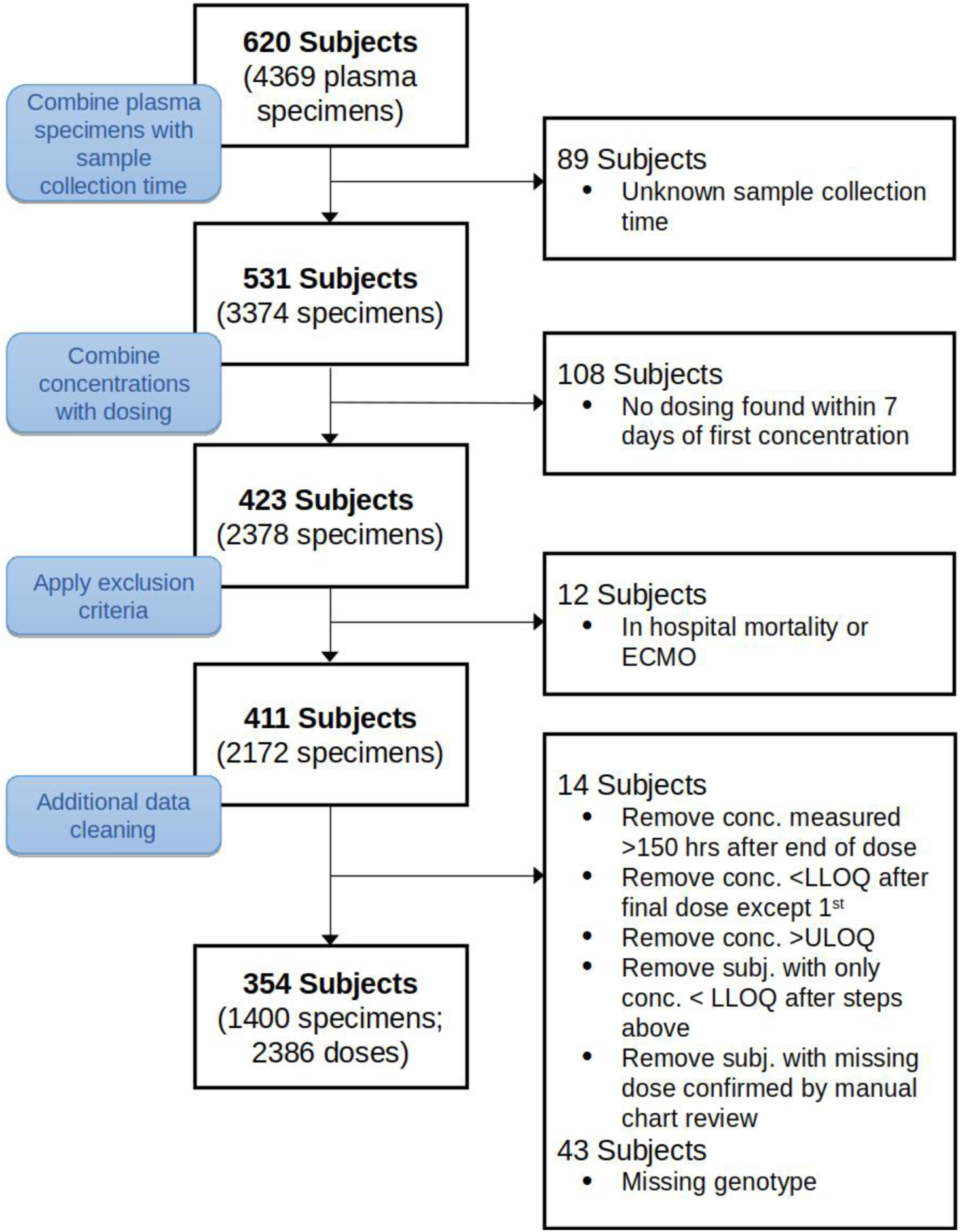
Study Cohort Flow Diagram in Data Processing with Exclusion Criteria.

There were 262 subjects (74%) with no variants of *UGT1A4*, 87 (25%) with 1 variant and 5 (1%) with 2 variants. For *UGT2B10*, 186 subjects (53%) had no variants, 117 (33%) had 1 variant and 51 (14%) had 2 or 3 variants. The *CYP2A6* predicted activity score was available for 350 of the 354 subjects (median 2.04, IQR 2.00 – 2.21). There were 2,386 dexmedetomidine dosing events (2,351 IV infusions and 35 bolus administrations). The median infusion rate was 0.6 mcg/kg/hr (IQR 0.5 – 1.0) and the median infusion duration was 2 hours (IQR 1 - 5); the median bolus dose was 1.0 mcg/kg (IQR 0.96 – 1.01). The top ten concomitant medications were acetaminophen (92.5%), cefazolin (92%), famotidine (89.1%), morphine (88.5%), furosemide (80.5%), fentanyl (77.9%), rocuronium (69.5%), oxycodone (59.8%), heparin (58.3%), and lorazepam (51.4%). **Supplemental Table S2** includes all concomitant medications administered to at least 5% of subjects. The number of dexmedetomidine concentration measurements per subject varied from a minimum of 1 to a maximum of 18 with a median of 3 specimens (IQR 2 – 5). The median time of first dexmedetomidine measurement after dose start was 5 hours (IQR 4 – 11) and the median time of final dexmedetomidine measurement after dose start was 68 hours (IQR 39 – 131).

### Population PK Model

In the first stage of modeling, a two-compartment model with additive and proportional residual error was chosen as the base model based on BICc and graphical checks. The main PK parameters are total clearance (CL, L/h), volume of distribution for the central compartment (V_1_, L), inter-compartmental clearance (Q, L/h) and volume of distribution for the peripheral compartment (V_2_, L). The results for the base and covariate models without genotype are presented in **Table 2A**. The coefficients of variation (CV) for CL, V_1_, Q, and V_2_ in the base model were 201%, 161%, 146%, and 672%, respectively.

**Table 2.** Estimates of Parameters for Population Pharmacokinetic Models.

| Base Model<br>(Obj = -1517.3; BICc = -1415.1) |  | Weight Only Model<br>(Obj = -1923.3; BICc = -1821.1) |  | Weight and Maturation with simplified variance structure<br>(Obj = -1915.1; BICc = -1835.0) |  |
| --- | --- | --- | --- | --- | --- |
| Parameters | Estimates (SE)<br>[95% CI]<sup>a</sup> | Parameters | Estimates (SE)<br>[95% CI] | Parameters | Estimates (SE)<br>[95% CI] |
| *CL* | | $CL = \theta_1(WT/70)^{0.75}$ | | $CL = \theta_1(WT/70)^{0.75} \left( \frac{1}{1 + \left( \frac{TM_{50}}{PMA} \right)^{Hill}} \right)$ | |
| | 6.27 (0.52)<br>[5.33, 7.37] | $\theta_1$ | 22.3 (1.85)<br>[19.0, 26.2] | $\theta_1$ | 27.3 (1.82)<br>[24.0, 31.1] |
| | | | | $TM_{50}$ | 41.9 (0.28)<br>[41.4, 42.5] |
|  |  |  |  | Hill | 7.04 (0.022)<br>[6.99, 7.08] |
| *V<sub>I</sub>* | | $V_1 = \theta_2(WT/70)$ | | $V_1 = \theta_2(WT/70)$ | |
| | 19.5 (1.37)<br>[17.0, 22.4] | $\theta_2$ | 123 (11.5)<br>[102, 148] | $\theta_2$ | 161 (12.1)<br>[139, 187] |
| *Q* | | $Q = \theta_3(WT/70)^{0.75}$ | | $Q = \theta_3(WT/70)^{0.75}$ | |
| | 5.26 (0.44)<br>[4.47, 6.20] | $\theta_3$ | 26.6 (2.23)<br>[22.6, 31.3] | $\theta_3$ | 26.0 (1.90)<br>[22.5, 30.0] |
| *V<sub>2</sub>* | | $V_2 = \theta_4(WT/70)$ | | $V_2 = \theta_4(WT/70)$ | |
| | 763 (126)<br>[555, 1048] | $\theta_4$ | 6674 (1499)<br>[4363, 10211] | $\theta_4$ | 7903 (1408)<br>[5617, 11119] |
| $\omega_{CL}$ (%CV) | 201 (12) [178, 226] | $\omega_{CL}$ (%CV) | 123 (13) [100, 150] | $\omega_{CL}$ (%CV) | 103 (8) [88, 120] |
| $\omega_{V1}$ (%CV) | 161 (9) [145, 179] | $\omega_{V1}$ (%CV) | 168 (22) [130, 217] | $\omega_{V1}$ (%CV) | 138 (13) [114, 166] |
| $\omega_Q$ (%CV) | 146 (8) [132, 162] | $\omega_Q$ (%CV) | 91 (11) [71, 115] | $\omega_Q$ (%CV) | 82 (9) [65, 102] |
| $\omega_{V2}$ (%CV) | 672 (81) [534, 855] | $\omega_{V2}$ (%CV) | 857 (256) [494, 1595] | $\omega_{V2}$ (%CV) | 624 (157) [391, 1048] |
| $\rho_{CL,V1}$ | 0.964 (0.003)<br>[0.959, 0.97] | $\rho_{CL,V1}$ | 0.849 (0.055)<br>[0.741, 0.956] | $\rho_{CL,V1}$ | 0.923 (0.027)<br>[0.871, 0.975] |
| $\rho_{CL,V2}$ | -0.032 (0.043)<br>[-0.116, 0.053] | $\rho_{CL,V2}$ | -0.174 (0.101)<br>[-0.372, 0.023] | $\rho_{CL,V2}$ | 0 (fixed) |
| $\rho_{CL,Q}$ | 0.043 (0.076)<br>[-0.106, 0.192] | $\rho_{CL,Q}$ | -0.509 (0.110)<br>[-0.726, -0.293] | $\rho_{CL,Q}$ | 0 (fixed) |
| $\rho_{V1,V2}$ | 0.195 (0.044)<br>[0.109, 0.28] | $\rho_{V1,V2}$ | 0.277 (0.107)<br>[0.068, 0.486] | $\rho_{V1,V2}$ | 0 (fixed) |
| $\rho_{V1,Q}$ | -0.076 (0.073)<br>[-0.22, 0.067] | $\rho_{V1,Q}$ | -0.410 (0.125)<br>[-0.656, -0.165] | $\rho_{V1,Q}$ | 0 (fixed) |
| $\rho_{V2,Q}$ | 0.089 (0.083)<br>[-0.074, 0.252] | $\rho_{V2,Q}$ | 0.21 (0.121)<br>[-0.028, 0.448] | $\rho_{V2,Q}$ | 0 (fixed) |
| $\sigma_{\text{add}}$ (ng/mL) | 2.22e-16 (5.65e-13)<br>[0, 1.11e-12] | $\sigma_{\text{add}}$ (ng/mL) | 1.9e-08 (3.73e-07)<br>[0, 7.51e-07] | $\sigma_{\text{add}}$ (ng/mL) | 0 (fixed) |
| $\sigma_{\text{prop}}$ (%CV) | 50.3 (1.4)<br>[47.6, 53.0] | $\sigma_{\text{prop}}$ (%CV) | 50.3 (1.6)<br>[47.2, 53.4] | $\sigma_{\text{prop}}$ (%CV) | 50.5 (1.6)<br>[47.5, 53.5] |

(B) Estimates from categorical gene models for *UGT1A4* or *UGT2B10*.
| ***UGT1A4* Categorical Gene Model**<br>(Obj = -1918.3; BICc = -1832.4) |  | ***UGT2B10* Categorical Gene Model**<br>(Obj = -1913.8; BICc = -1827.9) |  |
| --- | --- | --- | --- |
| Parameters | Estimates (SE)<br>[95% CI] | Parameters | Estimates (SE)<br>[95% CI] |
| $CL = \theta_1(WT/70)^{0.75} \left( \frac{1}{1 + \left( \frac{TM_{50}}{PMA} \right)^{Hill}} \right) \exp(\theta_5 I[UGT1A4 > 0])$ | | $CL = \theta_1(WT/70)^{0.75} \left( \frac{1}{1 + \left( \frac{TM_{50}}{PMA} \right)^{Hill}} \right) \exp(\theta_5 I[UGT2B10 > 0])$ | |
| $\theta_1$ | 24.0 (2.19)<br>[20.1, 28.7] | $\theta_1$ | 20.4 (2.07)<br>[16.8, 24.9] |
| $TM_{50}$ | 42.2 (0.041)<br>[42.1, 42.3] | $TM_{50}$ | 45.7 (0.078)<br>[45.6, 45.9] |
| Hill | 4.17 (0.0016)<br>[4.16, 4.17] | Hill | 4.96 (0.0073)<br>[4.95, 4.97] |
| $\theta_5$ | -0.221 (0.16)<br>[-0.54, 0.09] | $\theta_5$ | -0.104 (0.11)<br>[-0.32, 0.11] |
| $V_1 = \theta_2(WT/70)$ | | $V_1 = \theta_2(WT/70)$ | |
| $\theta_2$ | 169 (20.4)<br>[133, 213] | $\theta_2$ | 178 (12.6) [155,<br>204] |
| $Q = \theta_3(WT/70)^{0.75}$ | | $Q = \theta_3(WT/70)^{0.75}$ | |
| $\theta_3$ | 31.2 (2.74)<br>[26.3, 37.1] | $\theta_3$ | 33.2 (1.84)<br>[29.8, 37.0] |
| $V_2 = \theta_4(WT/70)$ | | $V_2 = \theta_4(WT/70)$ | |
| $\theta_4$ | 14346 (2782)<br>[9908, 20770] | $\theta_4$ | 19655 (2804)<br>[14921, 25889] |
| $\omega_{CL}$ (%CV) | 113 (13)<br>[90, 140] | $\omega_{CL}$ (%CV) | 131 (13)<br>[108, 158] |
| $\omega_{V1}$ (%CV) | 139 (31)<br>[88, 216] | $\omega_{V1}$ (%CV) | 126 (12)<br>[104, 152] |
| $\omega_Q$ (%CV) | 67 (7)<br>[54, 81] | $\omega_Q$ (%CV) | 67 (6)<br>[56, 79] |
| $\omega_{V2}$ (%CV) | 640 (158)<br>[404, 1067] | $\omega_{V2}$ (%CV) | 544 (114)<br>[367, 837] |
| $\rho_{CL,V1}$ | 0.953 (0.043)<br>[0.869, 1.0] | $\rho_{CL,V1}$ | 0.939 (0.018)<br>[0.904, 0.974] |
| $\rho_{CL,V2}$ | 0 (fixed) | $\rho_{CL,V2}$ | 0 (fixed) |
| $\rho_{CL,Q}$ | 0 (fixed) | $\rho_{CL,Q}$ | 0 (fixed) |
| $\rho_{V1,V2}$ | 0 (fixed) | $\rho_{V1,V2}$ | 0 (fixed) |
| $\rho_{V1,Q}$ | 0 (fixed) | $\rho_{V1,Q}$ | 0 (fixed) |
| $\rho_{V2,Q}$ | 0 (fixed) | $\rho_{V2,Q}$ | 0 (fixed) |
| $\sigma_{\text{add}}$ (ng/mL) | 0 (fixed) | $\sigma_{\text{add}}$ (ng/mL) | 0 (fixed) |
| $\sigma_{\text{prop}}$ (%CV) | 50.6 (1.6)<br>[47.5, 53.8] | $\sigma_{\text{prop}}$ (%CV) | 50.7 (1.6) [47.6,<br>53.8] |

(C) Estimates from additive gene models for *UGT1A4* or *UGT2B10*.
| ***UGT1A4* Additive Gene Model**<br>(Obj = -1917.6; BICc = -1831.7) |  | ***UGT2B10* Additive Gene Model**<br>(Obj = -1917.3; BICc = -1831.4) |  |
| --- | --- | --- | --- |
| Parameters | Estimates (SE)<br>[95% CI] | Parameters | Estimates (SE)<br>[95% CI] |
| $CL = \theta_1(WT/70)^{0.75} \left( \frac{1}{1 + \left( \frac{TM_{50}}{PMA} \right)^{Hill}} \right) \exp(\theta_5 UGT1A4)$ | | $CL = \theta_1(WT/70)^{0.75} \left( \frac{1}{1 + \left( \frac{TM_{50}}{PMA} \right)^{Hill}} \right) \exp(\theta_5 UGT2B10)$ | |
| $\theta_1$ | 23.8 (2.47)<br>[19.5, 29.1] | $\theta_1$ | 23.5 (2.43)<br>[19.2, 28.7] |
| $TM_{50}$ | 47.6 (0.136)<br>[47.3, 47.9] | $TM_{50}$ | 41.6 (0.206)<br>[41.2, 42.0] |
| Hill | 5.55 (0.013)<br>[5.53, 5.58] | Hill | 17.6 (0.052)<br>[17.5, 17.7] |
| $\theta_5$ | -0.166 (0.12)<br>[-0.406, 0.073] | $\theta_5$ | -0.108 (0.068)<br>[-0.241, 0.025] |
| $V_1 = \theta_2(WT/70)$ | | $V_1 = \theta_2(WT/70)$ | |
| $\theta_2$ | 171 (16.2)<br>[142, 206] | $\theta_2$ | 161 (14.2)<br>[136, 191] |
| $Q = \theta_3(WT/70)^{0.75}$ | | $Q = \theta_3(WT/70)^{0.75}$ | |
| $\theta_3$ | 31.6 (2.1)<br>[27.7, 36.0] | $\theta_3$ | 31.5 (2.2)<br>[27.5, 36.0] |
| $V_2 = \theta_4(WT/70)$ | | $V_2 = \theta_4(WT/70)$ | |
| $\theta_4$ | 16576 (3229)<br>[11431, 24038] | $\theta_4$ | 12588 (2603) [8495,<br>18654] |
| $\omega_{CL}$ (%CV) | 110 (10)<br>[91, 131] | $\omega_{CL}$ (%CV) | 109 (12)<br>[88, 135] |
| $\omega_{V1}$ (%CV) | 136 (18)<br>[104, 177] | $\omega_{V1}$ (%CV) | 139 (14)<br>[113, 170] |
| $\omega_Q$ (%CV) | 69 (7)<br>[56, 84] | $\omega_Q$ (%CV) | 64 (7)<br>[52, 78] |
| $\omega_{V2}$ (%CV) | 548 (120)<br>[364, 858] | $\omega_{V2}$ (%CV) | 792 (212)<br>[482, 1377] |
| $\rho_{CL,V1}$ | 0.933 (0.03)<br>[0.88, 0.99] | $\rho_{CL,V1}$ | 0.943 (0.04)<br>[0.87, 1.0] |
| $\rho_{CL,V2}$ | 0 (fixed) | $\rho_{CL,V2}$ | 0 (fixed) |
| $\rho_{CL,Q}$ | 0 (fixed) | $\rho_{CL,Q}$ | 0 (fixed) |
| $\rho_{V1,V2}$ | 0 (fixed) | $\rho_{V1,V2}$ | 0 (fixed) |
| $\rho_{V1,Q}$ | 0 (fixed) | $\rho_{V1,Q}$ | 0 (fixed) |
| $\rho_{V2,Q}$ | 0 (fixed) | $\rho_{V2,Q}$ | 0 (fixed) |
| $\sigma_{\text{add}}$ (ng/mL) | 0 (fixed) | $\sigma_{\text{add}}$ (ng/mL) | 0 (fixed) |
| $\sigma_{\text{prop}}$ (%CV) | 50.4 (1.6)<br>[47.4, 53.5] | $\sigma_{\text{prop}}$ (%CV) | 50.6 (1.6)<br>[47.5, 53.7] |

(D) Estimates from models using weight and maturation with simplified variance structure in *CYP2A6* PRS subset.
| Without *CYP2A6* Score<br>(Obj = -1889.2; BICc = -1809.3) |  | With *CYP2A6* Score<br>(Obj = -1889.9; BICc = -1804.2) |  |
| --- | --- | --- | --- |
| Parameters | Estimates (SE)<br>[95% CI] | Parameters | Estimates (SE)<br>[95% CI] |
| $CL = \theta_1(WT/70)^{0.75} \left( \frac{1}{1 + \left( \frac{TM_{50}}{PMA} \right)^{Hill}} \right)$ | | $CL = \theta_1(WT/70)^{0.75} \left( \frac{1}{1 + \left( \frac{TM_{50}}{PMA} \right)^{Hill}} \right) \exp(\theta_5(CY2A6score))$ | |
| $\theta_1$ | 30.5 (2.16)<br>[26.5, 35.0] | $\theta_1$ | 24.8 (14.7)<br>[9.42, 65.2] |
| $TM_{50}$ | 41.5 (0.046)<br>[41.4, 41.6] | $TM_{50}$ | 42.6 (0.098)<br>[42.4, 42.8] |
| Hill | 4.52 (0.011)<br>[4.50, 4.54] | Hill | 7.45 (0.015)<br>[7.42, 7.48] |
| | | $\theta_5$ | 0.0885 (0.28)<br>[-0.46, 0.64] |
| $V_1 = \theta_2(WT/70)$ | | $V_1 = \theta_2(WT/70)$ | |
| $\theta_2$ | 157 (16.1)<br>[128, 191] | $\theta_2$ | 152 (12.1) [130, 178] |
| $Q = \theta_3(WT/70)^{0.75}$ | | $Q = \theta_3(WT/70)^{0.75}$ | |
| $\theta_3$ | 24.8 (1.99)<br>[21.2, 29.0] | $\theta_3$ | 24.5 (1.78)<br>[21.2, 28.2] |
| $V_2 = \theta_4(WT/70)$ | | $V_2 = \theta_4(WT/70)$ | |
| $\theta_4$ | 5720 (1129)<br>[3925, 8335] | $\theta_4$ | 5756 (1017)<br>[4102, 8077] |
| $\omega_{CL}$ (%CV) | 103 (10)<br>[86, 124] | $\omega_{CL}$ (%CV) | 100 (8)<br>[86, 116] |
| $\omega_{V1}$ (%CV) | 129 (20)<br>[95, 173] | $\omega_{V1}$ (%CV) | 138 (15)<br>[111, 171] |
| $\omega_Q$ (%CV) | 87 (10)<br>[69, 109] | $\omega_Q$ (%CV) | 86 (9)<br>[69, 106] |
| $\omega_{V2}$ (%CV) | 514 (118)<br>[334, 825] | $\omega_{V2}$ (%CV) | 549 (133)<br>[349, 906] |
| $\rho_{CL,V1}$ | 0.945 (0.03)<br>[0.88, 1.0] | $\rho_{CL,V1}$ | 0.942 (0.02)<br>[0.91, 0.98] |
| $\rho_{CL,V2}$ | 0 (fixed) | $\rho_{CL,V2}$ | 0 (fixed) |
| $\rho_{CL,Q}$ | 0 (fixed) | $\rho_{CL,Q}$ | 0 (fixed) |
| $\rho_{V1,V2}$ | 0 (fixed) | $\rho_{V1,V2}$ | 0 (fixed) |
| $\rho_{V1,Q}$ | 0 (fixed) | $\rho_{V1,Q}$ | 0 (fixed) |
| $\rho_{V2,Q}$ | 0 (fixed) | $\rho_{V2,Q}$ | 0 (fixed) |
| $\sigma_{add}$ (ng/mL) | 0 (fixed) | $\sigma_{add}$ (ng/mL) | 0 (fixed) |
| $\sigma_{prop}$ (%CV) | 50.8 (1.6)<br>[47.6, 54.0] | $\sigma_{prop}$ (%CV) | 50.7 (1.6)<br>[47.7, 53.8] |

Abbreviations: <sup>a</sup> 95% Asymptotic confidence intervals (CIs); SE, standard error; Obj, objective function value; BICc, corrected Bayesian information criteria; CL, total clearance (L/hr); Q, intercompartmental clearance (L/hr); V<sub>1</sub>, volume of distribution for the central compartment (L); V<sub>2</sub>, volume of distribution for the peripheral compartment (L); TM<sub>50</sub> postmenstrual age at which clearance is 50% of adult value; Hill, maturation factor slope coefficient; CV, coefficient of variation; WT, body weight in kg; PMA, postmenstrual age in weeks; $\omega_{CL}$ , $\omega_{V1}$ , $\omega_Q$ , $\omega_{V2}$ , the standard deviation for $\eta_i^{CL}$ , $\eta_i^{V1}$ , $\eta_i^Q$ , and $\eta_i^{V2}$ , respectively; For the standard deviation of random effects, $\omega$ , coefficient of variation was calculated as $CV\% = 100 \times \sqrt{\exp(\omega^2) - 1}$ ; $\rho$ are correlation terms between random effects; $\sigma_{prop}$ and $\sigma_{add}$ are proportional and additive residual error terms.

Including weight as covariate for all PK parameters with fixed allometric scaling parameters substantially improved the model fit (both OFV and BICc decreased by 406 from the base model, **Table 2A**) and plots of individual predicted vs. observed concentration, individual weighted residuals vs. predicted concentration and random effects vs. covariates also improved (**Supplemental Figures S8 – S12**). The CV for CL, V_1_, Q, and V_2_ were 123%, 168%, 91%, and 857%, respectively. Using estimated allometric parameters did not improve the model fit.

Among the four models adjusting for both weight and age maturation, the model with sigmoid postmenstrual age maturation had the largest improvement in BICc compared to the model with only weight (difference of 8.2, **Table S5**). This model was further simplified by estimating models with fixed effects for V_2_ or V_2_ and Q, no additive residual error component, and several correlation structures for the covariance between random effects (**Table S6**). The final model without genetic effects includes proportional residual error, fixed allometric scaling for all parameters and sigmoid (Hill) maturation for total clearance, random effects for all PK parameters and correlation only between the random effects of CL and V_1_ (**Table 2A**). No further improvement was seen by adding other covariates including either form of *UGT1A4* or *UGT2B10* or predicted *CYP2A6* activity score (Tables **2B, 2C**, and **2D**) and no strong covariate effects were seen for V_1_, Q, or V_2_ based on graphical goodness-of-fit plots. The final covariate model is described as follows:

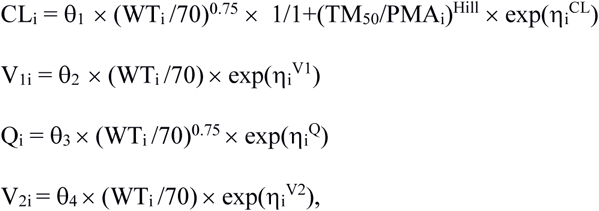

where CL_i_, V_1i_, Q_i_, and V_2i_ are the individual-specific PK parameters corresponding to CL, V_1_, Q, and V_2_, WT_i_ is subject weight in kilograms (kg), and PMA_i_ is subject postmenstrual age in weeks. The 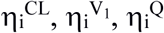, and 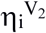 are random effects explaining inter-individual variability for the PK parameters which follow a normal distribution with mean zero and variance of ω^2^_CL_, ω^2^_V1_, ω^2^_Q_, and ω^2^_V2_, respectively. The θs are estimated model parameters. Diagnostic plots for the final model are shown in **Figure 2**. The plot of observed dexmedetomidine concentrations vs. population predictions reflects the relatively large inter-individual variability and potential misspecification for small concentration values, however no major deficiencies in the structural or error models are seen when comparing observed and individual predicted concentrations. No trends were detected in plots of individual weighted residuals vs. predicted concentration or time. The prediction-corrected visual predictive check showed good agreement between the observed and theoretical median and 90^th^ percentiles; model misspecification is seen for the 10^th^ percentile where many values are below the LLOQ and were simulated from the estimated model and where data are sparse (e.g., times greater than 5 days after first dose).

**Figure 2.**
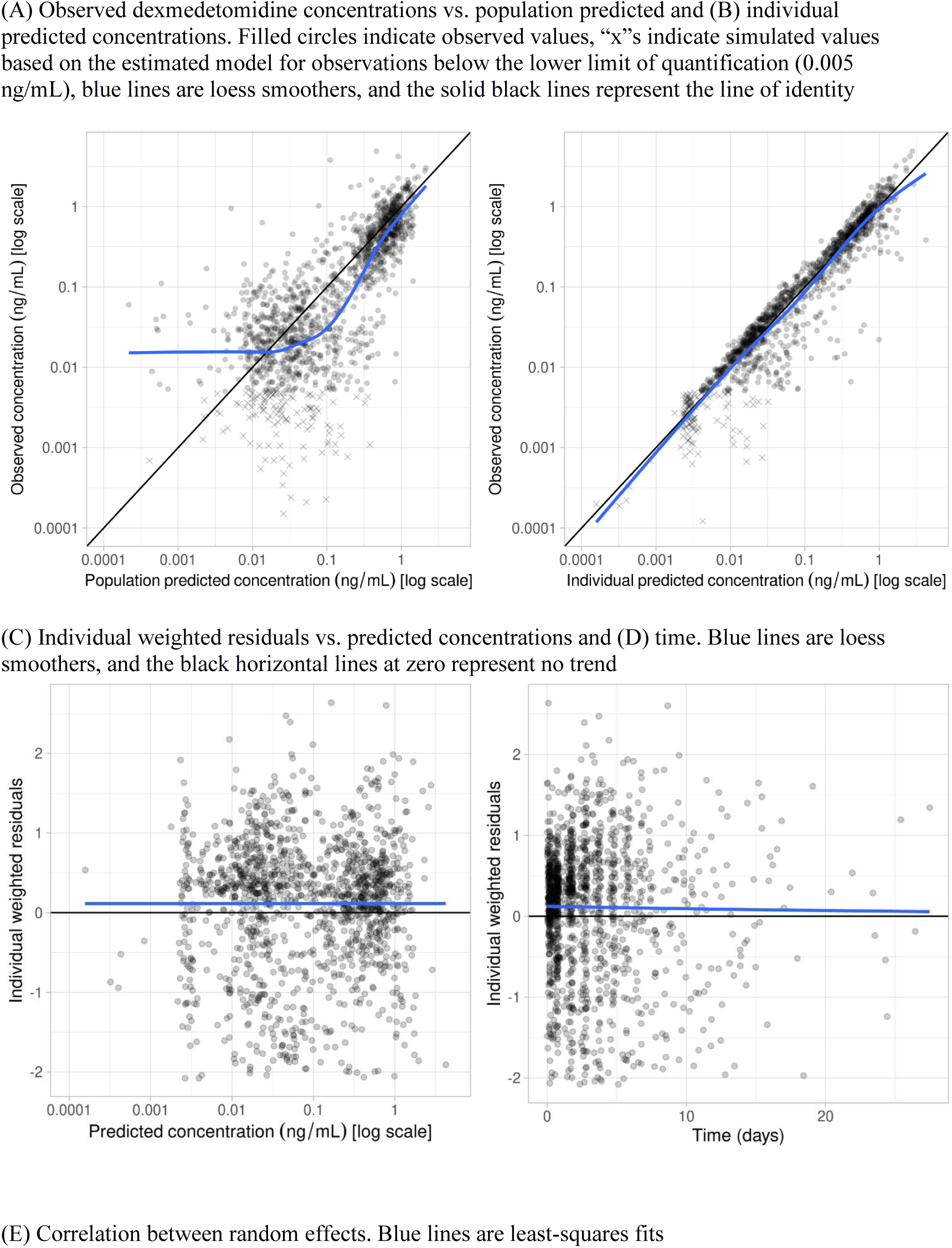
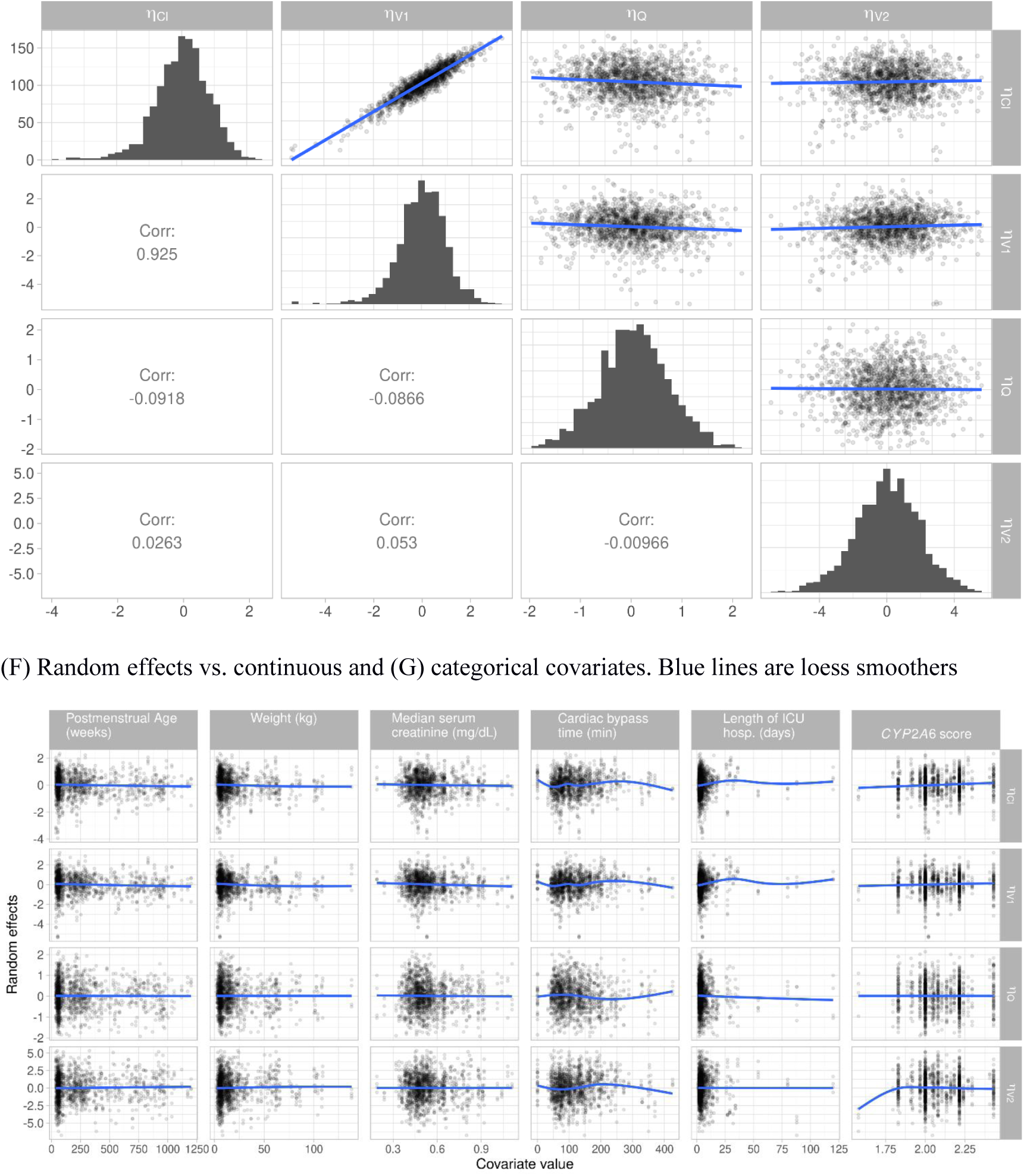
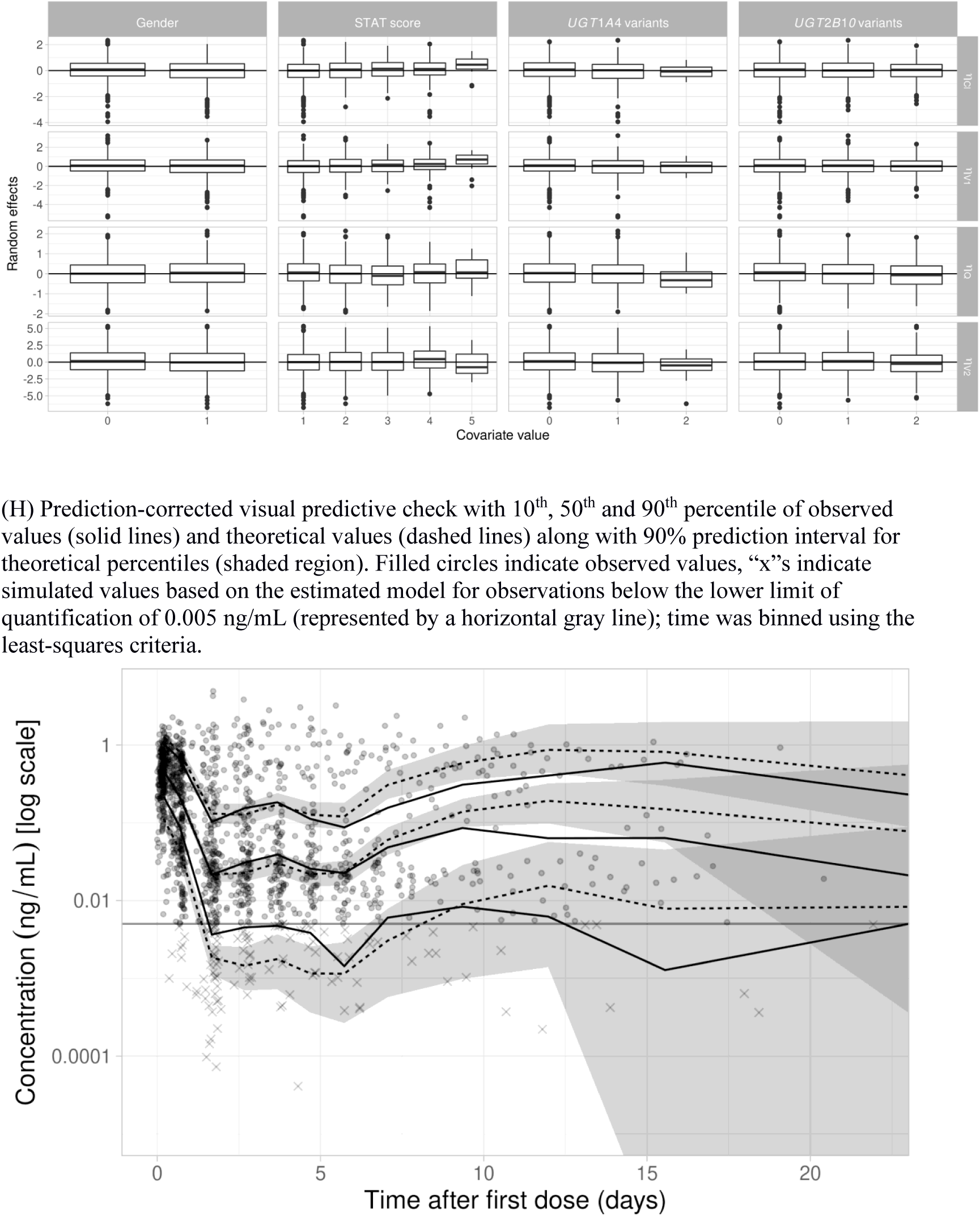
Diagnostic plots for the final population PK model.

The estimates of CL, V_1_, Q and V_2_ in terms of a standard weight of 70 kg are shown in **Table 2A**: CL (θ_1_) = 27.3 L/h, V_1_ (θ_2_) = 161 L, Q (θ_3_) = 26.0 L/h, and V_2_ (θ_4_) = 7903 L. We estimate CL, V_1_, Q and V_2_ as 6.04 L/h, 21.6 L, 5.7 L/h, and 1061.26 L for a child at the median weight of 9.4 kg and median postmenstrual age of 104.6 weeks; After including covariates, the CV for CL was substantially reduced from 201% estimated in the base model to 123% in the weight only model to 103% in the final model. CV remains large for some parameters, especially V_2_ (624%) and V_1_ (138%), indicating that we lack the data to estimate them with precision.

Model estimated clearance from the final model for seven age groups across a range of plausible weights is shown in **Figure 3** (overlap between lines indicates weights that are plausible for multiple age groups). Weight impacts mean estimated CL for all ages while postmenstrual age has a large impact only for the youngest age groups. For those over 93 weeks postmenstrual age, maturation is near adult level and mean estimated CL is primarily determined by weight.

**Figure 3.**
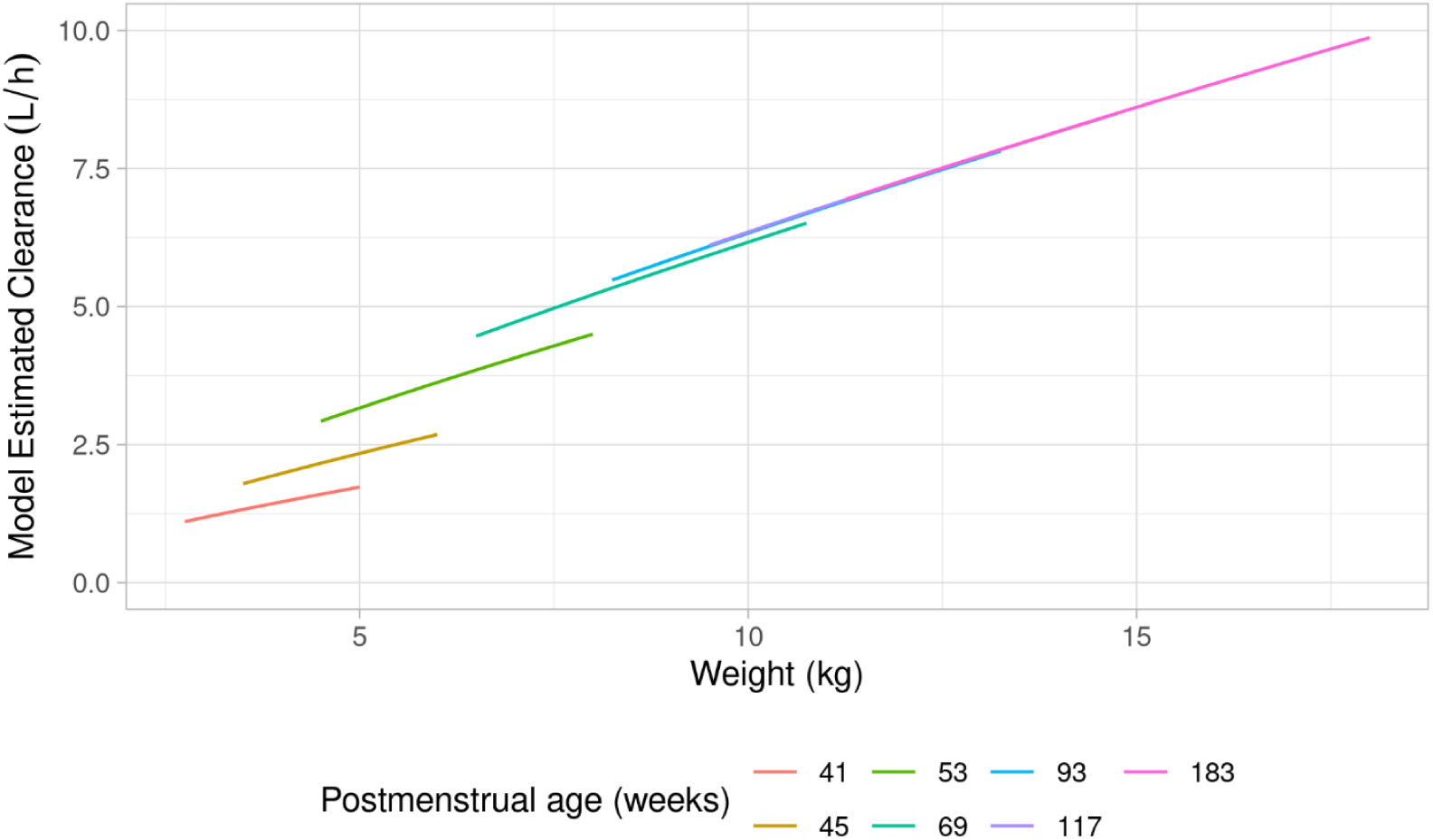
Predicted clearance by weight for selected ages from the final weight and age maturation model. Plausible weight ranges for each age group are: 41 weeks (2.7 to 5.1 kg), 45 weeks (3.3 to 6.1 kg), 53 weeks (4.5 to 8.0 kg), 69 weeks (6.3 to 10.8 kg), 93 weeks (8.1 to 13.4 kg), 117 weeks (9.3 to 14.9 kg), 183 weeks (11.2 to 18.1 kg). Overlapping lines between different age categories represent weights that are plausible for multiple age groups.

### Genetic Effects on Clearance and Concentration

*UGT1A4, UGT2B10*, and *CYP2A6* were not significant at the α = 0.05 level. For the *UGT1A4* categorical gene model the estimated effect of any variants vs. no variants was −0.221 (95% CI: −0.54 to 0.09) for a 20% decrease [exp(−0.221) ≈ 0.80] in CL on average for those with any *UGT1A4* variants holding age and weight constant. In the *UGT1A4* additive gene model, the estimated effect of each additional variant was −0.166 (95% CI: −0.406 to 0.073). For the *UGT2B10* categorical model, the estimated genotype effect was −0.104 (95% CI: −0.32 to 0.11), indicating a 10% decrease on average. The *UGT2B10* additive model estimated the effect of additional variants as −0.108 (95% CI: −0.241 to 0.025). For the *CYP2A6* model, the estimated effect of a unit increase in risk score was 0.0885 (95% CI: −0.46 to 0.64).

Although these effects are not statistically significant using the α = 0.05 threshold, we perform simulations to assess the hypothetical impact on total clearance and clinical dosing if the categorical *UGT1A4* or *UGT2B10* model estimates were utilized. Results are included in **Supplemental Figures S29 – S34**. Including these effects in the PK model has a negligible impact on dosing.

## Discussion

Using remnant specimens along with dosing, clinical, and demographic information from an EHR system we were able to develop a dexmedetomidine population PK model for a large pediatric cohort of 354 patients. We identified patient characteristics that alter the PK profile. This study is one of the largest pediatric dexmedetomidine population PK studies reported.

We confirmed a structural model and covariate relationships which are in line with those previously reported for dexmedetomidine PK. Specifically, our model included both weight and age maturation effects on CL. We estimated a weight-standardized CL of 27.3 L/h (CV 103%). Our estimated CL is somewhat smaller (with larger CV) than those reported in other pediatric PK studies. For a standard weight of 70 kg, Potts et al.^17^ found a population CL estimate of 42.1 L/h (CV 30.9%); including a scaling factor of 0.73 for children given infusion (vs. bolus) reduced the CL estimate to 30.7 L/h. Zuppa et al.^9^ estimated CL of 37.3 L/h (CV 48%) for neonates and infants age 0 – 6 months after cardiac bypass and Su et al.^15^ estimated CL as 39.4 L/h (CV 28%) for children age 1 – 24 months after open heart surgery. The discrepancy between studies could be related to several factors including study design and study population. For example, our study used sparse and opportunistic sampling and included a more heterogenous population which included older children while the other studies used densely measured drug levels and were performed in a well-controlled clinical setting with a younger and more homogeneous population. After controlling for weight and age maturation, we found little evidence to support the importance of UGT1A4, UGT2B10, or CYP2A6 effects in explaining variability of CL between subjects.

Using population PK models derived from EHR data and remnant specimens offers the possibility of more accurate prediction of individual dosing requirements in a real-life setting, especially in populations where large, intensive-sampling PK clinical trials are difficult to perform due to ethical or logistical considerations. The results from such model-informed precision dosing could also be integrated into EHR-embedded decision support tools; the development and implementation of several of these tools has been recently described by Mizuno et al.^41^ and Vinks et al.^42^

There are several limitations related to the use of EHR and remnant specimens for our study. Although our data were generated using a standardized system to construct the PK data,^25^ there may be some errors due to inherent limitations of EHR data, which is not primarily collected for research use. First, data collected for clinical purposes may be subject to errors related to data entry or missingness. Further, real-world dosing data are not standardized with large heterogeneity in the frequency, duration, and timing of administered infusion and bolus doses. In addition, the specimens are very sparse for some subjects and their collection is not timed to facilitate optimal PK estimation. These limitations may be related to the imprecision in estimates for some PK parameters, notably V_2_. Future studies could address some of these limitations by incorporating prior information from previous or smaller pilot studies with more densely sampled data.

Despite these limitations, our study provides further evidence for the feasibility of using EHR data and remnant specimens for population PK analysis. Our study findings, such as weight effects on CL, could be helpful to develop a model-based dosing that may be superior to the current fixed weight-based dosing scheme. However, this should be tested in a future study for its clinical utility in the pediatric population. Because dexmedetomidine is used to achieve specific sedation goals, it would also be of interest to incorporate the current study results into a joint pharmacokinetic-pharmacodynamic model using sedation outcomes also derived from the EHR. These models are an important step toward the ultimate goal of precision dosing.

## Data Availability

Data is not publicly available.

## Acknowledgements

The authors thank the Vanderbilt Clinical Laboratory staff for their assistance in obtaining the remnant specimens for this research, the patients and families who participated in this study, Ahmed Elboraie and Rachel Tyndale for assistance with the *CYP2A6* genetic risk score, and Cole Beck for assistance with the concomitant medications.

## Author Contributions

All authors participated in critical review and revision of the final manuscript and approved the final manuscript draft.

NTJ: analyzed data, performed research, and wrote manuscript. JHB: analyzed data.

RC: designed research and contributed analytical tools. TE: performed research.

BH: designed research, performed research and contributed analytical tools. PJK: designed research and wrote manuscript. JK: performed research. MDM: extracted data and analyzed data.

SLV: designed research, performed research and wrote manuscript. LC: designed research, analyzed data, performed research, and wrote manuscript.

## Data Availability Statement

The data that support the findings of this study are available from the corresponding author upon reasonable request.

