## Supplemental Information, Tables, and Figures for "Population Pharmacokinetic Analysis of Dexmedetomidine in Children using Real World Data from Electronic Health Records and Remnant Specimens"

### SAEM algorithm parameters

The SAEM algorithm was used with the following settings:

Burn-in phase (prior SAEM): 5

Exploratory phase with auto-stop (max iterations: 2000, min iterations 200, stepsize exponent: 0) and simulated annealing (decreasing rate for variance of residual errors: 0.95, decreasing rate for variance of individual parameters: 0.95)

Smoothing phase with auto-stop (max iterations: 400, min iterations 200, stepsize exponent: 0.7)

All other settings used Monolix defaults.

### Supplemental Tables

**Table S1.** Postnatal age categories

| | Entire cohort |
|---|---|
| n | 354 |
| Postnatal Age | |
| <6 mo | 111 (31%) |
| 6 - 24 mo | 75 (21%) |
| 2 - 13 yr | 134 (38%) |
| 14 - 18 yr | 25 (7%) |
| >18 yr | 9 (3%) |

**Table S2.** Concomitant medications administered to at least 5% of participants

| Medication Name | Count | Percent |
|---|---|---|
| acetaminophen | 322 | 92.5 |
| cefazolin | 320 | 92.0 |
| famotidine | 310 | 89.1 |
| morphine | 308 | 88.5 |
| furosemide | 280 | 80.5 |
| fentanyl | 271 | 77.9 |
| rocuronium | 242 | 69.5 |
| oxycodone | 208 | 59.8 |
| heparin | 203 | 58.3 |
| lorazepam | 179 | 51.4 |
| ketorolac | 162 | 46.6 |
| midazolam | 160 | 46.0 |
| nicardipine | 155 | 44.5 |
| chlorothiazide | 151 | 43.4 |
| milrinone | 129 | 37.1 |
| epinephrine | 119 | 34.2 |
| ondansetron | 116 | 33.3 |
| hydromorphone | 114 | 32.8 |
| docusate | 93 | 26.7 |
| aminocaprioc | 92 | 26.4 |
| aspirin | 85 | 24.4 |
| dexamethasone | 83 | 23.9 |
| phenylephrine | 77 | 22.1 |

| Medication Name | Count | Percent |
|---|---|---|
| vecuronium | 74 | 21.3 |
| vancomycin | 59 | 17.0 |
| aminocaproic | 53 | 15.2 |
| hydrocortisone | 52 | 14.9 |
| nitroprusside | 52 | 14.9 |
| spironolactone | 47 | 13.5 |
| enalapril | 43 | 12.4 |
| ketamine | 43 | 12.4 |
| omeprazole | 43 | 12.4 |
| diphenhydramine | 39 | 11.2 |
| cefepime | 35 | 10.1 |
| ephedrine | 35 | 10.1 |
| albuterol | 33 | 9.5 |
| propofol | 32 | 9.2 |
| dopamine | 30 | 8.6 |
| glycopyrrolate | 30 | 8.6 |
| vasopressin | 27 | 7.8 |
| ibuprofen | 22 | 6.3 |
| neostigmine | 22 | 6.3 |
| lidocaine | 21 | 6.0 |
| lisinopril | 19 | 5.5 |
| captopril | 18 | 5.2 |

Note: Concomitant medications unavailable for 6 participants in final study cohort.

**Table S3.** Tests of deviation from Hardy Weinberg Equilibrium

| **rs ID** | ***p* - value** |
|---|---|
| rs2942857 | 0.518922 |
| rs112561475 | 0.762602 |
| rs61750900 | 0.825786 |
| rs2011425 | 0.586849 |
| rs3892221 | 0.500481 |
| rs6755571 | 0.53469 |
| rs56113850 | 0.466614 |
| rs2316204 | 0.762141 |
| rs113288603 | 0.656682 |
| rs28399442 | 0.122089 |
| rs1801272 | 0.312122 |
| rs28399433 | 0.615292 |

### Stage 1 – Find adequate non-genotype covariate PK model

**Table S3.** Base models (model with **bold text** included in main results Table 2)

1. One compartment model with combined additive and proportional error

**2. Two compartment model with combined additive and proportional error**

3. Two compartment model with additive error

4. Two compartment model with proportional error

| Model | Structural Model | Residual variability | Submodels | Between subject variability | OFV | BICc |
|---|---|---|---|---|---|---|
| 1 | One compartment multidose IV infusion/bolus with linear elimination | $y = C(\psi; x)(1+\varepsilon_1)+\varepsilon_2$<br>$\varepsilon_1 \sim N(0, \sigma^2_{\text{prop}})$<br>$\varepsilon_2 \sim N(0, \sigma^2_{\text{add}})$ | $\psi_i = \{Cl_i, V_i\}$<br>$Cl_i = \theta_1 \exp(\eta_{Cli})$<br>$V_i = \theta_2 \exp(\eta_{Vi})$<br><br>$\eta_{Cli} \sim N(0, \omega^2_{Cl})$; $\eta_{Vi} \sim N(0, \omega^2_V)$ | $\Omega = \begin{bmatrix} \omega^2_{Cl} & \omega_{Cl,V} \\ & \omega^2_V \end{bmatrix}$ | -961.3 | -914.7 |
| 2 | Two compartment multidose IV infusion/bolus with linear elimination | $y = C(\psi; x)(1+\varepsilon_1)+\varepsilon_2$<br>$\varepsilon_1 \sim N(0, \sigma^2_{\text{prop}})$<br>$\varepsilon_2 \sim N(0, \sigma^2_{\text{add}})$ | $\psi_i = \{Cl_i, V_{1i}, Q_i, V_{2i}\}$<br>$Cl_i = \theta_1 \exp(\eta_{Cli})$<br>$V_{1i} = \theta_2 \exp(\eta_{V1i})$<br>$Q_i = \theta_3 \exp(\eta_{Qi})$<br>$V_{2i} = \theta_4 \exp(\eta_{V2i})$<br><br>$\eta_{Cli} \sim N(0, \omega^2_{Cl})$; $\eta_{V1i} \sim N(0, \omega^2_{V1})$<br>$\eta_{Qi} \sim N(0, \omega^2_Q)$; $\eta_{V2i} \sim N(0, \omega^2_{V2})$ | $\Omega = \begin{bmatrix} \omega^2_{Cl} & \omega_{Cl,V_1} & \omega_{Cl,Q} & \omega_{Cl,V_2} \\ & \omega^2_{V_1} & \omega_{V_1,Q} & \omega_{V_1,V_2} \\ & & \omega^2_Q & \omega_{Q,V_2} \\ & & & \omega^2_{V_2} \end{bmatrix}$ | -1517.3 | -1415.1 |
| 3 | Two compartment multidose IV infusion/bolus with linear elimination | $y = C(\psi; x)+\varepsilon$<br>$\varepsilon \sim N(0, \sigma^2_{\text{add}})$ | $\psi_i = \{Cl_i, V_{1i}, Q_i, V_{2i}\}$<br>$Cl_i = \theta_1 \exp(\eta_{Cli})$<br>$V_{1i} = \theta_2 \exp(\eta_{V1i})$<br>$Q_i = \theta_3 \exp(\eta_{Qi})$<br>$V_{2i} = \theta_4 \exp(\eta_{V2i})$<br><br>$\eta_{Cli} \sim N(0, \omega^2_{Cl})$; $\eta_{V1i} \sim N(0, \omega^2_{V1})$<br>$\eta_{Qi} \sim N(0, \omega^2_Q)$; $\eta_{V2i} \sim N(0, \omega^2_{V2})$ | $\Omega = \begin{bmatrix} \omega^2_{Cl} & \omega_{Cl,V_1} & \omega_{Cl,Q} & \omega_{Cl,V_2} \\ & \omega^2_{V_1} & \omega_{V_1,Q} & \omega_{V_1,V_2} \\ & & \omega^2_Q & \omega_{Q,V_2} \\ & & & \omega^2_{V_2} \end{bmatrix}$ | 1018.7 | 1113.6 |

| Model | Structural Model | Residual variability | Submodels | Between subject variability | OFV | BICc |
|---|---|---|---|---|---|---|
| 4 | Two compartment multidose IV infusion/bolus with linear elimination | $y = C(\psi; x)(1+\varepsilon)$<br>$\varepsilon \sim N(0, \sigma^2_{\text{prop}})$ | $\psi_i = \{Cl_i, V_{1i}, Q_i, V_{2i}\}$<br>$Cl_i = \theta_1 \exp(\eta_{Cli})$<br>$V_{1i} = \theta_2 \exp(\eta_{V1i})$<br>$Q_i = \theta_3 \exp(\eta_{Qi})$<br>$V_{2i} = \theta_4 \exp(\eta_{V2i})$<br><br>$\eta_{Cli} \sim N(0, \omega^2_{Cl});\ \eta_{V1i} \sim N(0, \omega^2_{V1})$<br>$\eta_{Qi} \sim N(0, \omega^2_{Q});\ \eta_{V2i} \sim N(0, \omega^2_{V2})$ | $\Omega = \begin{bmatrix} \omega^2_{Cl} & \omega_{Cl,V_1} & \omega_{Cl,Q} & \omega_{Cl,V_2} \\ & \omega^2_{V_1} & \omega_{V_1,Q} & \omega_{V_1,V_2} \\ & & \omega^2_{Q} & \omega_{Q,V_2} \\ & & & \omega^2_{V_2} \end{bmatrix}$ | -1352.5 | -1257.6 |

**Table S4.** Allometric scaling models (model with **bold text** included in main results Table 2)

**5. Two compartment model with combined error and fixed theory-based allometric scaling parameters**

6. Two compartment model with combined error and estimated allometric scaling parameters

| Model | Structural Model | Residual variability | Submodels | Between subject variability | OFV | BICc |
|---|---|---|---|---|---|---|
| 5 | Two compartment multidose IV infusion/bolus with linear elimination | $y = C(\psi; x)(1+\varepsilon_1)+\varepsilon_2$<br>$\varepsilon_1 \sim N(0, \sigma^2_{\text{prop}})$<br>$\varepsilon_2 \sim N(0, \sigma^2_{\text{add}})$ | $\psi_i = \{Cl_i, V_{1i}, Q_i, V_{2i}\}$<br>$Cl_i = \theta_1(WT/70)^{0.75}\exp(\eta_{Cli})$<br>$V_{1i} = \theta_2(WT/70)\ \exp(\eta_{V1i})$<br>$Q_i = \theta_3(WT/70)^{0.75}\exp(\eta_{Qi})$<br>$V_{2i} = \theta_4(WT/70)\ \exp(\eta_{V2i})$<br><br>$\eta_{Cli} \sim N(0, \omega^2_{Cl});\ \eta_{V1i} \sim N(0, \omega^2_{V1})$<br>$\eta_{Qi} \sim N(0, \omega^2_{Q});\ \eta_{V2i} \sim N(0, \omega^2_{V2})$ | $\Omega = \begin{bmatrix} \omega^2_{Cl} & \omega_{Cl,V_1} & \omega_{Cl,Q} & \omega_{Cl,V_2} \\ & \omega^2_{V_1} & \omega_{V_1,Q} & \omega_{V_1,V_2} \\ & & \omega^2_{Q} & \omega_{Q,V_2} \\ & & & \omega^2_{V_2} \end{bmatrix}$ | -1923.3 | -1821.1 |
| 6 | Two compartment multidose IV infusion/bolus with linear elimination | $y = C(\psi; x)(1+\varepsilon_1)+\varepsilon_2$<br>$\varepsilon_1 \sim N(0, \sigma^2_{\text{prop}})$<br>$\varepsilon_2 \sim N(0, \sigma^2_{\text{add}})$ | $\psi_i = \{Cl_i, V_{1i}, Q_i, V_{2i}\}$<br>$Cl_i = \theta_1(WT/70)^{\beta_1}\exp(\eta_{Cli})$<br>$V_{1i} = \theta_2(WT/70)^{\beta_2}\exp(\eta_{V1i})$<br>$Q_i = \theta_3(WT/70)^{\beta_3}\exp(\eta_{Qi})$<br>$V_{2i} = \theta_4(WT/70)^{\beta_4}\exp(\eta_{V2i})$<br><br>$\eta_{Cli} \sim N(0, \omega^2_{Cl});\ \eta_{V1i} \sim N(0, \omega^2_{V1})$<br>$\eta_{Qi} \sim N(0, \omega^2_{Q});\ \eta_{V2i} \sim N(0, \omega^2_{V2})$ | $\Omega = \begin{bmatrix} \omega^2_{Cl} & \omega_{Cl,V_1} & \omega_{Cl,Q} & \omega_{Cl,V_2} \\ & \omega^2_{V_1} & \omega_{V_1,Q} & \omega_{V_1,V_2} \\ & & \omega^2_{Q} & \omega_{Q,V_2} \\ & & & \omega^2_{V_2} \end{bmatrix}$ | -1735.5 | -1604.3 |

**Table S5.** Allometric scaling and maturation models

7. Two compartment model with combined error and fixed theory-based allometric scaling parameters and exponential age

8. Two compartment model with combined error and fixed theory-based allometric scaling parameters and sigmoid (Hill) maturation

9. Two compartment model with combined error and bodyweight-dependent allometric scaling parameter for total clearance

10. Two compartment model with combined error and age-dependent allometric scaling parameter for total clearance

| Model | Structural Model | Residual variability | Submodels | Between subject variability | OFV | BICc |
|---|---|---|---|---|---|---|
| 7 | Two compartment multidose IV infusion/bolus with linear elimination | $y = C(\psi; x)(1+\varepsilon_1)+\varepsilon_2$<br>$\varepsilon_1 \sim N(0, \sigma^2_{\text{prop}})$<br>$\varepsilon_2 \sim N(0, \sigma^2_{\text{add}})$ | $\psi_i = \{Cl_i, V_{1i}, Q_i, V_{2i}\}$<br>$Cl_i = \theta_1 (WT/70)^{0.75} \exp(\theta_5 AGE + \eta_{Cli})$<br>$V_{1i} = \theta_2 (WT/70) \ \exp(\eta_{V1i})$<br>$Q_i = \theta_3 (WT/70)^{0.75} \exp(\eta_{Qi})$<br>$V_{2i} = \theta_4 (WT/70) \ \exp(\eta_{V2i})$<br><br>$\eta_{Cli} \sim N(0, \omega^2_{Cl});\ \eta_{V1i} \sim N(0, \omega^2_{V1})$<br>$\eta_{Qi} \sim N(0, \omega^2_{Q});\ \eta_{V2i} \sim N(0, \omega^2_{V2})$ | $\Omega = \begin{bmatrix} \omega^2_{Cl} & \omega_{Cl,V_1} & \omega_{Cl,Q} & \omega_{Cl,V_2} \\ & \omega^2_{V_1} & \omega_{V_1,Q} & \omega_{V_1,V_2} \\ & & \omega^2_Q & \omega_{Q,V_2} \\ & & & \omega^2_{V_2} \end{bmatrix}$ | -1932.3 | -1824.3 |
| 8 | Two compartment multidose IV infusion/bolus with linear elimination | $y = C(\psi; x)(1+\varepsilon_1)+\varepsilon_2$<br>$\varepsilon_1 \sim N(0, \sigma^2_{\text{prop}})$<br>$\varepsilon_2 \sim N(0, \sigma^2_{\text{add}})$ | $\psi_i = \{Cl_i, V_{1i}, Q_i, V_{2i}\}$<br>$Cl_i = \theta_1 (WT/70)^{0.75} \left( \frac{1}{1+\left(\frac{TM_{50}}{PMA}\right)^{Hill}} \right) \exp(\eta_{Cli})$<br>$V_{1i} = \theta_2 (WT/70) \ \exp(\eta_{V1i})$<br>$Q_i = \theta_3 (WT/70)^{0.75} \exp(\eta_{Qi})$<br>$V_{2i} = \theta_4 (WT/70) \ \exp(\eta_{V2i})$<br><br>$\eta_{Cli} \sim N(0, \omega^2_{Cl});\ \eta_{V1i} \sim N(0, \omega^2_{V1})$<br>$\eta_{Qi} \sim N(0, \omega^2_{Q});\ \eta_{V2i} \sim N(0, \omega^2_{V2})$ | $\Omega = \begin{bmatrix} \omega^2_{Cl} & \omega_{Cl,V_1} & \omega_{Cl,Q} & \omega_{Cl,V_2} \\ & \omega^2_{V_1} & \omega_{V_1,Q} & \omega_{V_1,V_2} \\ & & \omega^2_Q & \omega_{Q,V_2} \\ & & & \omega^2_{V_2} \end{bmatrix}$ | -1945.9 | -1829.3 |

| Model | Structural Model | Residual variability | Submodels | Between subject variability | OFV | BICc |
|---|---|---|---|---|---|---|
| 9 | Two compartment multidose IV infusion/bolus with linear elimination | $y = C(\psi; x)(1+\varepsilon_1)+\varepsilon_2$<br>$\varepsilon_1 \sim N(0, \sigma^2_{\text{prop}})$<br>$\varepsilon_2 \sim N(0, \sigma^2_{\text{add}})$ | $\psi_i = \{Cl_i, V_{1i}, Q_i, V_{2i}\}$<br>$Cl_i = \theta_1 (WT/70)^{k_1} \exp(\eta_{Cli})$<br>$k_1 = k_0 - \frac{k_{max} WT^{Hill}}{k_{50}^{Hill} + WT^{Hill}}$<br>$V_{1i} = \theta_2 (WT/70) \ \exp(\eta_{V1i})$<br>$Q_i = \theta_3 (WT/70)^{0.75} \exp(\eta_{Qi})$<br>$V_{2i} = \theta_4 (WT/70) \ \exp(\eta_{V2i})$<br>$\eta_{Cli} \sim N(0, \omega^2_{Cl});\ \eta_{V1i} \sim N(0, \omega^2_{V1})$<br>$\eta_{Qi} \sim N(0, \omega^2_{Q});\ \eta_{V2i} \sim N(0, \omega^2_{V2})$ | $\Omega = \begin{bmatrix} \omega^2_{Cl} & \omega_{Cl,V_1} & \omega_{Cl,Q} & \omega_{Cl,V_2} \\ & \omega^2_{V_1} & \omega_{V_1,Q} & \omega_{V_1,V_2} \\ & & \omega^2_Q & \omega_{Q,V_2} \\ & & & \omega^2_{V_2} \end{bmatrix}$ | -1946.9 | -1815.8 |
| 10 | Two compartment multidose IV infusion/bolus with linear elimination | $y = C(\psi; x)(1+\varepsilon_1)+\varepsilon_2$<br>$\varepsilon_1 \sim N(0, \sigma^2_{\text{prop}})$<br>$\varepsilon_2 \sim N(0, \sigma^2_{\text{add}})$ | $\psi_i = \{Cl_i, V_{1i}, Q_i, V_{2i}\}$<br>$Cl_i = \theta_1 (WT/70)^{k_1} \exp(\eta_{Cli})$<br>$k_1 = k_0 - \frac{k_{max} AGE^{Hill}}{k_{50}^{Hill} + AGE^{Hill}}$<br>$V_{1i} = \theta_2 (WT/70) \ \exp(\eta_{V1i})$<br>$Q_i = \theta_3 (WT/70)^{0.75} \exp(\eta_{Qi})$<br>$V_{2i} = \theta_4 (WT/70) \ \exp(\eta_{V2i})$<br>$\eta_{Cli} \sim N(0, \omega^2_{Cl});\ \eta_{V1i} \sim N(0, \omega^2_{V1})$<br>$\eta_{Qi} \sim N(0, \omega^2_{Q});\ \eta_{V2i} \sim N(0, \omega^2_{V2})$ | $\Omega = \begin{bmatrix} \omega^2_{Cl} & \omega_{Cl,V_1} & \omega_{Cl,Q} & \omega_{Cl,V_2} \\ & \omega^2_{V_1} & \omega_{V_1,Q} & \omega_{V_1,V_2} \\ & & \omega^2_Q & \omega_{Q,V_2} \\ & & & \omega^2_{V_2} \end{bmatrix}$ | -1939.1 | -1808.0 |

**Table S6.** Simplified variance component models (model with **bold text** included in main results Table 2)

11. Two compartment model with combined error and fixed theory-based allometric scaling parameters and sigmoid (Hill) maturation, no $V_2$ random effects

12. Two compartment model with combined error and fixed theory-based allometric scaling parameters and sigmoid (Hill) maturation, no $V_2$ or Q random effects

13. Two compartment model with proportional error and fixed theory-based allometric scaling parameters and sigmoid (Hill) maturation, all random effects

14. Two compartment model with proportional error and fixed theory-based allometric scaling parameters and sigmoid (Hill) maturation, no $V_2$ random effects

15. Two compartment model with proportional error and fixed theory-based allometric scaling parameters and sigmoid (Hill) maturation, block correlation 1

**16. Two compartment model with proportional error and fixed theory-based allometric scaling parameters and sigmoid (Hill) maturation, block correlation 2**

| Model | Structural Model | Residual variability | Submodels | Between subject variability | OFV | BICc |
|---|---|---|---|---|---|---|
| 11 | Two compartment multidose IV infusion/bolus with linear elimination | $y = C(\psi; x)(1+\varepsilon_1) + \varepsilon_2$<br>$\varepsilon_1 \sim N(0, \sigma^2_{\text{prop}})$<br>$\varepsilon_2 \sim N(0, \sigma^2_{\text{add}})$ | $\psi_i = \{Cl_i, V_{1i}, Q_i, V_{2i}\}$<br>$Cl_i = \theta_1 (WT/70)^{0.75} \left( \frac{1}{1+\left(\frac{TM_{50}}{PMA}\right)^{Hill}} \right) \exp(\eta_{Cli})$<br>$V_{1i} = \theta_2 (WT/70) \ \exp(\eta_{V1i})$<br>$Q_i = \theta_3 (WT/70)^{0.75} \exp(\eta_{Qi})$<br>$V_{2i} = \theta_4 (WT/70)$<br>$\eta_{Cli} \sim N(0, \omega^2_{Cl});\ \eta_{V1i} \sim N(0, \omega^2_{V1})$<br>$\eta_{Qi} \sim N(0, \omega^2_Q)$ | $\Omega = \begin{bmatrix} \omega^2_{Cl} & \omega_{Cl,V_1} & \omega_{Cl,Q} \\ & \omega^2_{V_1} & \omega_{V_1,Q} \\ & & \omega^2_Q \end{bmatrix}$ | -1504.7 | -1411.6 |

| Model | Structural Model | Residual variability | Submodels | Between subject variability | OFV | BICc |
|---|---|---|---|---|---|---|
| 12 | Two compartment multidose IV infusion/bolus with linear elimination | $y = C(\psi; x)(1+\varepsilon_1)+\varepsilon_2$<br>$\varepsilon_1 \sim N(0, \sigma^2_{\text{prop}})$<br>$\varepsilon_2 \sim N(0, \sigma^2_{\text{add}})$ | $\psi_i = \{Cl_i, V_{1i}, Q_i, V_{2i}\}$<br>$Cl_i = \theta_1 (WT/70)^{0.75}\left(\frac{1}{1+\left(\frac{TM_{50}}{PMA}\right)^{Hill}}\right)\exp(\eta_{Cli})$<br>$V_{1i} = \theta_2(WT/70)\ \exp(\eta_{V1i})$<br>$Q_i = \theta_3(WT/70)^{0.75}$<br>$V_{2i} = \theta_4(WT/70)$<br>$\eta_{Cli} \sim N(0, \omega^2_{Cl});\ \eta_{V1i} \sim N(0, \omega^2_{V1})$ | $\Omega = \begin{bmatrix} \omega^2_{Cl} & \omega_{Cl,V_1} \\ & \omega^2_{V_1} \end{bmatrix}$ | -1408.4 | -1332.9 |
| 13 | Two compartment multidose IV infusion/bolus with linear elimination | $y = C(\psi; x)(1+\varepsilon)$<br>$\varepsilon \sim N(0, \sigma^2_{\text{prop}})$ | $\psi_i = \{Cl_i, V_{1i}, Q_i, V_{2i}\}$<br>$Cl_i = \theta_1 (WT/70)^{0.75}\left(\frac{1}{1+\left(\frac{TM_{50}}{PMA}\right)^{Hill}}\right)\exp(\eta_{Cli})$<br>$V_{1i} = \theta_2(WT/70)\ \exp(\eta_{V1i})$<br>$Q_i = \theta_3(WT/70)^{0.75}\exp(\eta_{Qi})$<br>$V_{2i} = \theta_4(WT/70)\ \exp(\eta_{V2i})$<br>$\eta_{Cli} \sim N(0, \omega^2_{Cl});\ \eta_{V1i} \sim N(0, \omega^2_{V1})$<br>$\eta_{Qi} \sim N(0, \omega^2_{Q});\ \eta_{V2i} \sim N(0, \omega^2_{V2})$ | $\Omega = \begin{bmatrix} \omega^2_{Cl} & \omega_{Cl,V_1} & \omega_{Cl,Q} & \omega_{Cl,V_2} \\ & \omega^2_{V_1} & \omega_{V_1,Q} & \omega_{V_1,V_2} \\ & & \omega^2_{Q} & \omega_{Q,V_2} \\ & & & \omega^2_{V_2} \end{bmatrix}$ | -1932.9 | -1823.5 |
| 14 | Two compartment multidose IV infusion/bolus with linear elimination | $y = C(\psi; x)(1+\varepsilon)$<br>$\varepsilon \sim N(0, \sigma^2_{\text{prop}})$ | $\psi_i = \{Cl_i, V_{1i}, Q_i, V_{2i}\}$<br>$Cl_i = \theta_1 (WT/70)^{0.75}\left(\frac{1}{1+\left(\frac{TM_{50}}{PMA}\right)^{Hill}}\right)\exp(\eta_{Cli})$<br>$V_{1i} = \theta_2(WT/70)\ \exp(\eta_{V1i})$<br>$Q_i = \theta_3(WT/70)^{0.75}\exp(\eta_{Qi})$<br>$V_{2i} = \theta_4(WT/70)$<br>$\eta_{Cli} \sim N(0, \omega^2_{Cl});\ \eta_{V1i} \sim N(0, \omega^2_{V1})$<br>$\eta_{Qi} \sim N(0, \omega^2_{Q})$ | $\Omega = \begin{bmatrix} \omega^2_{Cl} & \omega_{Cl,V_1} & \omega_{Cl,Q} \\ & \omega^2_{V_1} & \omega_{V_1,Q} \\ & & \omega^2_{Q} \end{bmatrix}$ | -1479.6 | -1393.6 |

| Model | Structural Model | Residual variability | Submodels | Between subject variability | OFV | BICc |
|---|---|---|---|---|---|---|
| 15 | Two compartment multidose IV infusion/bolus with linear elimination | $y = C(\psi; x)(1+\varepsilon)$<br>$\varepsilon \sim N(0, \sigma^2_{\text{prop}})$ | $\psi_i = \{Cl_i, V_{1i}, Q_i, V_{2i}\}$<br>$Cl_i = \theta_1 (WT/70)^{0.75} \left( \frac{1}{1+\left(\frac{TM_{50}}{PMA}\right)^{Hill}} \right) \exp(\eta_{Cli})$<br>$V_{1i} = \theta_2 (WT/70) \exp(\eta_{V1i})$<br>$Q_i = \theta_3 (WT/70)^{0.75} \exp(\eta_{Qi})$<br>$V_{2i} = \theta_4 (WT/70) \exp(\eta_{V2i})$<br>$\eta_{Cli} \sim N(0, \omega^2_{Cl});\ \eta_{V1i} \sim N(0, \omega^2_{V1})$<br>$\eta_{Qi} \sim N(0, \omega^2_{Q});\ \eta_{V2i} \sim N(0, \omega^2_{V2})$ | $\Omega = \begin{bmatrix} \omega^2_{Cl} & \omega_{Cl,V_1} & 0 & 0 \\ & \omega^2_{V_1} & 0 & 0 \\ & & \omega^2_{Q} & \omega_{Q,V_2} \\ & & & \omega^2_{V_2} \end{bmatrix}$ | -1915.2 | -1829.3 |
| 16 | Two compartment multidose IV infusion/bolus with linear elimination | $y = C(\psi; x)(1+\varepsilon)$<br>$\varepsilon \sim N(0, \sigma^2_{\text{prop}})$ | $\psi_i = \{Cl_i, V_{1i}, Q_i, V_{2i}\}$<br>$Cl_i = \theta_1 (WT/70)^{0.75} \left( \frac{1}{1+\left(\frac{TM_{50}}{PMA}\right)^{Hill}} \right) \exp(\eta_{Cli})$<br>$V_{1i} = \theta_2 (WT/70) \exp(\eta_{V1i})$<br>$Q_i = \theta_3 (WT/70)^{0.75} \exp(\eta_{Qi})$<br>$V_{2i} = \theta_4 (WT/70) \exp(\eta_{V2i})$<br>$\eta_{Cli} \sim N(0, \omega^2_{Cl});\ \eta_{V1i} \sim N(0, \omega^2_{V1})$<br>$\eta_{Qi} \sim N(0, \omega^2_{Q});\ \eta_{V2i} \sim N(0, \omega^2_{V2})$ | $\Omega = \begin{bmatrix} \omega^2_{Cl} & \omega_{Cl,V_1} & 0 & 0 \\ & \omega^2_{V_1} & 0 & 0 \\ & & \omega^2_{Q} & 0 \\ & & & \omega^2_{V_2} \end{bmatrix}$ | -1915.1 | -1835.0 |

**Table S7.** Additional simplified variance components models

17. Two compartment model with proportional error and estimated allometric scaling parameters, block correlation 2

18. Two compartment model with proportional error and estimated allometric scaling parameters and exponential age, block correlation 2

19. Two compartment model with proportional error and fixed theory-based allometric scaling parameters and exponential age, block correlation 2

20. Two compartment model with proportional error and estimate allometric scaling parameters and sigmoid (Hill) maturation, block correlation 2

| Model | Structural Model | Residual variability | Submodels | Between subject variability | OFV | BICc |
|---|---|---|---|---|---|---|
| 17 | Two compartment multidose IV infusion/bolus with linear elimination | $y = C(\psi; x)(1+\varepsilon)$<br>$\varepsilon \sim N(0, \sigma^2_{\text{prop}})$ | $\psi_i = \{Cl_i, V_{1i}, Q_i, V_{2i}\}$<br>$Cl_i = \theta_1 (WT/70)^{\beta_{Cl}} \exp(\eta_{Cli})$<br>$V_{1i} = \theta_2 (WT/70)^{\beta_{V1}} \exp(\eta_{V1i})$<br>$Q_i = \theta_3 (WT/70)^{\beta_Q} \exp(\eta_{Qi})$<br>$V_{2i} = \theta_4 (WT/70)^{\beta_{V2}} \exp(\eta_{V2i})$<br><br>$\eta_{Cli} \sim N(0, \omega^2_{Cl});\ \eta_{V1i} \sim N(0, \omega^2_{V1})$<br>$\eta_{Qi} \sim N(0, \omega^2_Q);\ \eta_{V2i} \sim N(0, \omega^2_{V2})$ | $\Omega = \begin{bmatrix} \omega^2_{Cl} & \omega_{Cl,V_1} & 0 & 0 \\ & \omega^2_{V_1} & 0 & 0 \\ & & \omega^2_Q & 0 \\ & & & \omega^2_{V_2} \end{bmatrix}$ | -1910.1 | -1815.5 |
| 18 | Two compartment multidose IV infusion/bolus with linear elimination | $y = C(\psi; x)(1+\varepsilon)$<br>$\varepsilon \sim N(0, \sigma^2_{\text{prop}})$ | $\psi_i = \{Cl_i, V_{1i}, Q_i, V_{2i}\}$<br>$Cl_i = \theta_1 (WT/70)^{\beta_{Cl}} \exp(\theta_{Cl,age} age + \eta_{Cli})$<br>$V_{1i} = \theta_2 (WT/70)^{\beta_{V1}} \exp(\eta_{V1i})$<br>$Q_i = \theta_3 (WT/70)^{\beta_Q} \exp(\eta_{Qi})$<br>$V_{2i} = \theta_4 (WT/70)^{\beta_{V2}} \exp(\eta_{V2i})$<br><br>$\eta_{Cli} \sim N(0, \omega^2_{Cl});\ \eta_{V1i} \sim N(0, \omega^2_{V1})$<br>$\eta_{Qi} \sim N(0, \omega^2_Q);\ \eta_{V2i} \sim N(0, \omega^2_{V2})$ | $\Omega = \begin{bmatrix} \omega^2_{Cl} & \omega_{Cl,V_1} & 0 & 0 \\ & \omega^2_{V_1} & 0 & 0 \\ & & \omega^2_Q & 0 \\ & & & \omega^2_{V_2} \end{bmatrix}$ | -1904.0 | -1803.6 |

| Model | Structural Model | Residual variability | Submodels | Between subject variability | OFV | BICc |
|---|---|---|---|---|---|---|
| 19 | Two compartment multidose IV infusion/bolus with linear elimination | $y = C(\psi; x)(1+\varepsilon)$<br>$\varepsilon \sim N(0, \sigma^2_{\text{prop}})$ | $\psi_i = \{Cl_i, V_{1i}, Q_i, V_{2i}\}$<br>$Cl_i = \theta_1(WT/70)^{0.75}\exp(\theta_{Cl,age}\,age + \eta_{Cli})$<br>$V_{1i} = \theta_2(WT/70)\ \exp(\eta_{V1i})$<br>$Q_i = \theta_3(WT/70)^{0.75}\exp(\eta_{Qi})$<br>$V_{2i} = \theta_4(WT/70)\ \exp(\eta_{V2i})$<br><br>$\eta_{Cli} \sim N(0, \omega^2_{Cl});\ \eta_{V1i} \sim N(0, \omega^2_{V1})$<br>$\eta_{Qi} \sim N(0, \omega^2_{Q});\ \eta_{V2i} \sim N(0, \omega^2_{V2})$ | $\Omega = \begin{bmatrix} \omega^2_{Cl} & \omega_{Cl,V_1} & 0 & 0 \\ & \omega^2_{V_1} & 0 & 0 \\ & & \omega^2_{Q} & 0 \\ & & & \omega^2_{V_2} \end{bmatrix}$ | -1899.1 | -1827.6 |
| 20 | Two compartment multidose IV infusion/bolus with linear elimination | $y = C(\psi; x)(1+\varepsilon)$<br>$\varepsilon \sim N(0, \sigma^2_{\text{prop}})$ | $\psi_i = \{Cl_i, V_{1i}, Q_i, V_{2i}\}$<br>$Cl_i = \theta_1(WT/70)^{\beta_{Cl}}\left(\frac{1}{1+\left(\frac{TM_{50}}{PMA}\right)^{Hill}}\right)\exp(\eta_{Cli})$<br>$V_{1i} = \theta_2(WT/70)^{\beta_{V1}}\exp(\eta_{V1i})$<br>$Q_i = \theta_3(WT/70)^{\beta_{Q}}\exp(\eta_{Qi})$<br>$V_{2i} = \theta_4(WT/70)^{\beta_{V2}}\exp(\eta_{V2i})$<br><br>$\eta_{Cli} \sim N(0, \omega^2_{Cl});\ \eta_{V1i} \sim N(0, \omega^2_{V1})$<br>$\eta_{Qi} \sim N(0, \omega^2_{Q});\ \eta_{V2i} \sim N(0, \omega^2_{V2})$ | $\Omega = \begin{bmatrix} \omega^2_{Cl} & \omega_{Cl,V_1} & 0 & 0 \\ & \omega^2_{V_1} & 0 & 0 \\ & & \omega^2_{Q} & 0 \\ & & & \omega^2_{V_2} \end{bmatrix}$ | -1927.4 | -1818.4 |

**Table S8.** Additional non-genotype covariate models

21. Add exponential gender effect to Model 16

22. Add exponential serum creatinine effect to Model 16

23. Add exponential cardiac bypass time effect to Model 16

24. Add exponential STAT score effect to Model 16

25. Add exponential ICU hospitalization length effect to Model 16

| Model | Structural Model | Residual variability | Submodels | Between subject variability | OFV | BICc |
|---|---|---|---|---|---|---|
| 21 | Two compartment multidose IV infusion/bolus with linear elimination | $y = C(\psi; x)(1+\varepsilon)$ <br> $\varepsilon \sim N(0, \sigma^2_{\text{prop}})$ | $\psi_i = \{Cl_i, V_{1i}, Q_i, V_{2i}\}$ <br> $Cl_i = \theta_1 (WT/70)^{0.75} \left( \frac{1}{1+\left(\frac{TM_{50}}{PMA}\right)^{Hill}} \right) \exp(\theta_{Cl,gender} I[female] + \eta_{Cli})$ <br> $V_{1i} = \theta_2 (WT/70) \ \exp(\eta_{V1i})$ <br> $Q_i = \theta_3 (WT/70)^{0.75} \exp(\eta_{Qi})$ <br> $V_{2i} = \theta_4 (WT/70) \ \exp(\eta_{V2i})$ <br> $\eta_{Cli} \sim N(0, \omega^2_{Cl});\ \eta_{V1i} \sim N(0, \omega^2_{V1})$ <br> $\eta_{Qi} \sim N(0, \omega^2_{Q});\ \eta_{V2i} \sim N(0, \omega^2_{V2})$ | $\Omega = \begin{bmatrix} \omega^2_{Cl} & \omega_{Cl,V_1} & 0 & 0 \\ & \omega^2_{V_1} & 0 & 0 \\ & & \omega^2_Q & 0 \\ & & & \omega^2_{V_2} \end{bmatrix}$ | -1914.1 | -1828.2 |
| 22 | Two compartment multidose IV infusion/bolus with linear elimination | $y = C(\psi; x)(1+\varepsilon)$ <br> $\varepsilon \sim N(0, \sigma^2_{\text{prop}})$ | $\psi_i = \{Cl_i, V_{1i}, Q_i, V_{2i}\}$ <br> $Cl_i = \theta_1 (WT/70)^{0.75} \left( \frac{1}{1+\left(\frac{TM_{50}}{PMA}\right)^{Hill}} \right) \exp(\theta_{Cl,creat} creat + \eta_{Cli})$ <br> $V_{1i} = \theta_2 (WT/70) \ \exp(\eta_{V1i})$ <br> $Q_i = \theta_3 (WT/70)^{0.75} \exp(\eta_{Qi})$ <br> $V_{2i} = \theta_4 (WT/70) \ \exp(\eta_{V2i})$ <br> $\eta_{Cli} \sim N(0, \omega^2_{Cl});\ \eta_{V1i} \sim N(0, \omega^2_{V1})$ <br> $\eta_{Qi} \sim N(0, \omega^2_{Q});\ \eta_{V2i} \sim N(0, \omega^2_{V2})$ | $\Omega = \begin{bmatrix} \omega^2_{Cl} & \omega_{Cl,V_1} & 0 & 0 \\ & \omega^2_{V_1} & 0 & 0 \\ & & \omega^2_Q & 0 \\ & & & \omega^2_{V_2} \end{bmatrix}$ | -1913.1 | -1827.2 |

| Model | Structural Model | Residual variability | Submodels | Between subject variability | OFV | BICc |
|---|---|---|---|---|---|---|
| 23 | Two compartment multidose IV infusion/bolus with linear elimination | $y = C(\psi; x)(1+\varepsilon)$<br>$\varepsilon \sim N(0, \sigma^2_{\text{prop}})$ | $\psi_i = \{Cl_i, V_{1i}, Q_i, V_{2i}\}$<br>$Cl_i = \theta_1 (WT/70)^{0.75} \left( \frac{1}{1+\left(\frac{TM_{50}}{PMA}\right)^{Hill}} \right) \exp(\theta_{Cl,cpb}(bypasstime) + \eta_{Cli})$<br>$V_{1i} = \theta_2 (WT/70) \ \exp(\eta_{V1i})$<br>$Q_i = \theta_3 (WT/70)^{0.75} \exp(\eta_{Qi})$<br>$V_{2i} = \theta_4 (WT/70) \ \exp(\eta_{V2i})$<br>$\eta_{Cli} \sim N(0, \omega^2_{Cl});\ \eta_{V1i} \sim N(0, \omega^2_{V1})$<br>$\eta_{Qi} \sim N(0, \omega^2_{Q});\ \eta_{V2i} \sim N(0, \omega^2_{V2})$ | $\Omega = \begin{bmatrix} \omega^2_{Cl} & \omega_{Cl,V_1} & 0 & 0 \\ & \omega^2_{V_1} & 0 & 0 \\ & & \omega^2_{Q} & 0 \\ & & & \omega^2_{V_2} \end{bmatrix}$ | -1913.4 | -1827.5 |
| 24 | Two compartment multidose IV infusion/bolus with linear elimination | $y = C(\psi; x)(1+\varepsilon)$<br>$\varepsilon \sim N(0, \sigma^2_{\text{prop}})$ | $\psi_i = \{Cl_i, V_{1i}, Q_i, V_{2i}\}$<br>$Cl_i = \theta_1 (WT/70)^{0.75} \left( \frac{1}{1+\left(\frac{TM_{50}}{PMA}\right)^{Hill}} \right) \exp(\theta_{Cl,STAT}(STATscore) + \eta_{Cli})$<br>$V_{1i} = \theta_2 (WT/70) \ \exp(\eta_{V1i})$<br>$Q_i = \theta_3 (WT/70)^{0.75} \exp(\eta_{Qi})$<br>$V_{2i} = \theta_4 (WT/70) \ \exp(\eta_{V2i})$<br>$\eta_{Cli} \sim N(0, \omega^2_{Cl});\ \eta_{V1i} \sim N(0, \omega^2_{V1})$<br>$\eta_{Qi} \sim N(0, \omega^2_{Q});\ \eta_{V2i} \sim N(0, \omega^2_{V2})$ | $\Omega = \begin{bmatrix} \omega^2_{Cl} & \omega_{Cl,V_1} & 0 & 0 \\ & \omega^2_{V_1} & 0 & 0 \\ & & \omega^2_{Q} & 0 \\ & & & \omega^2_{V_2} \end{bmatrix}$ | -1914.9 | -1829.0 |
| 25 | Two compartment multidose IV infusion/bolus with linear elimination | $y = C(\psi; x)(1+\varepsilon)$<br>$\varepsilon \sim N(0, \sigma^2_{\text{prop}})$ | $\psi_i = \{Cl_i, V_{1i}, Q_i, V_{2i}\}$<br>$Cl_i = \theta_1 (WT/70)^{0.75} \left( \frac{1}{1+\left(\frac{TM_{50}}{PMA}\right)^{Hill}} \right) \exp(\theta_{Cl,loi}(ICU\ time) + \eta_{Cli})$<br>$V_{1i} = \theta_2 (WT/70) \ \exp(\eta_{V1i})$<br>$Q_i = \theta_3 (WT/70)^{0.75} \exp(\eta_{Qi})$<br>$V_{2i} = \theta_4 (WT/70) \ \exp(\eta_{V2i})$<br>$\eta_{Cli} \sim N(0, \omega^2_{Cl});\ \eta_{V1i} \sim N(0, \omega^2_{V1})$<br>$\eta_{Qi} \sim N(0, \omega^2_{Q});\ \eta_{V2i} \sim N(0, \omega^2_{V2})$ | $\Omega = \begin{bmatrix} \omega^2_{Cl} & \omega_{Cl,V_1} & 0 & 0 \\ & \omega^2_{V_1} & 0 & 0 \\ & & \omega^2_{Q} & 0 \\ & & & \omega^2_{V_2} \end{bmatrix}$ | -1913.9 | -1827.9 |

### Stage 2 – Test for improvement in best covariate model by adding genotype effects

**Table S9.** Best covariate model and UGT* genotype effects (models with **bold text** included in main results Table 2)

**26. Add exponential UGT2B10 categorical effect (no variants vs. any variants) to Model 16**

**27. Add exponential UGT2B10 additive effect to Model 16**

**28. Add exponential UGT1A4 categorical effect (no variants vs. any variants) to Model 16**

**29. Add exponential UGT1A4 additive effect to Model 16**

| Model | Structural Model | Residual variability | Submodels | Between subject variability | OFV | BICc |
|---|---|---|---|---|---|---|
| 26 | Two compartment multidose IV infusion/bolus with linear elimination | $y = C(\psi;x)(1+\varepsilon)$<br>$\varepsilon \sim N(0,\sigma^2_{\text{prop}})$ | $\psi_i = \{Cl_i, V_{1i}, Q_i, V_{2i}\}$<br>$Cl_i = \theta_1 (WT/70)^{0.75}\left(\frac{1}{1+\left(\frac{TM_{50}}{PMA}\right)^{Hill}}\right)\exp(\theta_{Cl,UGT2B10} I[UGT2B10>0]+\eta_{Cli})$<br>$V_{1i} = \theta_2 (WT/70) \exp(\eta_{V1i})$<br>$Q_i = \theta_3 (WT/70)^{0.75}\exp(\eta_{Qi})$<br>$V_{2i} = \theta_4 (WT/70) \exp(\eta_{V2i})$<br>$\eta_{Cli} \sim N(0,\omega^2_{Cl});\ \eta_{V1i} \sim N(0,\omega^2_{V1})$<br>$\eta_{Qi} \sim N(0,\omega^2_{Q});\ \eta_{V2i} \sim N(0,\omega^2_{V2})$ | $\Omega = \begin{bmatrix} \omega^2_{Cl} & \omega_{Cl,V_1} & 0 & 0 \\ & \omega^2_{V_1} & 0 & 0 \\ & & \omega^2_Q & 0 \\ & & & \omega^2_{V_2} \end{bmatrix}$ | -1913.8 | -1827.9 |
| 27 | Two compartment multidose IV infusion/bolus with linear elimination | $y = C(\psi;x)(1+\varepsilon)$<br>$\varepsilon \sim N(0,\sigma^2_{\text{prop}})$ | $\psi_i = \{Cl_i, V_{1i}, Q_i, V_{2i}\}$<br>$Cl_i = \theta_1 (WT/70)^{0.75}\left(\frac{1}{1+\left(\frac{TM_{50}}{PMA}\right)^{Hill}}\right)\exp(\theta_{Cl,UGT2B10} UGT2B10+\eta_{Cli})$<br>$V_{1i} = \theta_2 (WT/70) \exp(\eta_{V1i})$<br>$Q_i = \theta_3 (WT/70)^{0.75}\exp(\eta_{Qi})$<br>$V_{2i} = \theta_4 (WT/70) \exp(\eta_{V2i})$<br>$\eta_{Cli} \sim N(0,\omega^2_{Cl});\ \eta_{V1i} \sim N(0,\omega^2_{V1})$<br>$\eta_{Qi} \sim N(0,\omega^2_{Q});\ \eta_{V2i} \sim N(0,\omega^2_{V2})$ | $\Omega = \begin{bmatrix} \omega^2_{Cl} & \omega_{Cl,V_1} & 0 & 0 \\ & \omega^2_{V_1} & 0 & 0 \\ & & \omega^2_Q & 0 \\ & & & \omega^2_{V_2} \end{bmatrix}$ | -1917.3 | -1831.4 |

| Model | Structural Model | Residual variability | Submodels | Between subject variability | OFV | BICc |
|---|---|---|---|---|---|---|
| 28 | Two compartment multidose IV infusion/bolus with linear elimination | $y = C(\psi;x)(1+\varepsilon)$<br>$\varepsilon \sim N(0,\sigma^2_{\text{prop}})$ | $\psi_i = \{Cl_i, V_{1i}, Q_i, V_{2i}\}$<br>$Cl_i = \theta_1(WT/70)^{0.75}\left(\frac{1}{1+\left(\frac{TM_{50}}{PMA}\right)^{Hill}}\right)\exp(\theta_{Cl,UGT1A4} I[UGT1A4 > 0] + \eta_{Cli})$<br>$V_{1i} = \theta_2(WT/70)\ \ \exp(\eta_{V1i})$<br>$Q_i = \theta_3(WT/70)^{0.75}\exp(\eta_{Qi})$<br>$V_{2i} = \theta_4(WT/70)\ \ \exp(\eta_{V2i})$<br>$\eta_{Cli} \sim N(0,\omega^2_{Cl});\ \eta_{V1i} \sim N(0,\omega^2_{V1})$<br>$\eta_{Qi} \sim N(0,\omega^2_{Q});\ \eta_{V2i} \sim N(0,\omega^2_{V2})$ | $\Omega = \begin{bmatrix} \omega^2_{Cl} & \omega_{Cl,V_1} & 0 & 0 \\ & \omega^2_{V_1} & 0 & 0 \\ & & \omega^2_Q & 0 \\ & & & \omega^2_{V_2} \end{bmatrix}$ | -1918.3 | -1832.4 |
| 29 | Two compartment multidose IV infusion/bolus with linear elimination | $y = C(\psi;x)(1+\varepsilon)$<br>$\varepsilon \sim N(0,\sigma^2_{\text{prop}})$ | $\psi_i = \{Cl_i, V_{1i}, Q_i, V_{2i}\}$<br>$Cl_i = \theta_1(WT/70)^{0.75}\left(\frac{1}{1+\left(\frac{TM_{50}}{PMA}\right)^{Hill}}\right)\exp(\theta_{Cl,UGT1A4} UGT1A4 + \eta_{Cli})$<br>$V_{1i} = \theta_2(WT/70)\ \ \exp(\eta_{V1i})$<br>$Q_i = \theta_3(WT/70)^{0.75}\exp(\eta_{Qi})$<br>$V_{2i} = \theta_4(WT/70)\ \ \exp(\eta_{V2i})$<br>$\eta_{Cli} \sim N(0,\omega^2_{Cl});\ \eta_{V1i} \sim N(0,\omega^2_{V1})$<br>$\eta_{Qi} \sim N(0,\omega^2_{Q});\ \eta_{V2i} \sim N(0,\omega^2_{V2})$ | $\Omega = \begin{bmatrix} \omega^2_{Cl} & \omega_{Cl,V_1} & 0 & 0 \\ & \omega^2_{V_1} & 0 & 0 \\ & & \omega^2_Q & 0 \\ & & & \omega^2_{V_2} \end{bmatrix}$ | -1917.6 | -1831.7 |

**Table S10.** Best covariate model and *CYP2A6* PRS score in subset with complete PRS data (n=350)

**30. Two compartment model with proportional error and fixed theory-based allometric scaling parameters and sigmoid (Hill) maturation, block correlation 2**

**31. Add exponential *CYP2A6* additive effect to Model 30**

| Model | Structural Model | Residual variability | Submodels | Between subject variability | OFV | BICc |
|---|---|---|---|---|---|---|
| 30 | Two compartment multidose IV infusion/bolus with linear elimination | $y = C(\psi; x)(1+\varepsilon)$<br>$\varepsilon \sim N(0, \sigma^2_{\text{prop}})$ | $\psi_i = \{Cl_i, V_{1i}, Q_i, V_{2i}\}$<br>$Cl_i = \theta_1(WT/70)^{0.75}\left(\frac{1}{1+\left(\frac{TM_{50}}{PMA}\right)^{Hill}}\right)\exp(\eta_{Cli})$<br>$V_{1i} = \theta_2(WT/70) \quad \exp(\eta_{V1i})$<br>$Q_i = \theta_3(WT/70)^{0.75}\exp(\eta_{Qi})$<br>$V_{2i} = \theta_4(WT/70) \quad \exp(\eta_{V2i})$<br>$\eta_{Cli} \sim N(0, \omega^2_{Cl});\ \eta_{V1i} \sim N(0, \omega^2_{V1})$<br>$\eta_{Qi} \sim N(0, \omega^2_{Q});\ \eta_{V2i} \sim N(0, \omega^2_{V2})$ | $\Omega = \begin{bmatrix} \omega^2_{Cl} & \omega_{Cl,V_1} & 0 & 0 \\ & \omega^2_{V_1} & 0 & 0 \\ & & \omega^2_Q & 0 \\ & & & \omega^2_{V_2} \end{bmatrix}$ | -1889.2 | -1809.3 |
| 31 | Two compartment multidose IV infusion/bolus with linear elimination | $y = C(\psi; x)(1+\varepsilon)$<br>$\varepsilon \sim N(0, \sigma^2_{\text{prop}})$ | $\psi_i = \{Cl_i, V_{1i}, Q_i, V_{2i}\}$<br>$Cl_i = \theta_1(WT/70)^{0.75}\left(\frac{1}{1+\left(\frac{TM_{50}}{PMA}\right)^{Hill}}\right)\exp(\theta_{Cl,CYP2A6}(CYP2A6score)+\eta_{Cli})$<br>$V_{1i} = \theta_2(WT/70) \quad \exp(\eta_{V1i})$<br>$Q_i = \theta_3(WT/70)^{0.75}\exp(\eta_{Qi})$<br>$V_{2i} = \theta_4(WT/70) \quad \exp(\eta_{V2i})$<br>$\eta_{Cli} \sim N(0, \omega^2_{Cl});\ \eta_{V1i} \sim N(0, \omega^2_{V1})$<br>$\eta_{Qi} \sim N(0, \omega^2_{Q});\ \eta_{V2i} \sim N(0, \omega^2_{V2})$ | $\Omega = \begin{bmatrix} \omega^2_{Cl} & \omega_{Cl,V_1} & 0 & 0 \\ & \omega^2_{V_1} & 0 & 0 \\ & & \omega^2_Q & 0 \\ & & & \omega^2_{V_2} \end{bmatrix}$ | -1889.9 | -1804.2 |

Abbreviations for Tables **S3 – S10**: IV, intravenous; C, concentration; $\sigma_{prop}$ and $\sigma_{add}$ are proportional and additive residual error terms; OFV, objective function value; BICc, corrected Bayesian information criteria; Cl, total clearance (L/hr); V, volume of distribution for the central compartment (L) in one compartment model; Q, intercompartmental clearance (L/hr); $V_1$, volume of distribution for the central compartment (L) in two compartment model; $V_2$, volume of distribution for the peripheral compartment (L); $\omega_{Cl}$, $\omega_{V1}$, $\omega_{Q}$, $\omega_{V2}$, the standard deviation for $\eta_i^{CL}$ ,$\eta_i^{V1}$, $\eta_i^{Q}$, and $\eta_i^{V2}$, respectively; $\Omega$ is intra-individual variance-covariance matrix; $TM_{50}$ postmenstrual age at which clearance is 50% of adult value; Hill, maturation factor slope coefficient; CV, coefficient of variation; WT, body weight in kg; PMA, postmenstrual age in weeks

**Supplemental Figures**

**Figure S1.** Distribution of (A) postnatal age and (B) postmenstrual age in final study population.

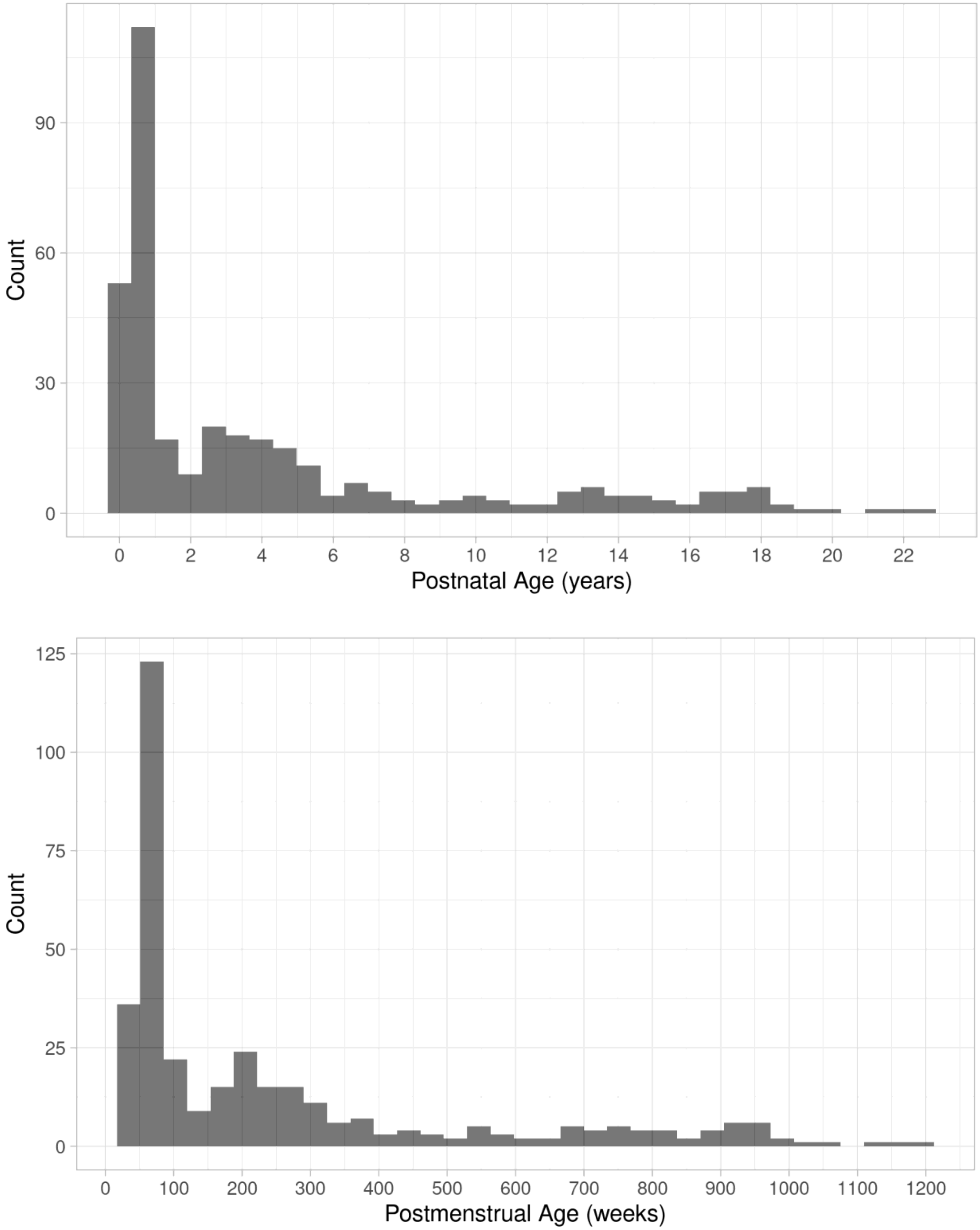

**Figure S2.** Distribution of weight in final study population.

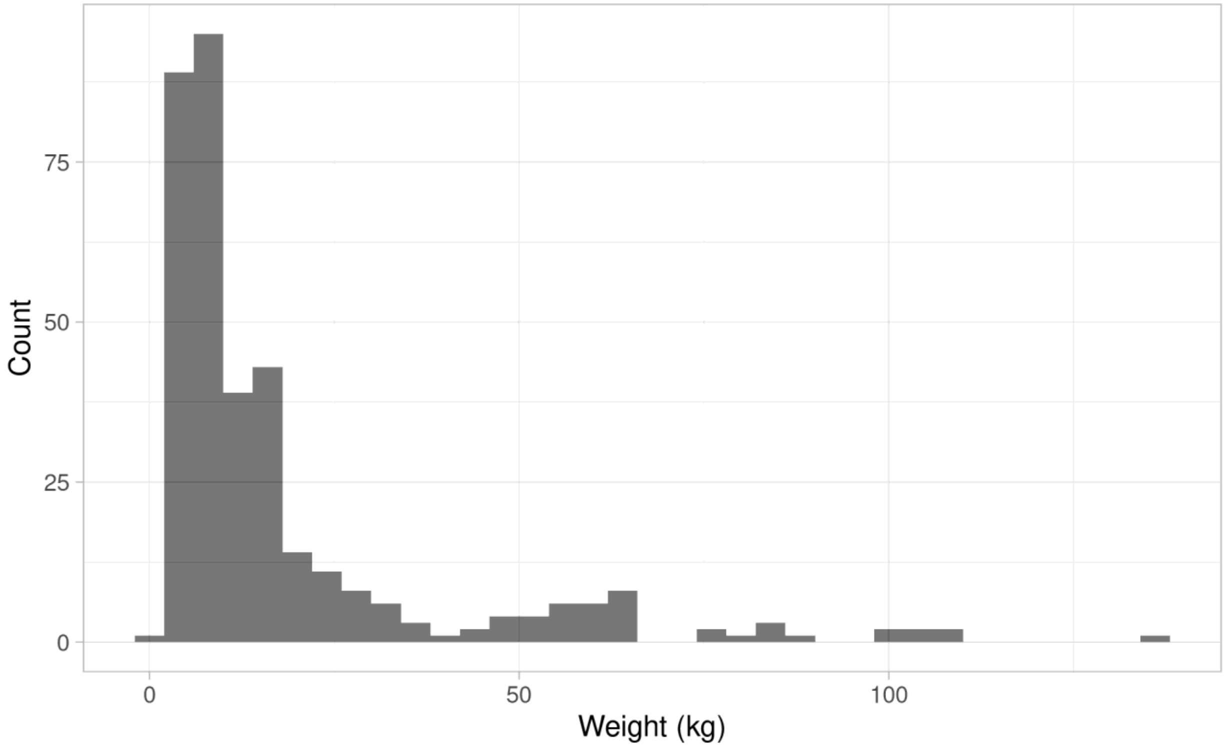


### Model 2 graphical goodness-of-fit checks

**Figure S3.** (A) Observed vs. population predicted concentrations and (B) observed vs. individual predicted concentrations.

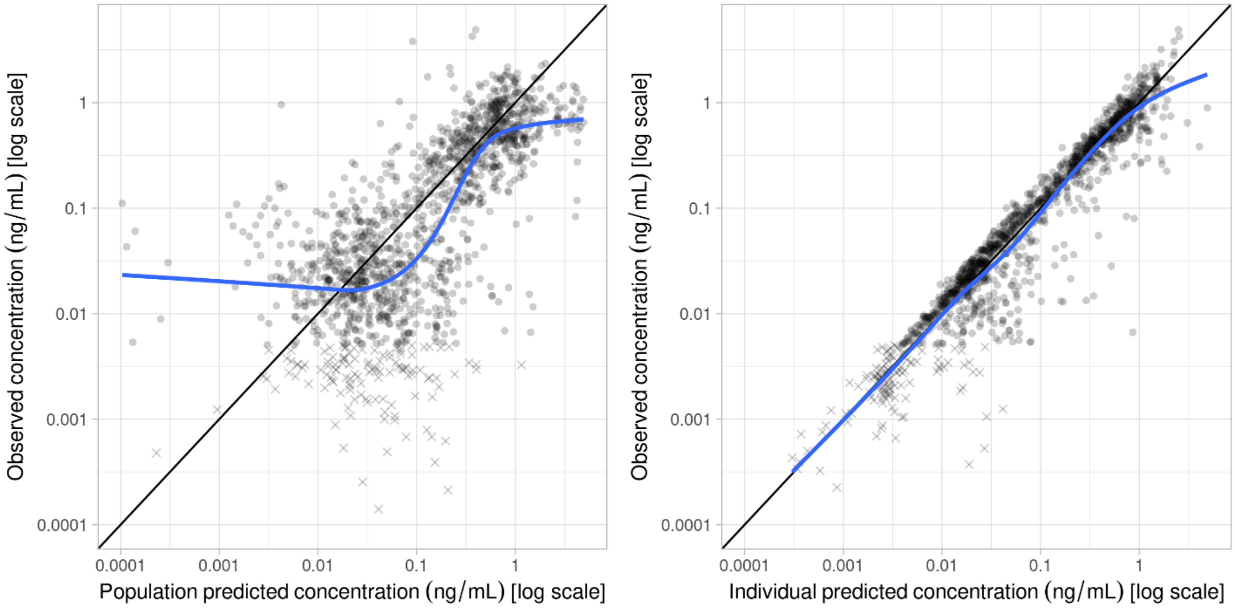

**Figure S4.** (A) Individual weighted residuals vs. predicted concentration and (B) individual weighted residuals vs. time.

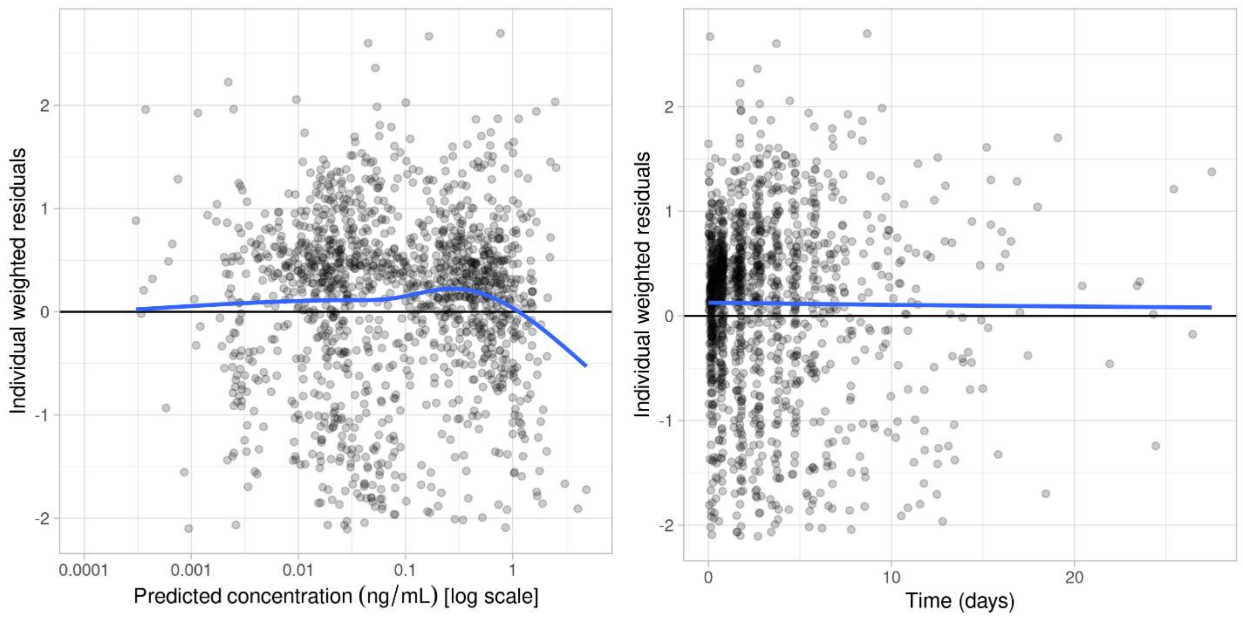


**Figure S5.** (A) Random effects correlations and (B) decorrelated random effects correlations.

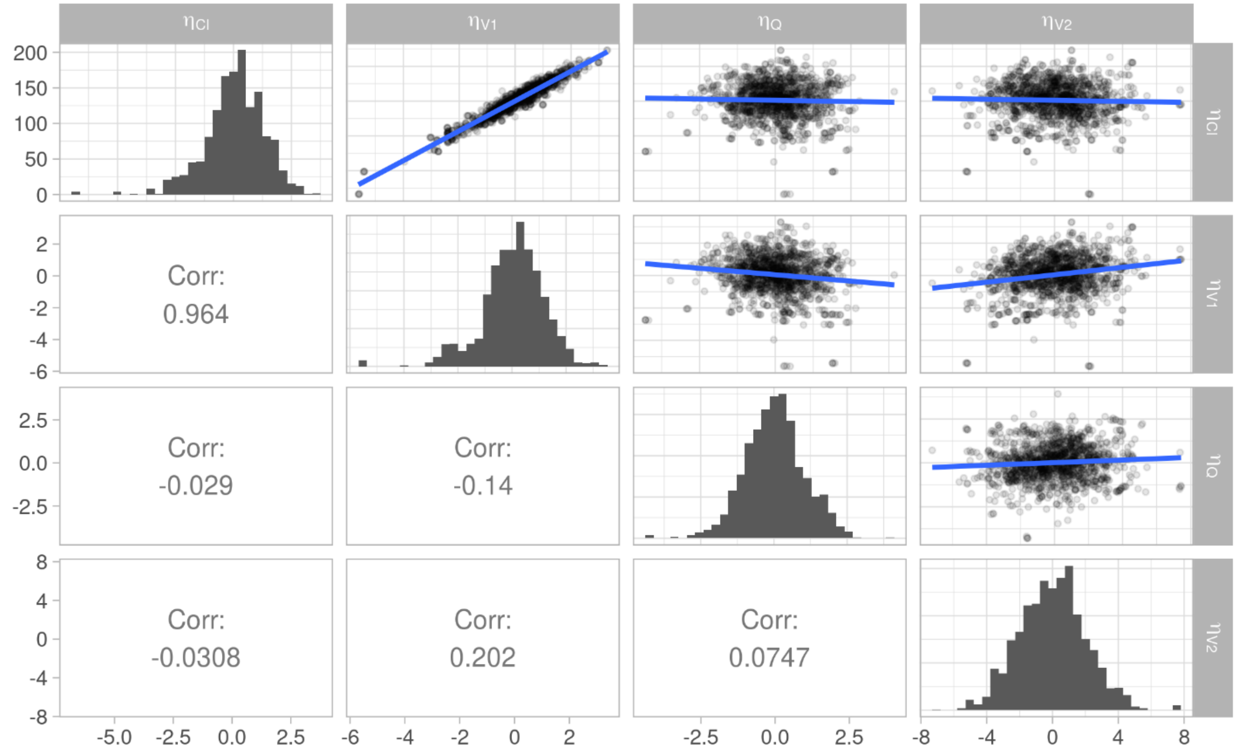

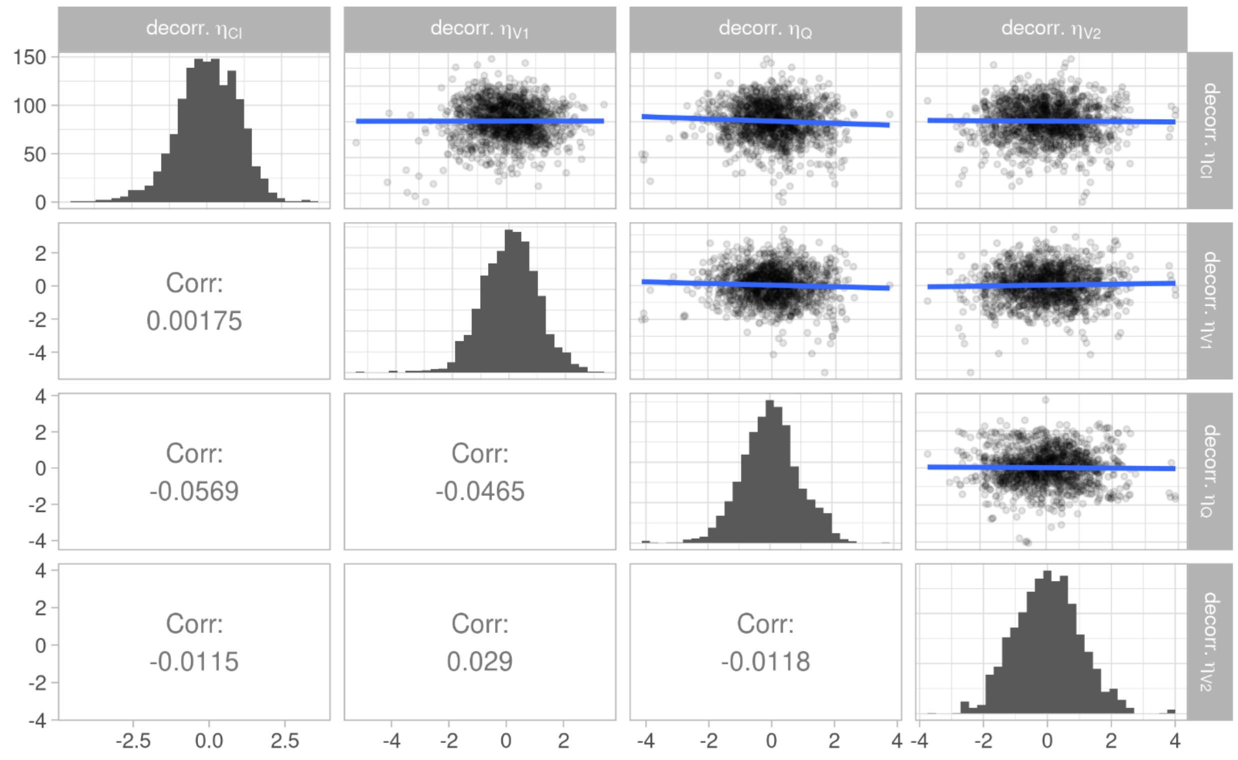


**Figure S6.** (A) Random effects vs. continuous covariates and (B) random effects vs. categorical covariates.

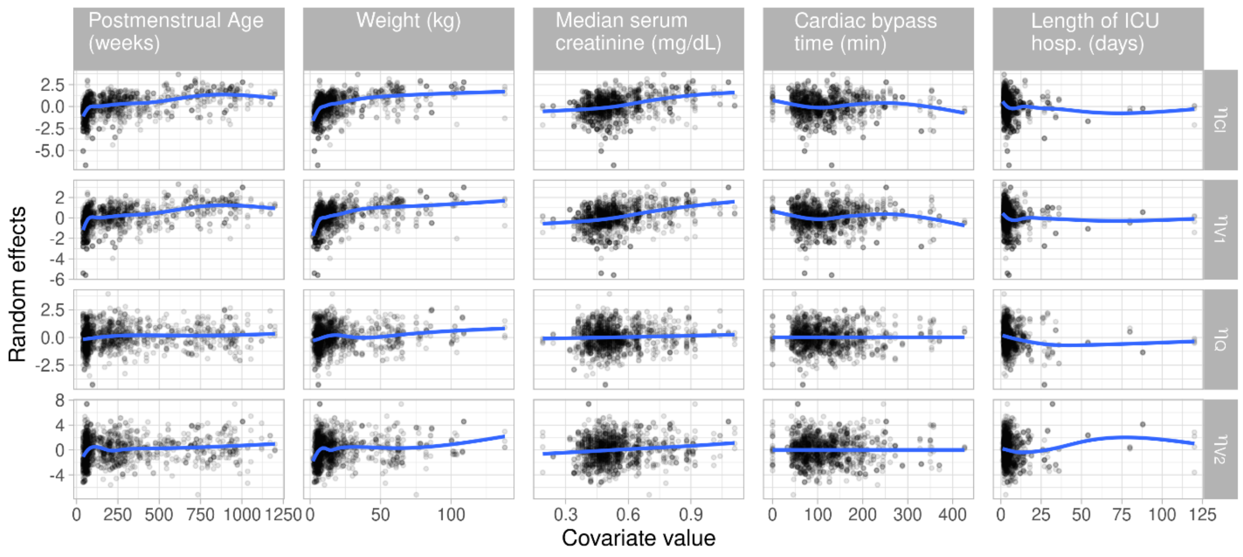

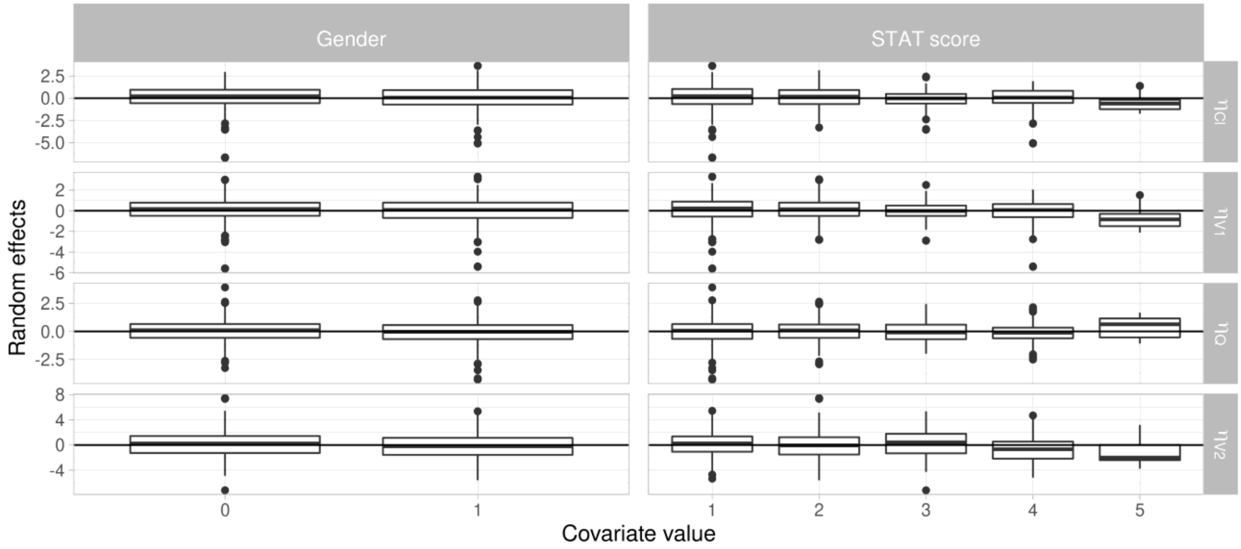


**Figure S7.** Prediction corrected visual predictive check.

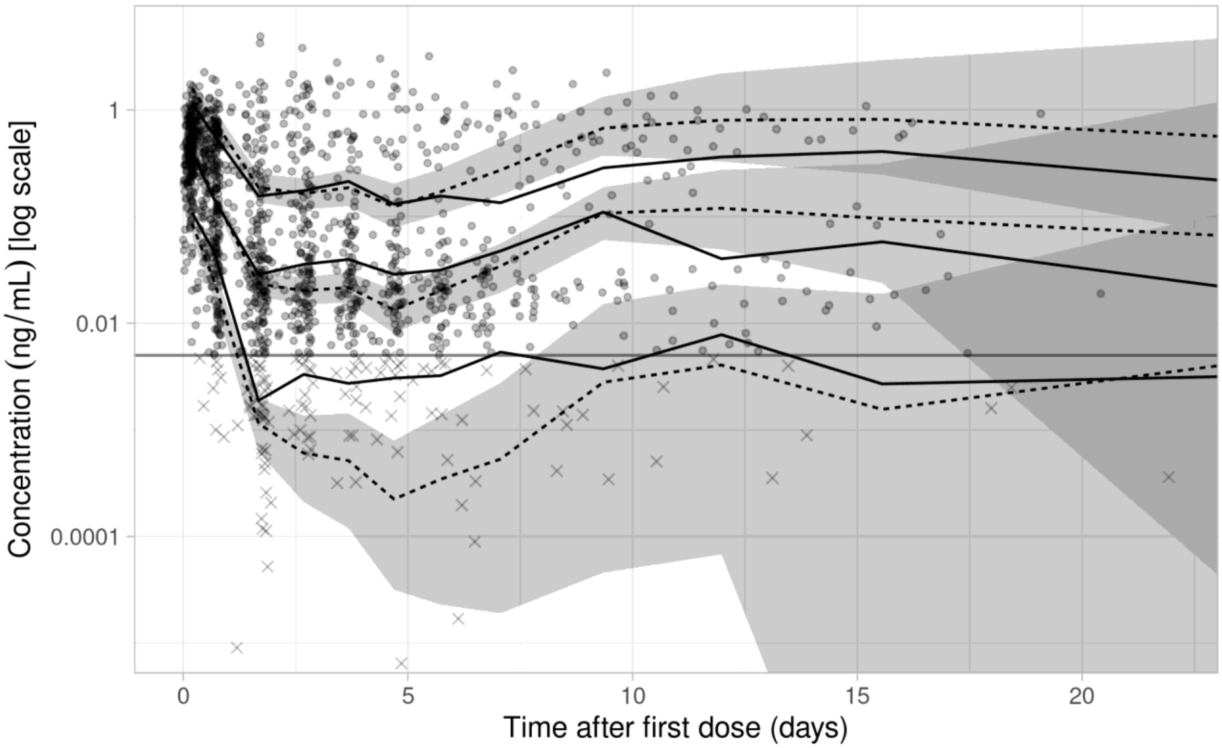

**Model 5 graphical goodness-of-fit checks**

**Figure S8.** (A) Observed vs. population predicted concentrations and (B) observed vs. individual predicted concentrations.

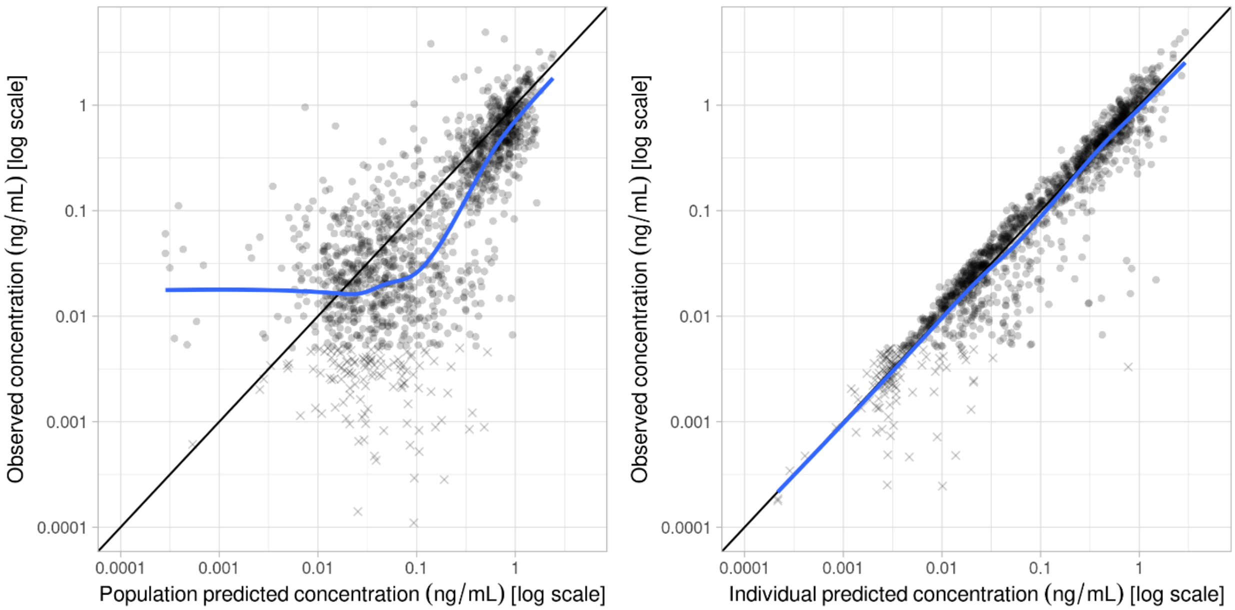


**Figure S9.** (A) Individual weighted residuals vs. predicted concentration and (B) individual weighted residuals vs. time.

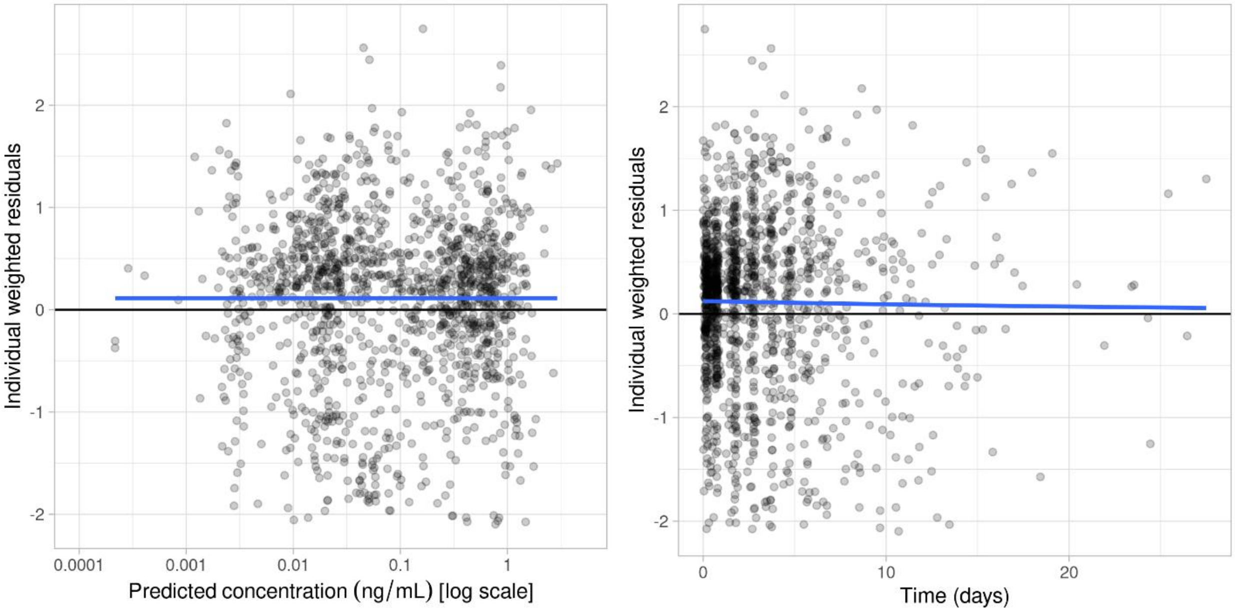

**Figure S10.** (A) Random effects correlations and (B) decorrelated random effects correlations.

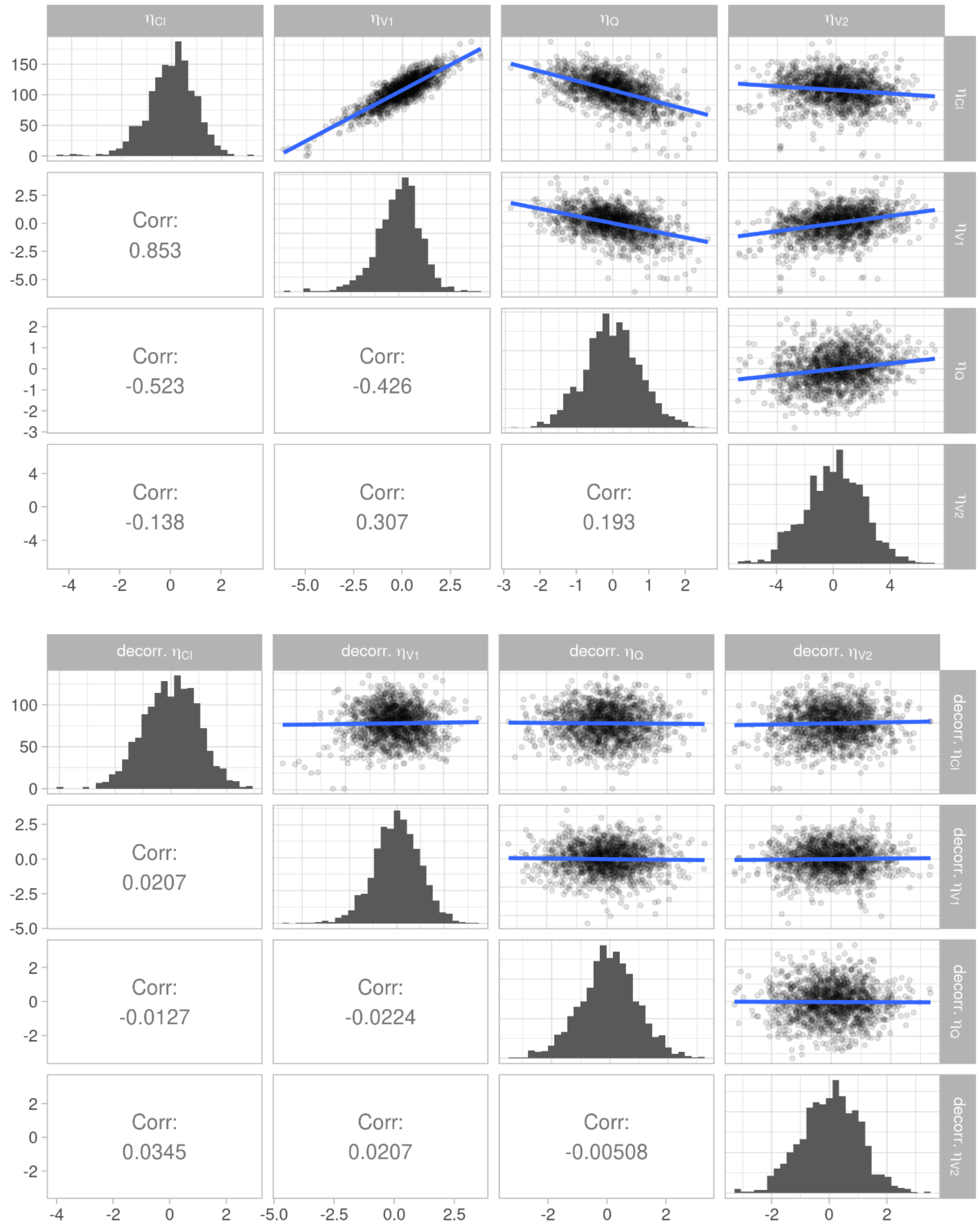

**Figure S11.** (A) Random effects vs. continuous covariates and (B) random effects vs. categorical covariates.

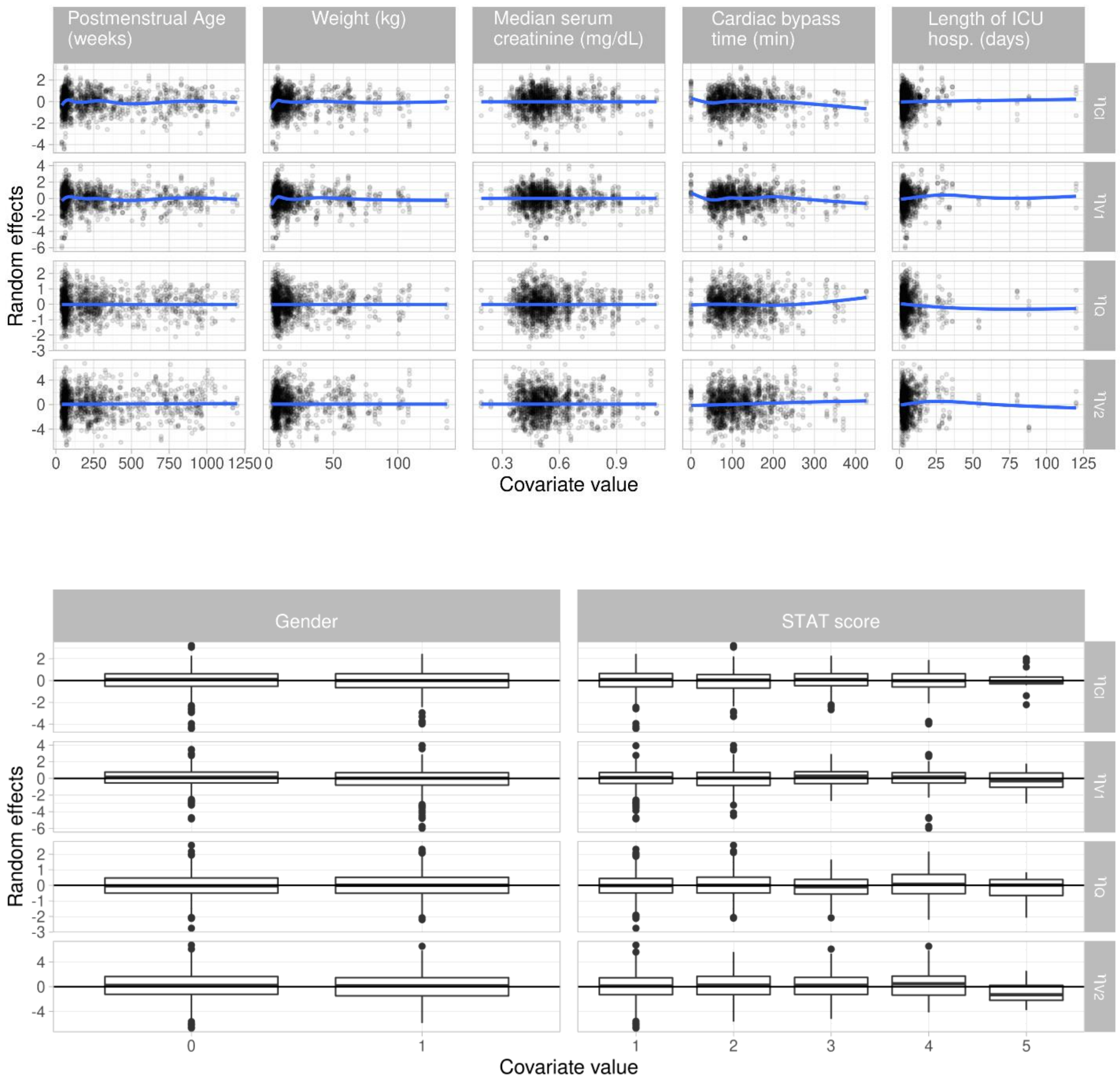

**Figure S12.** Prediction corrected visual predictive check.

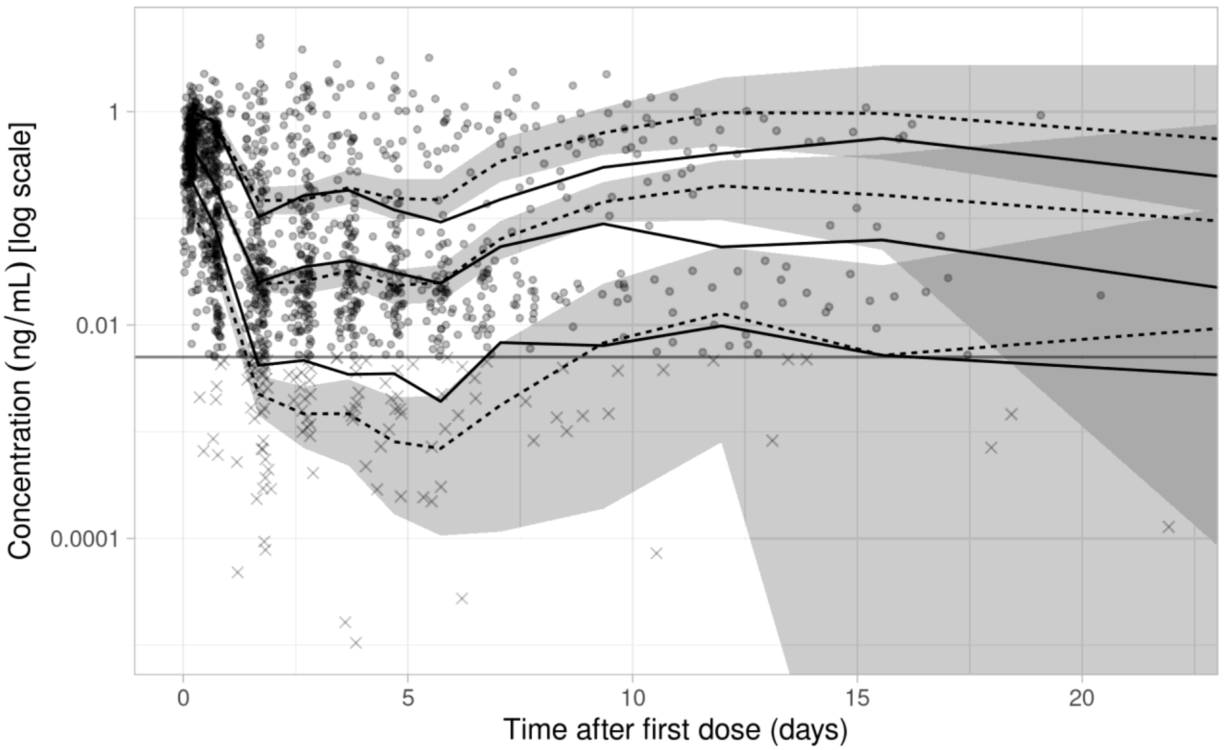


### Model 8 graphical goodness-of-fit checks

**Figure S13.** (A) Observed vs. population predicted concentrations and (B) observed vs. individual predicted concentrations.

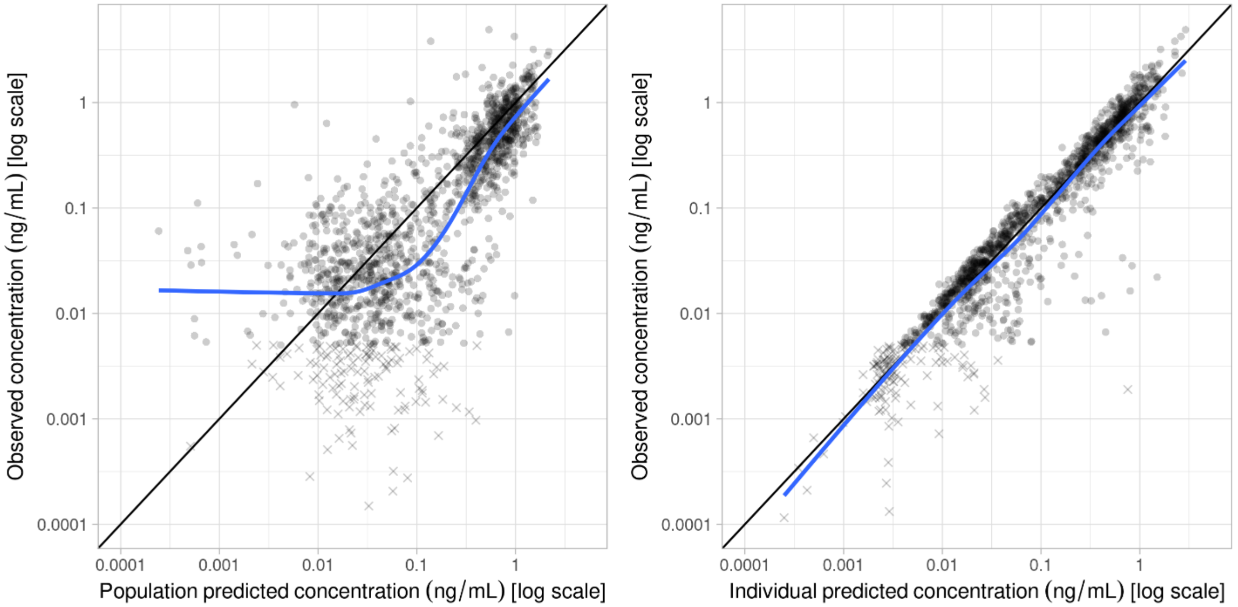

**Figure S14.** (A) Individual weighted residuals vs. predicted concentration and (B) individual weighted residuals vs. time.

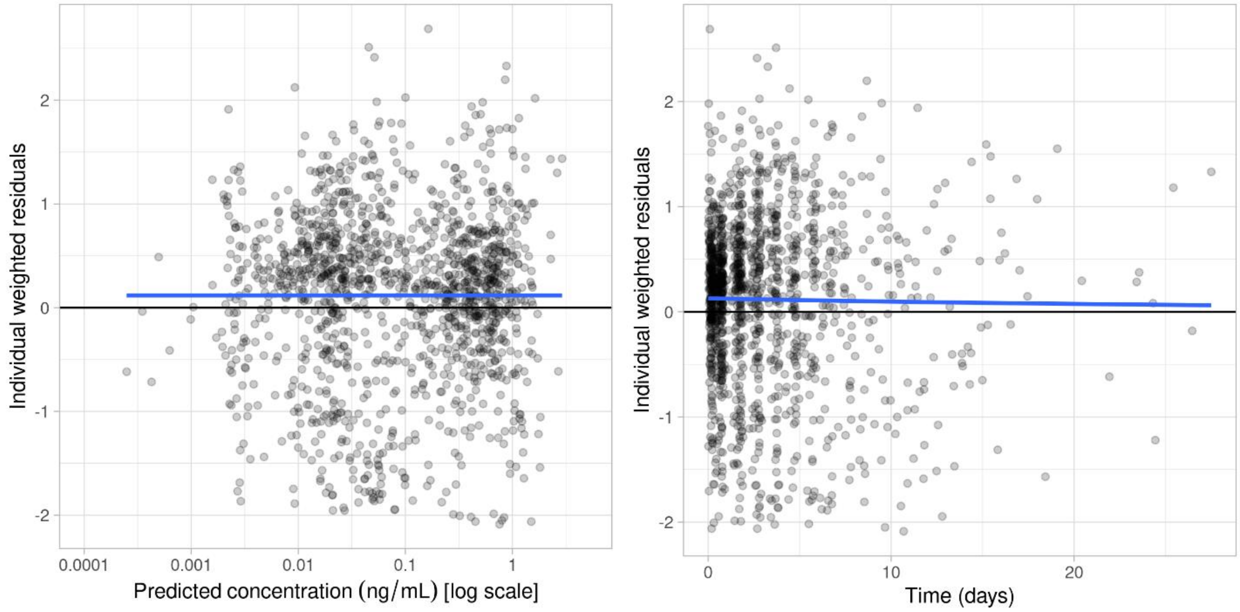


**Figure S15.** (A) Random effects correlations and (B) decorrelated random effects correlations.

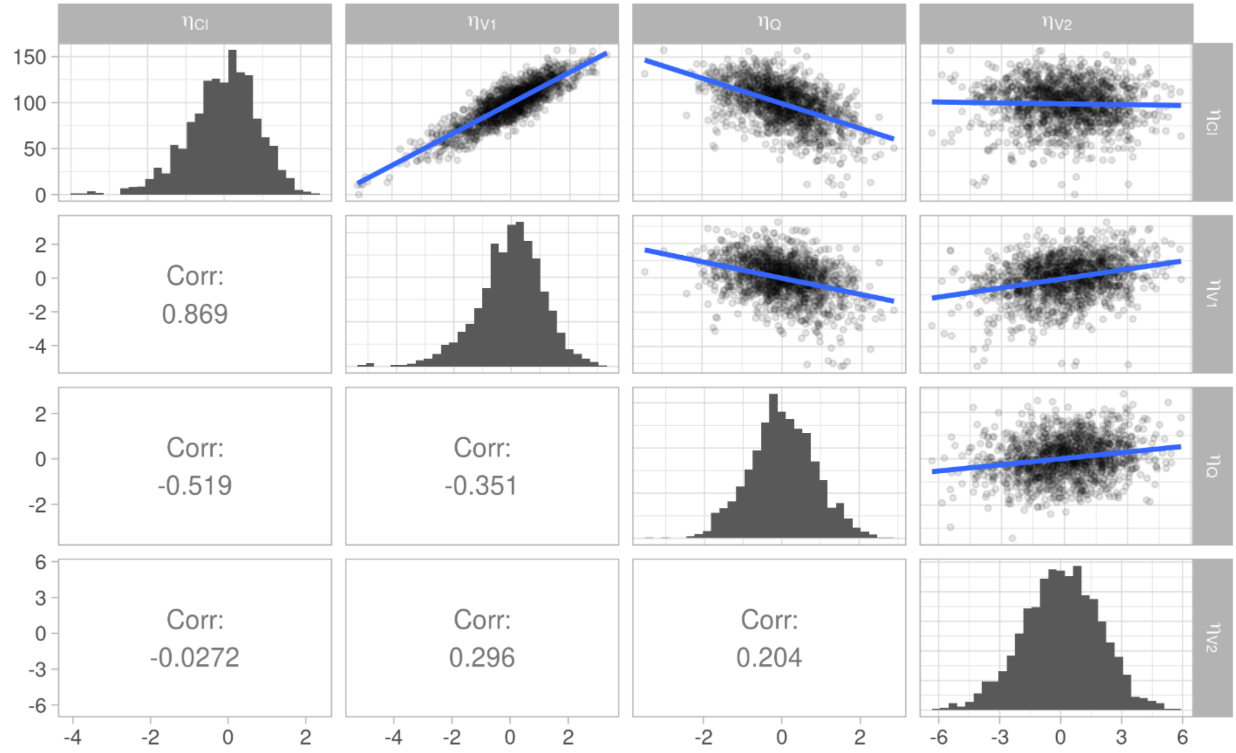

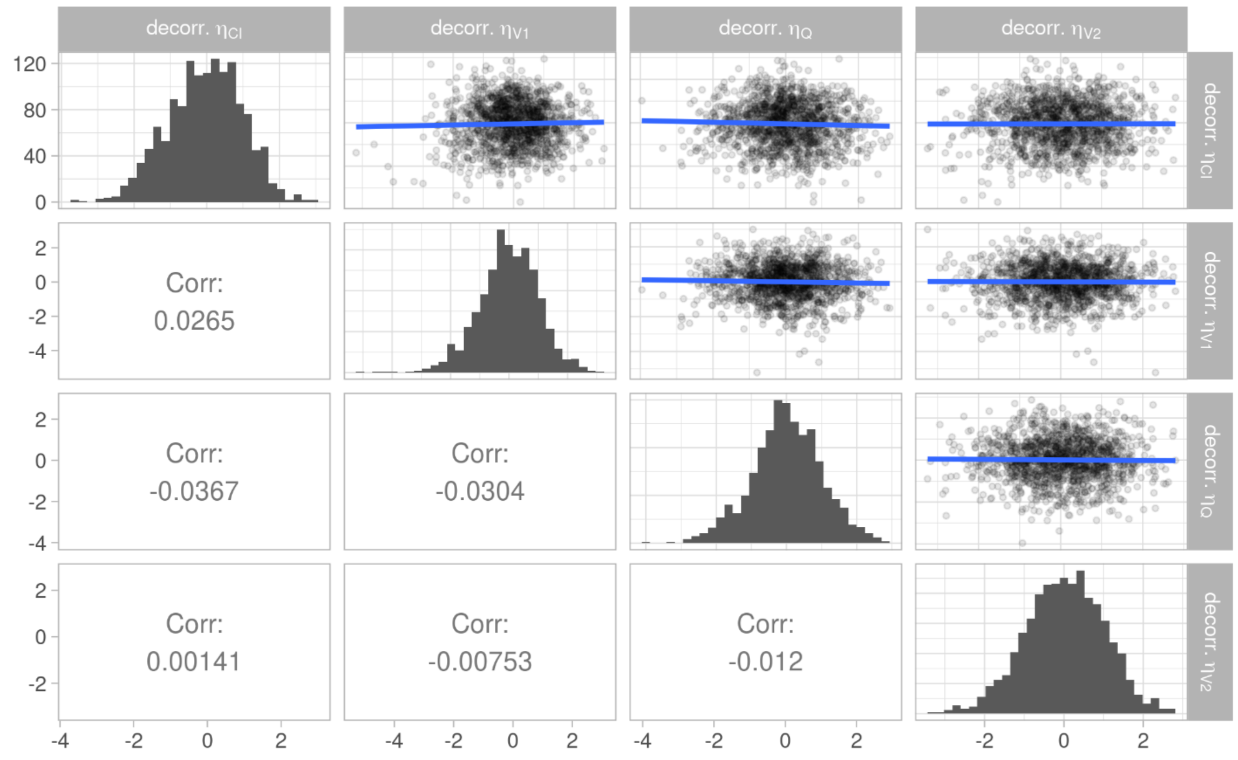


**Figure S16.** (A) Random effects vs. continuous covariates and (B) random effects vs. categorical covariates.

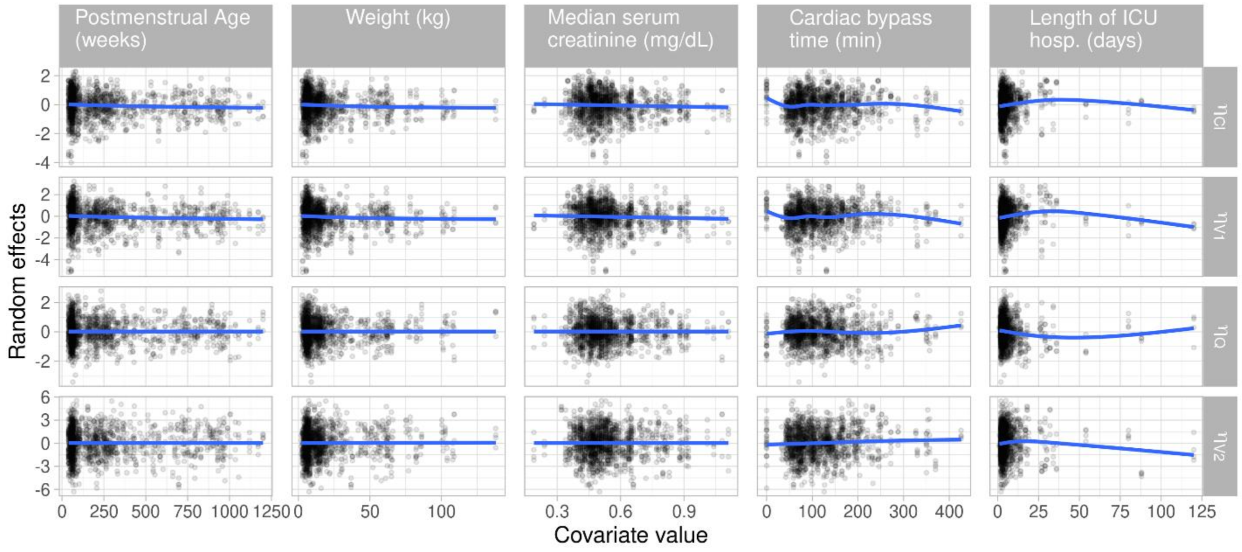

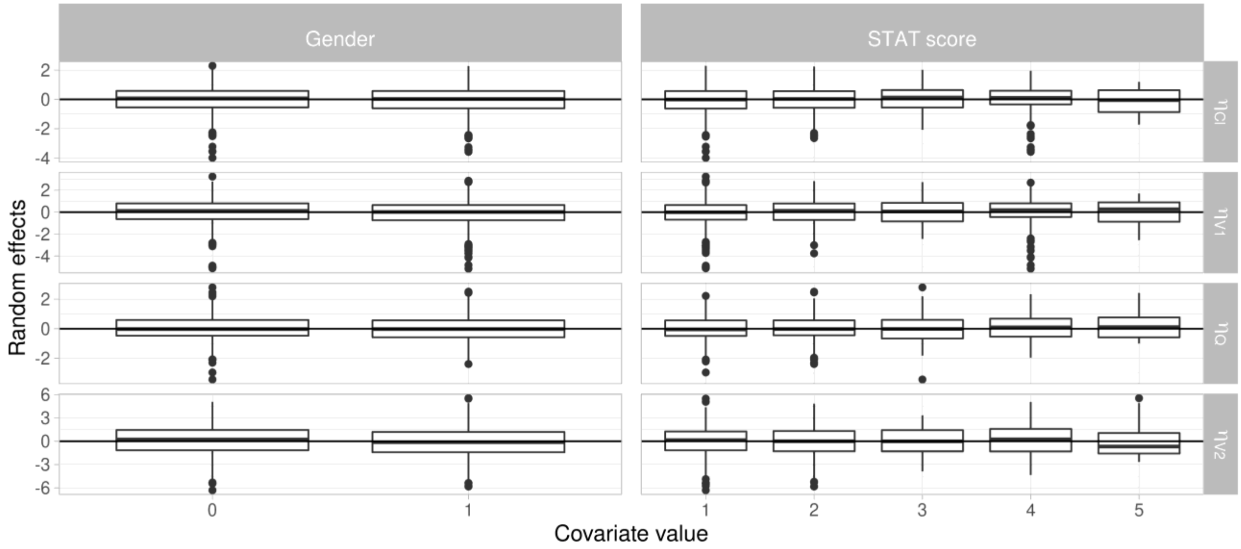


**Figure S17.** Prediction corrected visual predictive check.

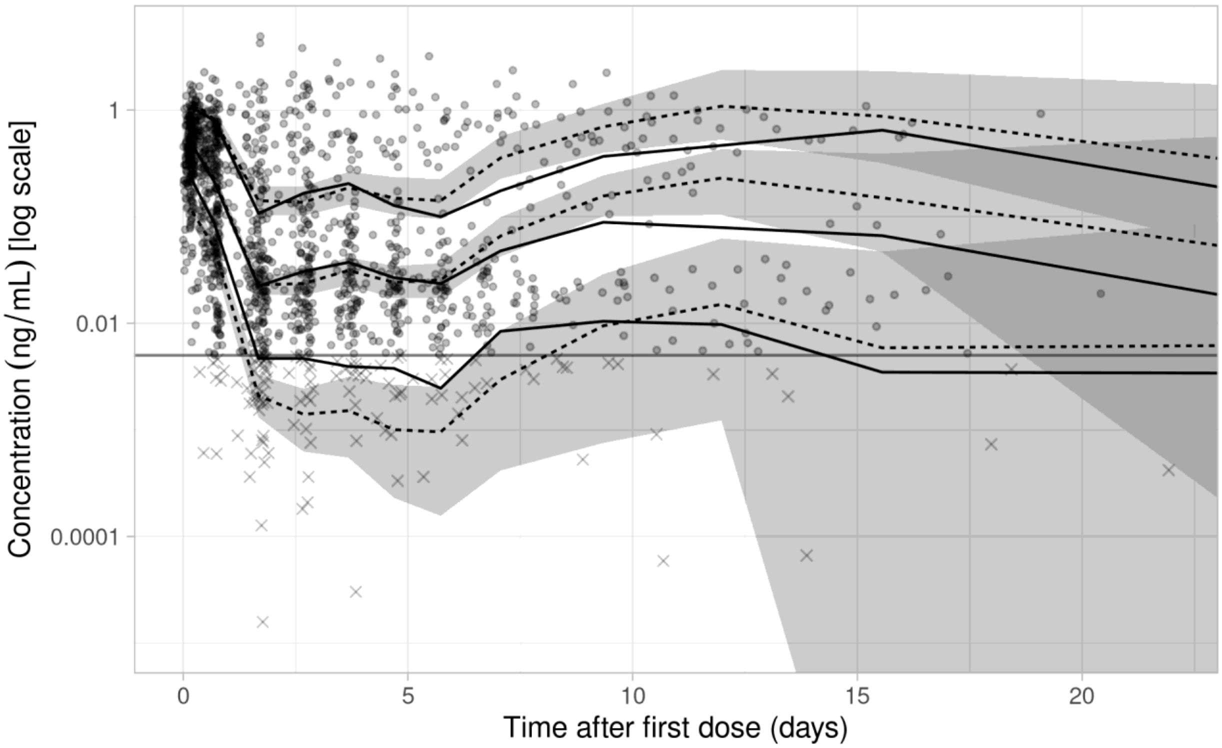

### Model 13 graphical goodness-of-fit checks

**Figure S18.** (A) Observed vs. population predicted concentrations and (B) observed vs. individual predicted concentrations.

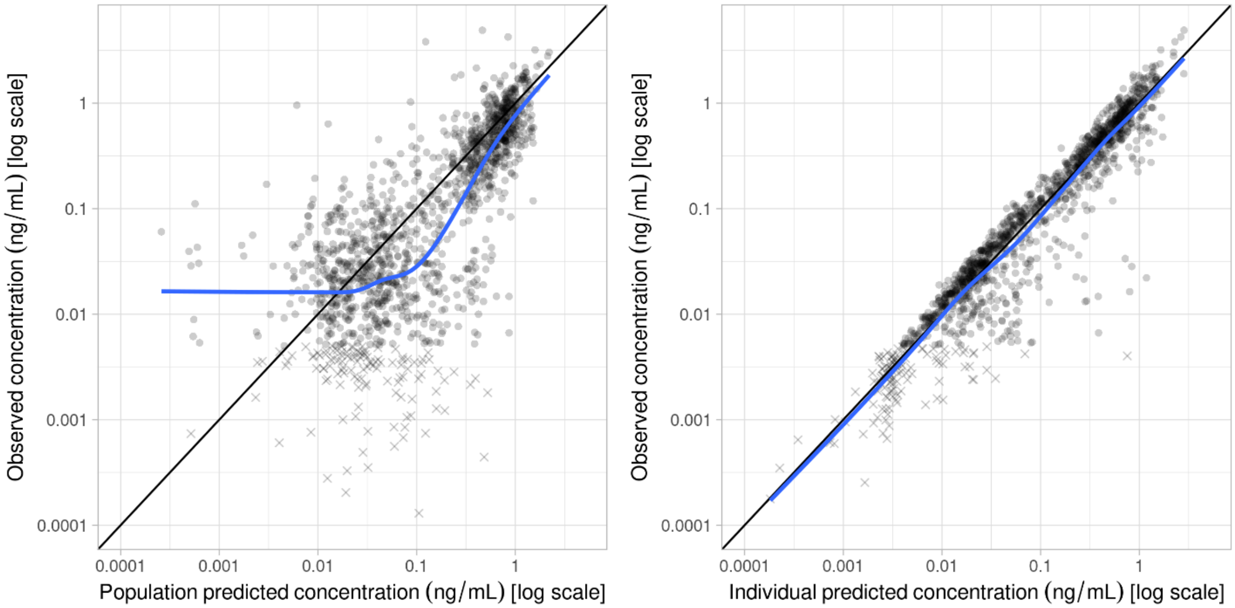


**Figure S19.** (A) Individual weighted residuals vs. predicted concentration and (B) individual weighted residuals vs. time.

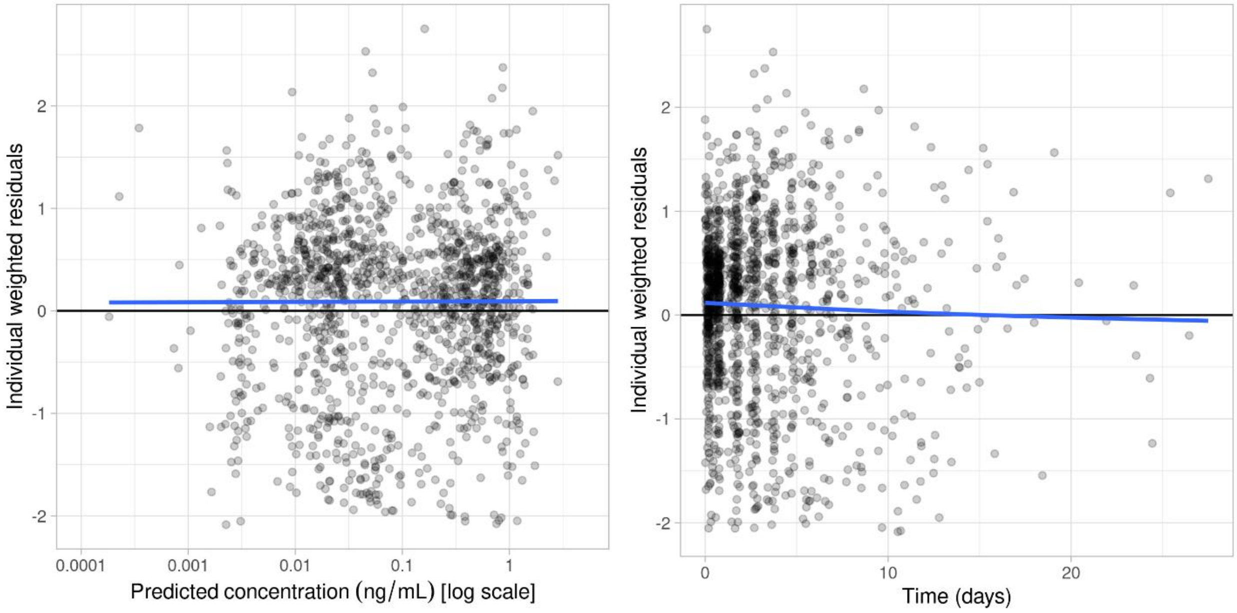

**Figure S20.** (A) Random effects correlations and (B) decorrelated random effects correlations.

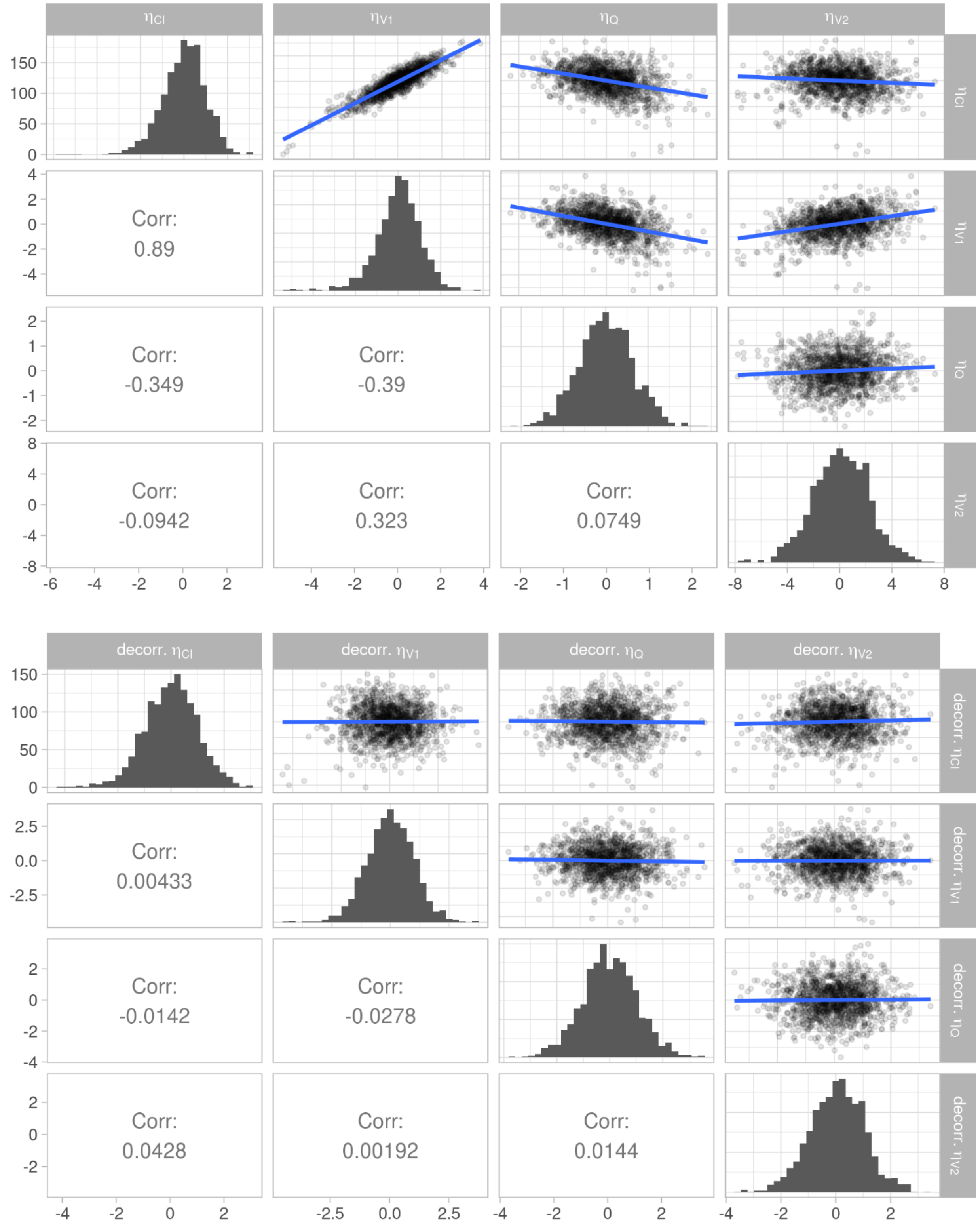

**Figure S21.** (A) Random effects vs. continuous covariates and (B) random effects vs. categorical covariates.

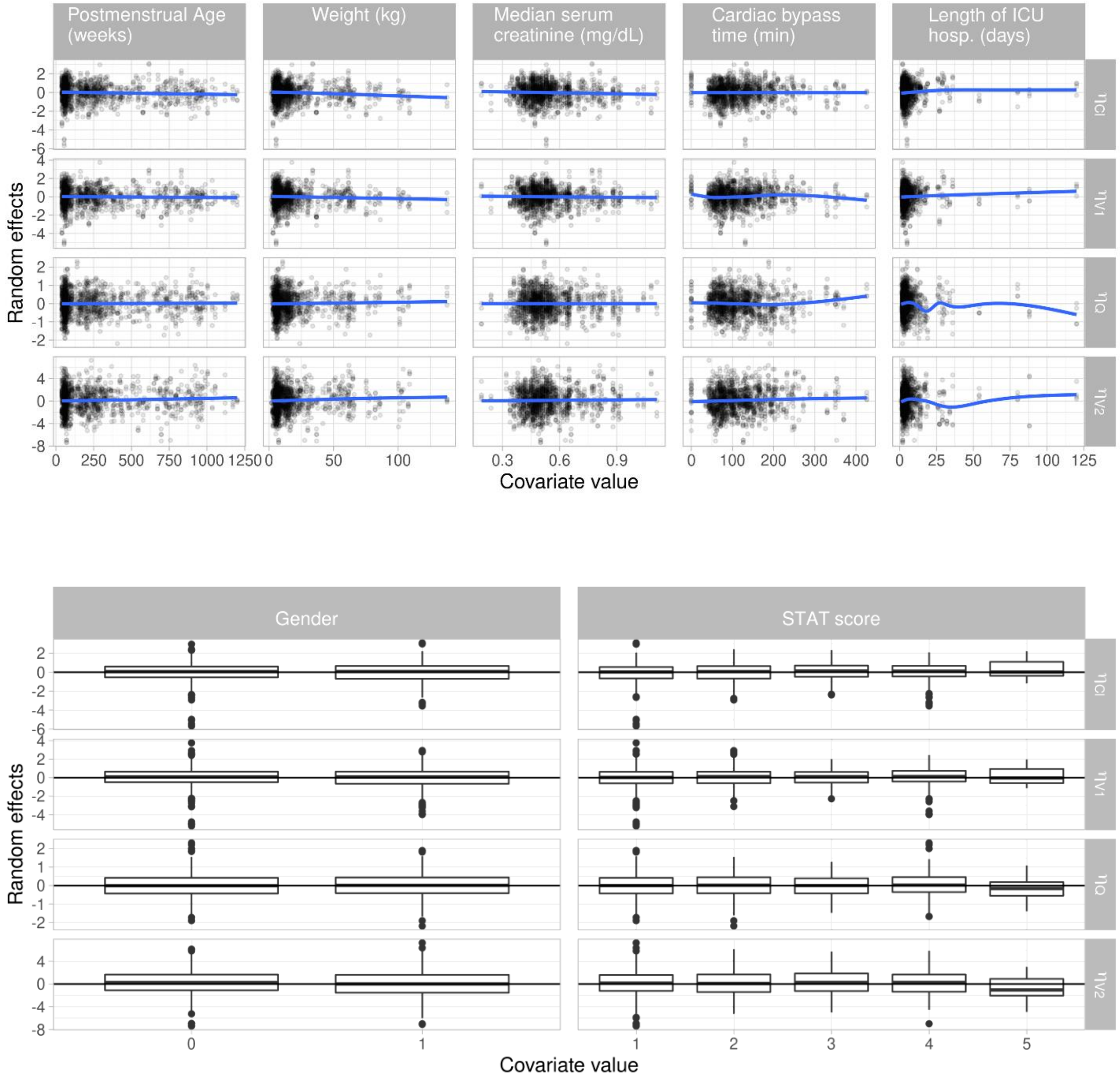

**Figure S22.** Prediction corrected visual predictive check.

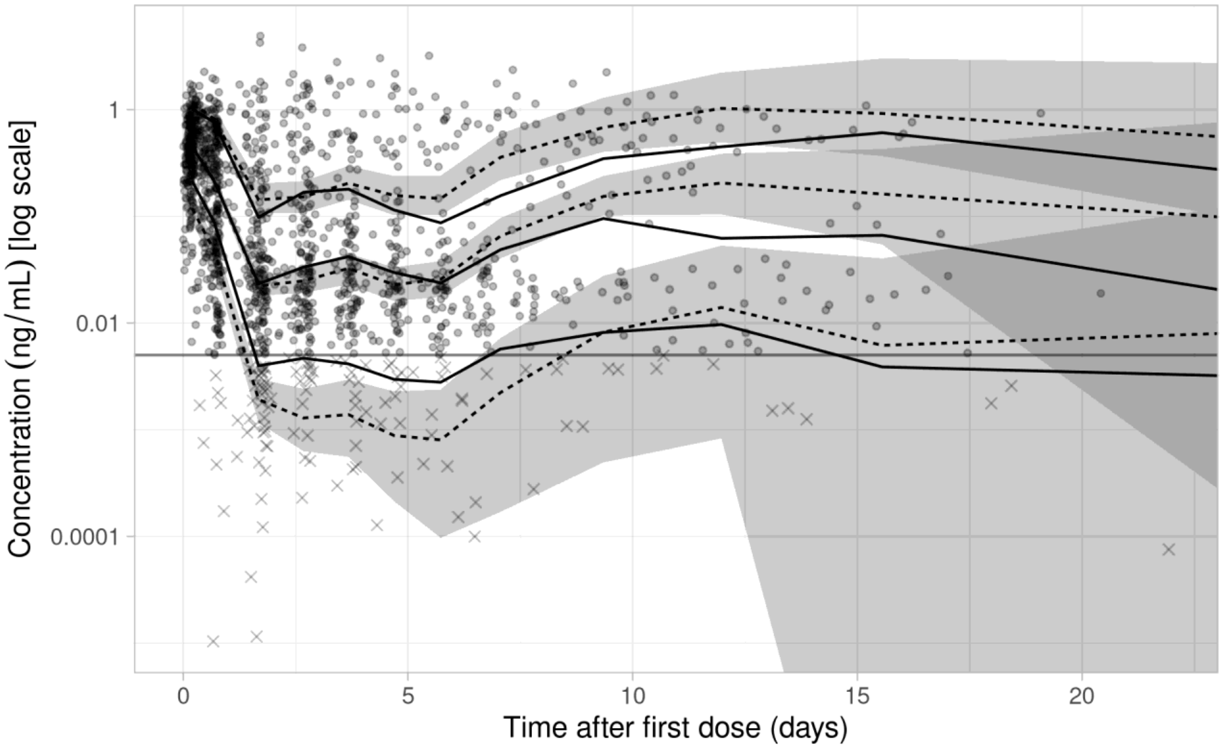


### Model 15 graphical goodness-of-fit checks

**Figure S23.** (A) Observed vs. population predicted concentrations and (B) observed vs. individual predicted concentrations.

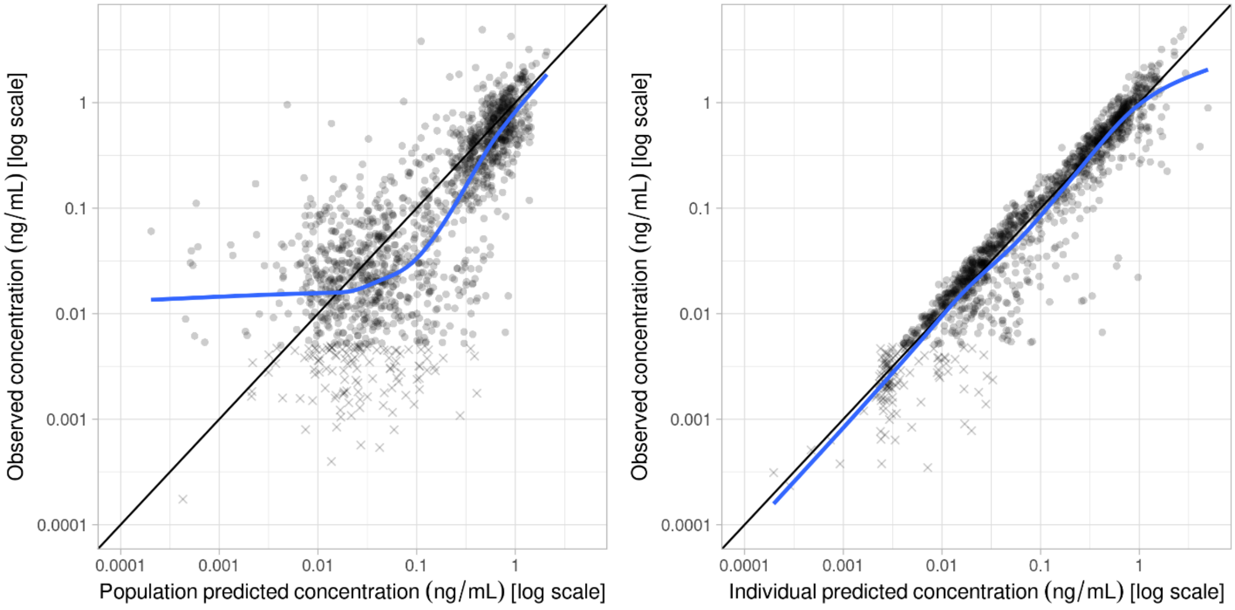

**Figure S24.** (A) Individual weighted residuals vs. predicted concentration and (B) individual weighted residuals vs. time.

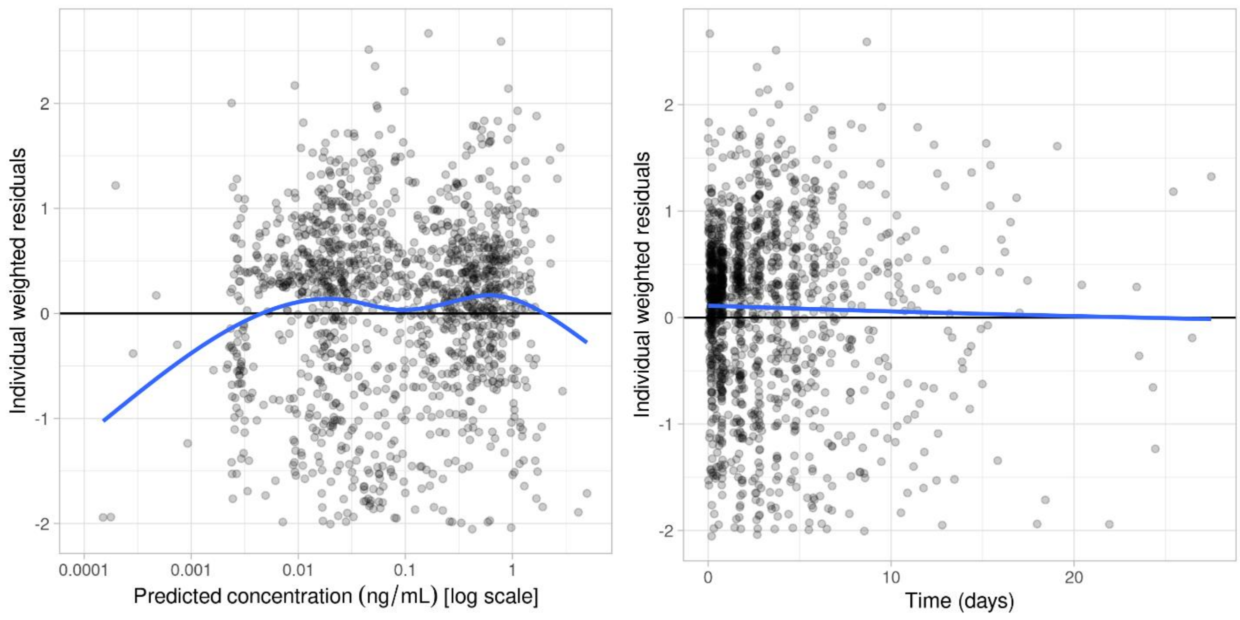


**Figure S25.** (A) Random effects correlations and (B) decorrelated random effects correlations.

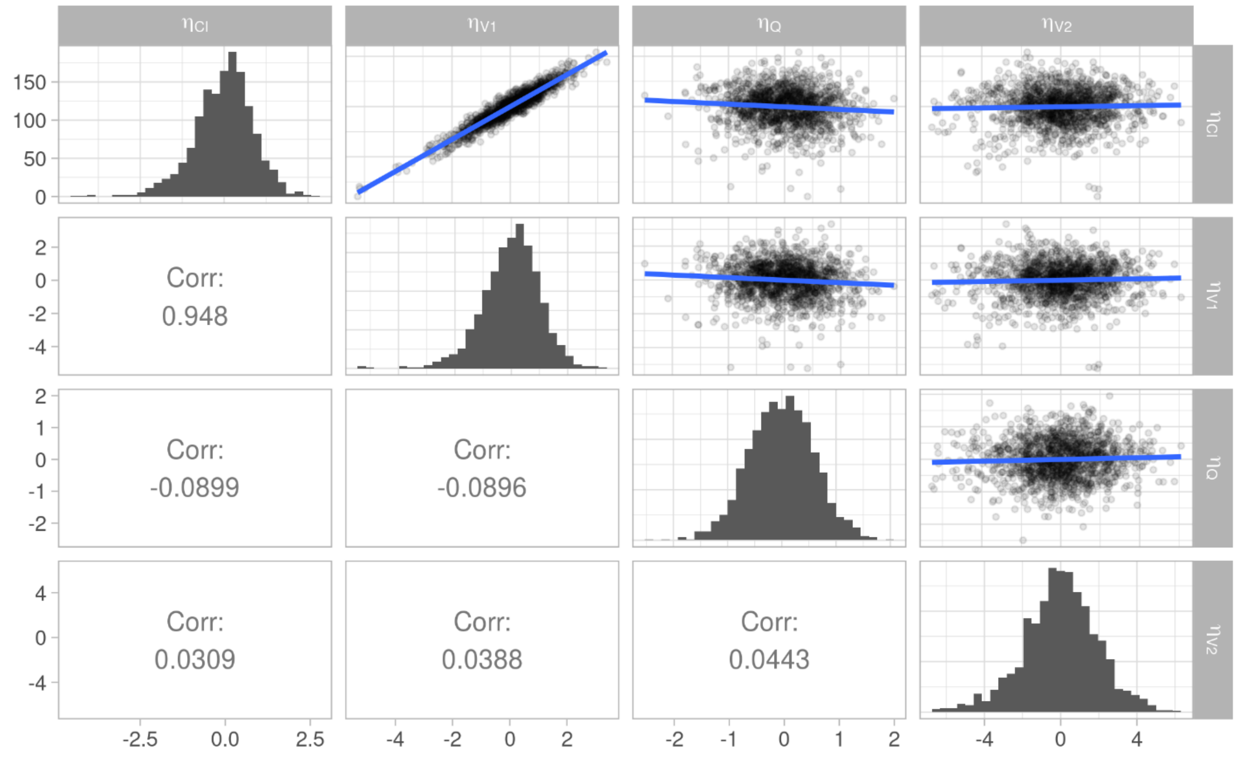

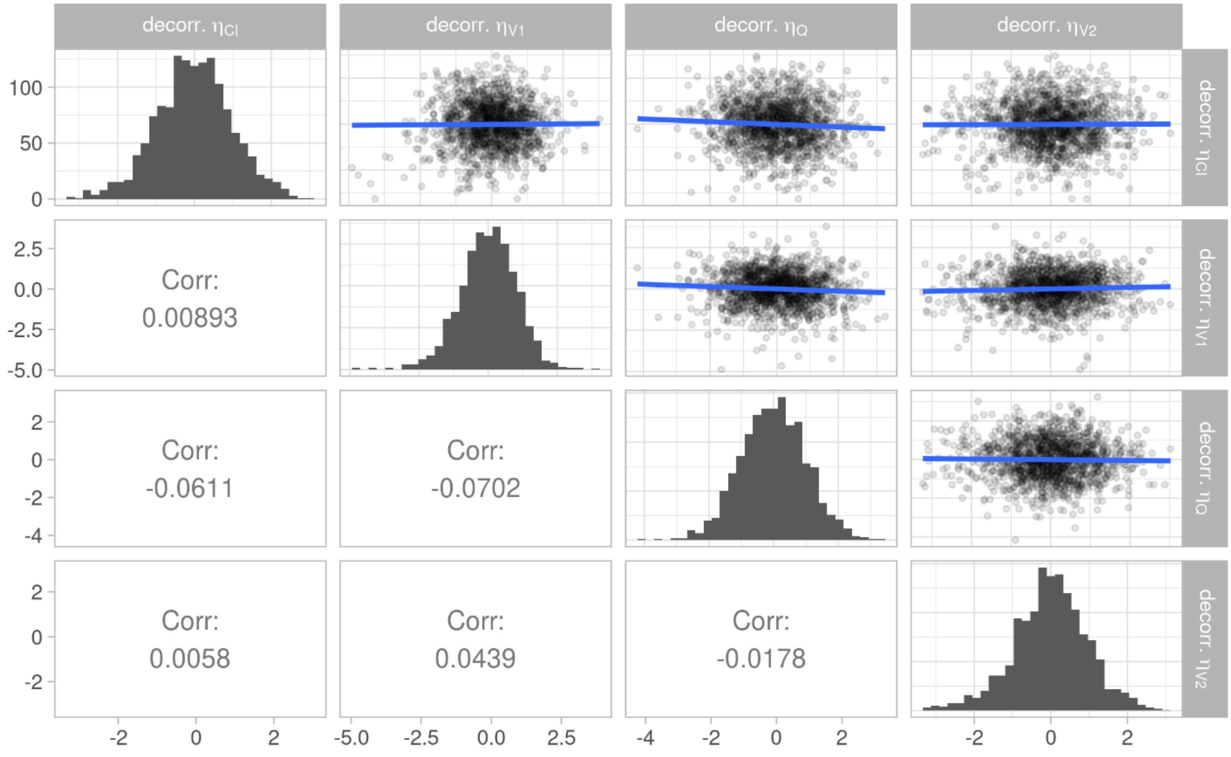


**Figure S26.** (A) Random effects vs. continuous covariates and (B) random effects vs. categorical covariates.

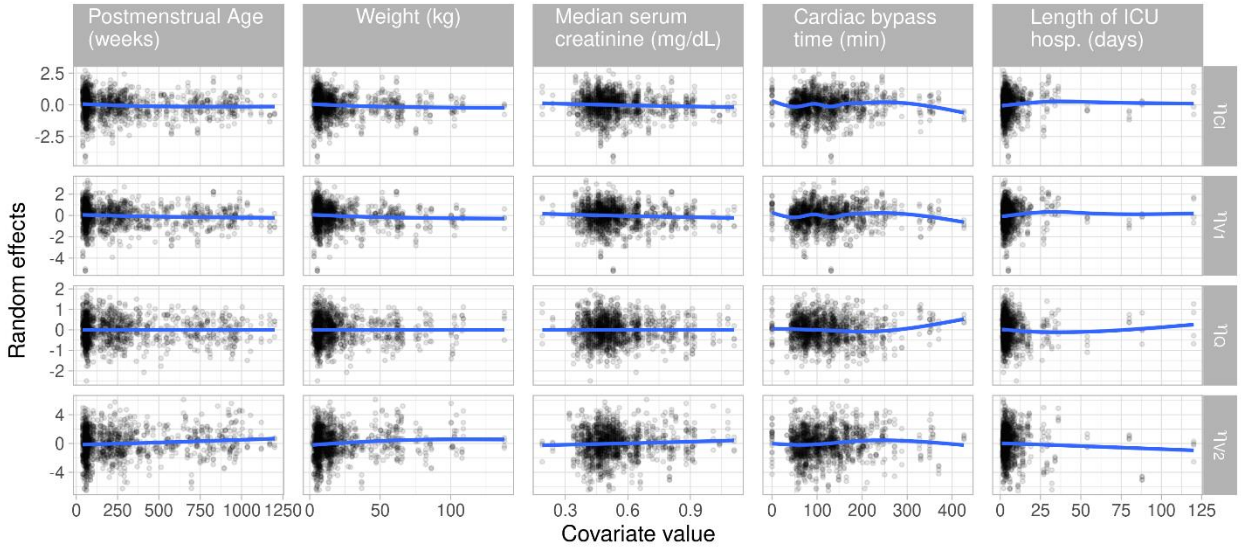

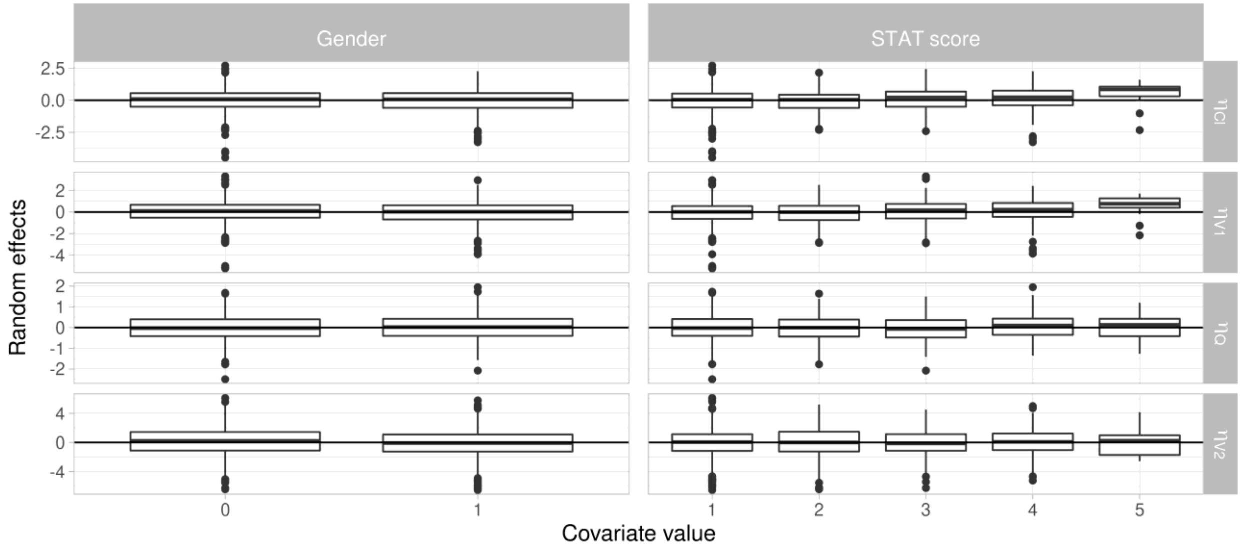


**Figure S27.** Prediction corrected visual predictive check.

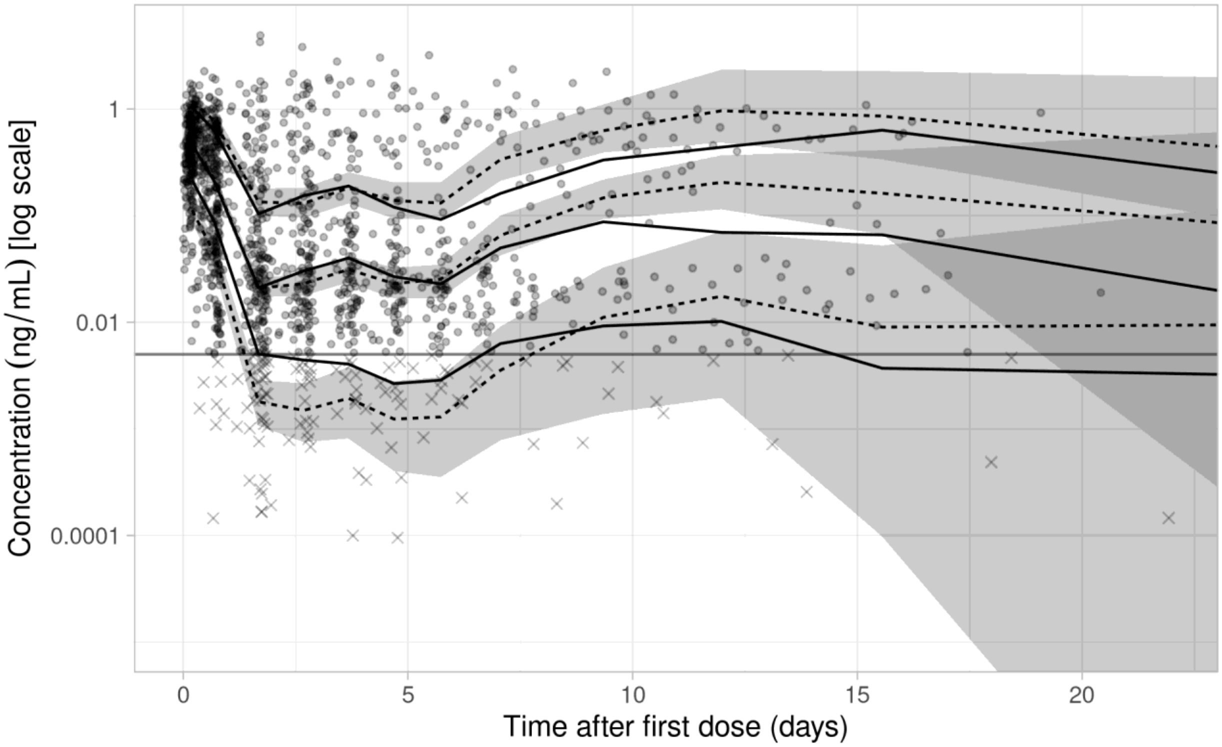

**Figure S28.** Prediction corrected visual predictive check for final model (Model 16) stratified by postnatal age group.

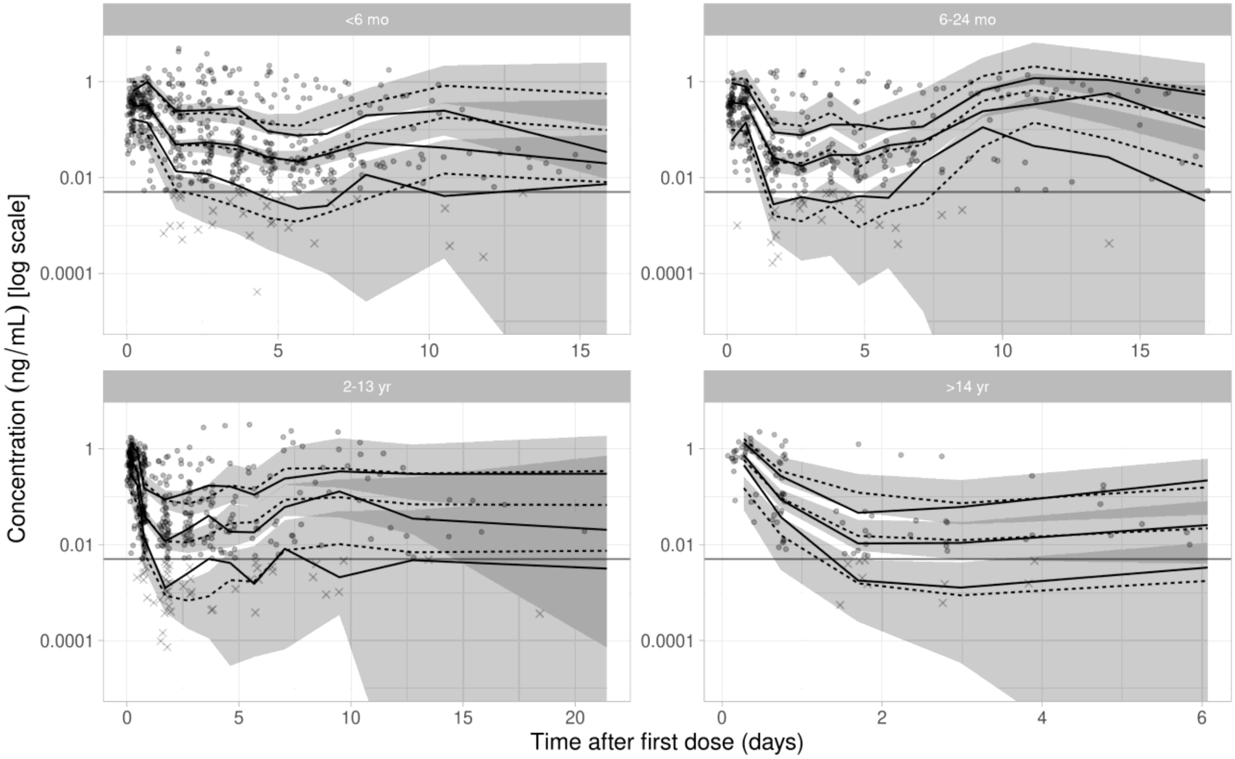

### Genetic Effects on Clearance and Concentration

**Figure S29.** Model estimated clearance by *UGT1A4* genotype.

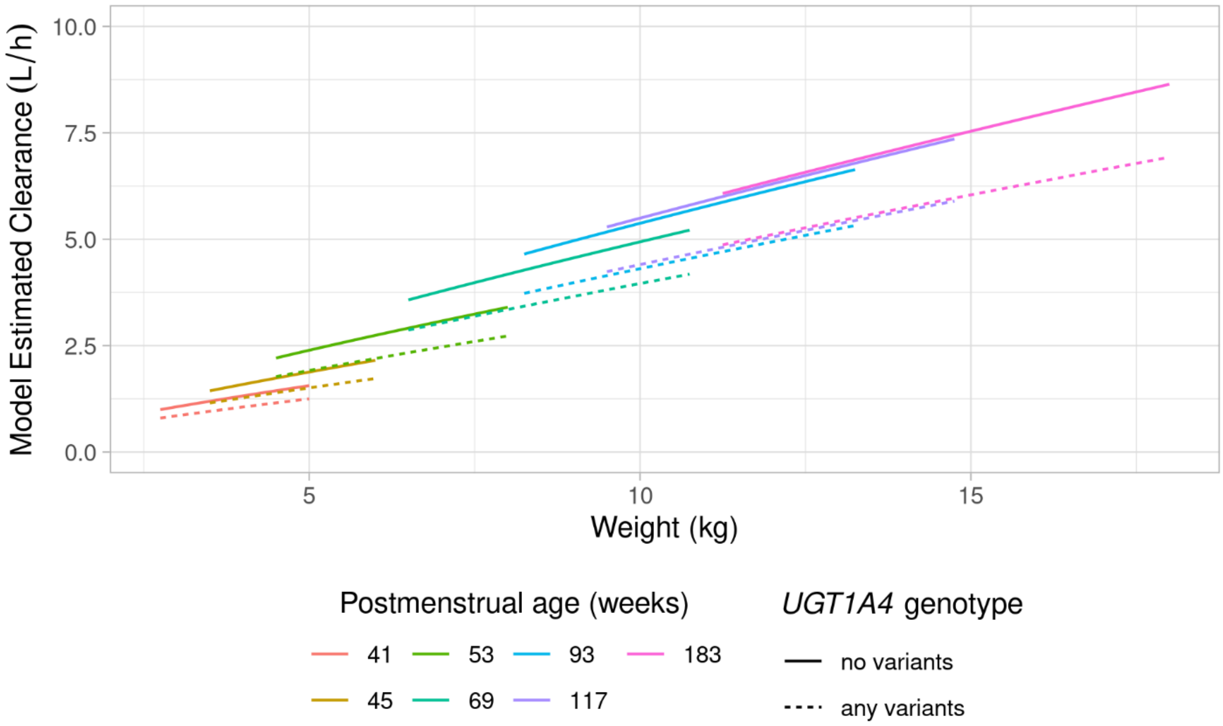


**Figure S30.** Model estimated clearance by *UGT2B10* genotype.

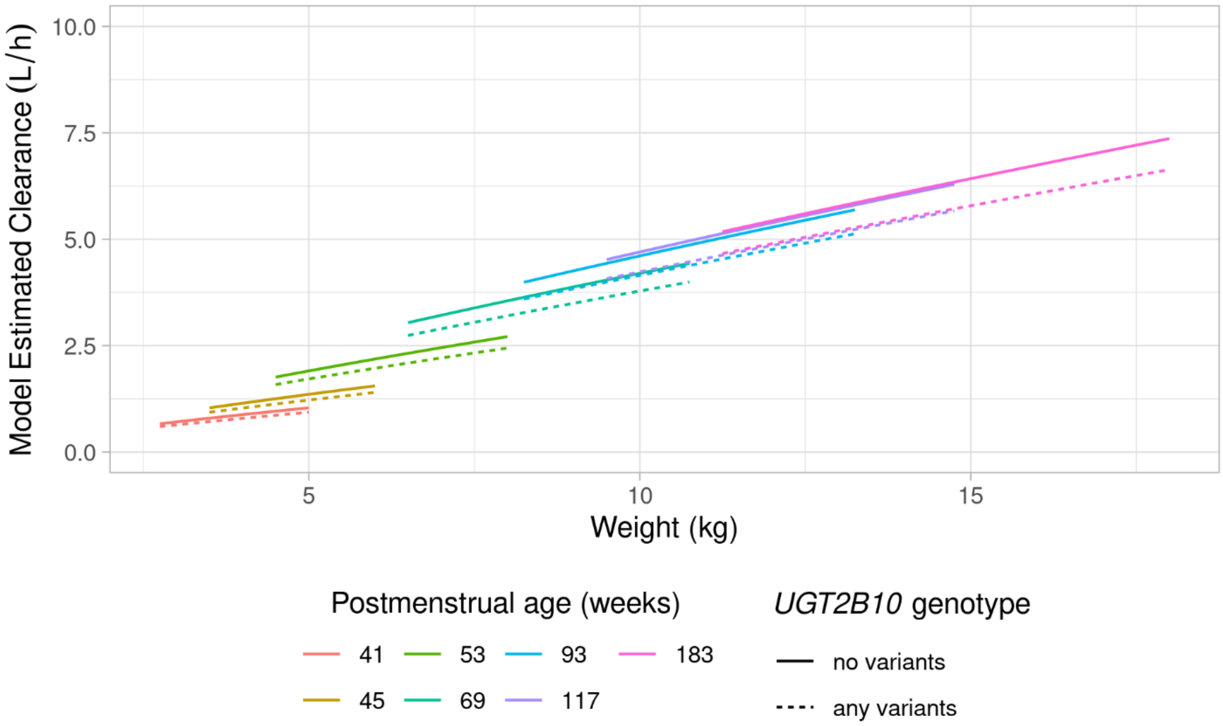

*Simulations of Concentration*

To demonstrate the hypothetical effect of genotype covariates on predicted dexmedetomidine concentration, we estimated predicted concentration-time profiles using PK parameter estimates from the *UGT1A4* and *UGT2B10* categorical models. 90% confidence bounds were produced using quantiles from 200 simulated subjects with residual error (proportional component standard deviation = 0.478 ng/mL) for the following combination of covariates and dosing groups: (1) the $5^{th}$, median, or $95^{th}$ percentile of weight and postmenstrual age; (2) with or without *UGT1A4* or *UGT2B10* genotype variants; (3) fixed infusion rates of 0.4 or 0.6 mcg/kg/h for a 12-hour infusion. Because we are primarily interested in differences in concentration for the 'same subject' with and without variants, inter-individual variability was not included.

Results are shown in **Figures S31** and **S32**. As expected, subjects with *UGT1A4* variants have higher predicted concentration than those without variants, however the difference between genotypes is much smaller than the residual variability within subjects represented by the colored regions. For the 0.4 mcg/kg/h dosing rate, the predicted concentration levels are below the target range of 0.4 to 0.8 ng/mL for subjects at the $5^{th}$ and median of weight and age both with and without variants. In contrast, the 0.6 mcg/kg/h rate yields predicted concentration levels within the target range for both genotype groups. Similar patterns are also seen for simulated concentrations from the *UGT2B10* model.

**Figure S31.** Predicted concentration simulations from *UGT1A4* categorical model.

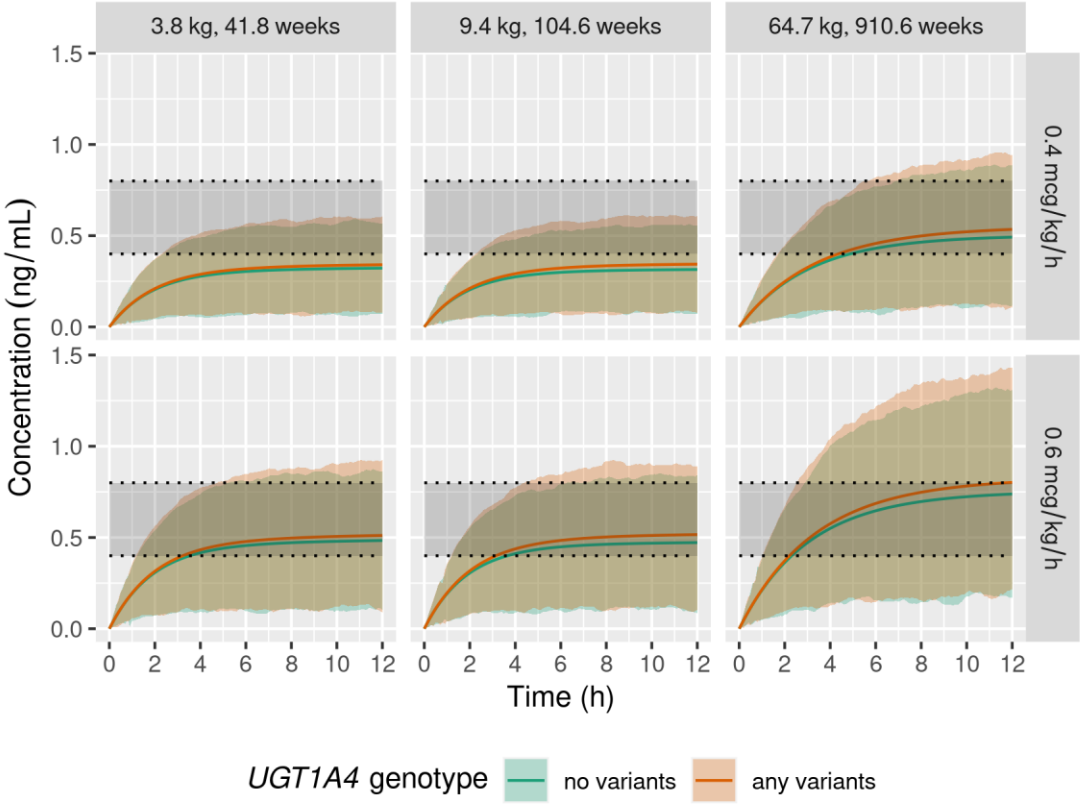

**Figure S32.** Predicted concentration simulations from *UGT2B10* categorical model.

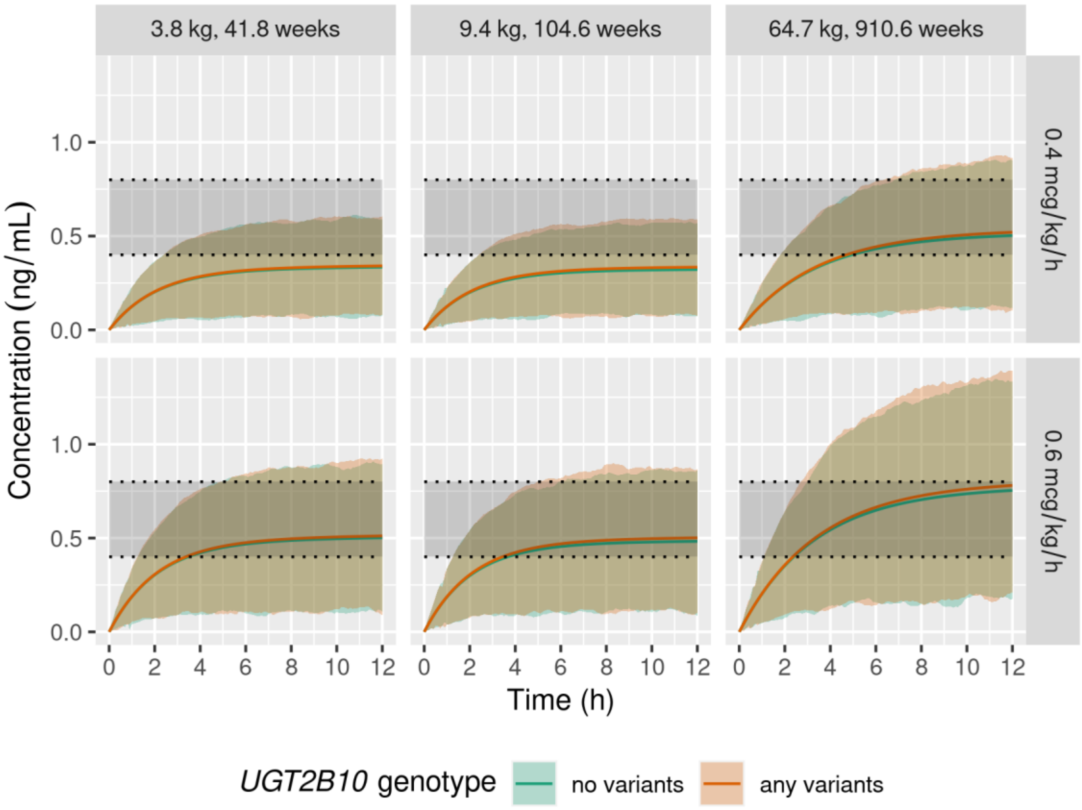

We also performed simulations to find the infusion rates that would yield similar concentrations for subjects of the same age and weight with and without genetic variants after a 12-hour infusion. First, concentration-time profiles were generated using identical dosing rate (0.6 mcg/kg/h), weight (50kg), and postmenstrual age (520 weeks) for a simulated subject with and without *UGT1A4* or *UGT2B10* variants, respectively. Then the infusion rate for the simulated subject with variants was adjusted in increments of 0.01 mcg/kg/h until the concentration achieved at the end of the 12-hour infusion most closely matched the concentration of the simulated subject without variants. The R package mrgsolve was used for all simulations. The simulated patient with *UGT1A4* variants required a rate of 0.56 mcg/kg/h to approximate the concentration of those without *UGT1A4* variants at a rate of 0.6 mcg/kg/h. The simulated patient with *UGT2B10* variants required a rate of 0.58 mcg/kg/h to approximate the concentration of those without *UGT1A4* variants at a rate of 0.6 mcg/kg/h. These changes in dose are not large enough to impact clinical dosing as increments for dexmedetomidine titration are typically 0.1 mcg/kg/h.

**Figure S33.** Simulated Dose needed to achieve same concentration for patients of same postmenstrual age (520 weeks) and weight (50 kg) with and without variants from *UGT1A4* model.

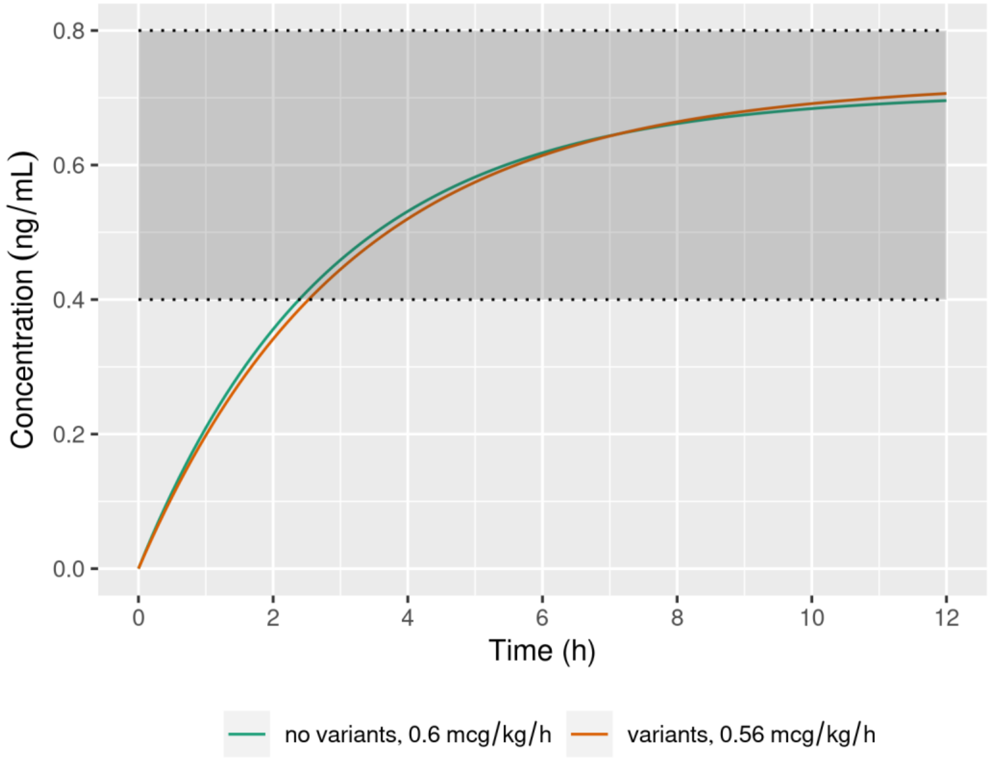

**Figure S34.** Simulated Dose needed to achieve same concentration for patients of same postmenstrual age (520 weeks) and weight (50 kg) with and without variants from *UGT2B10* model.

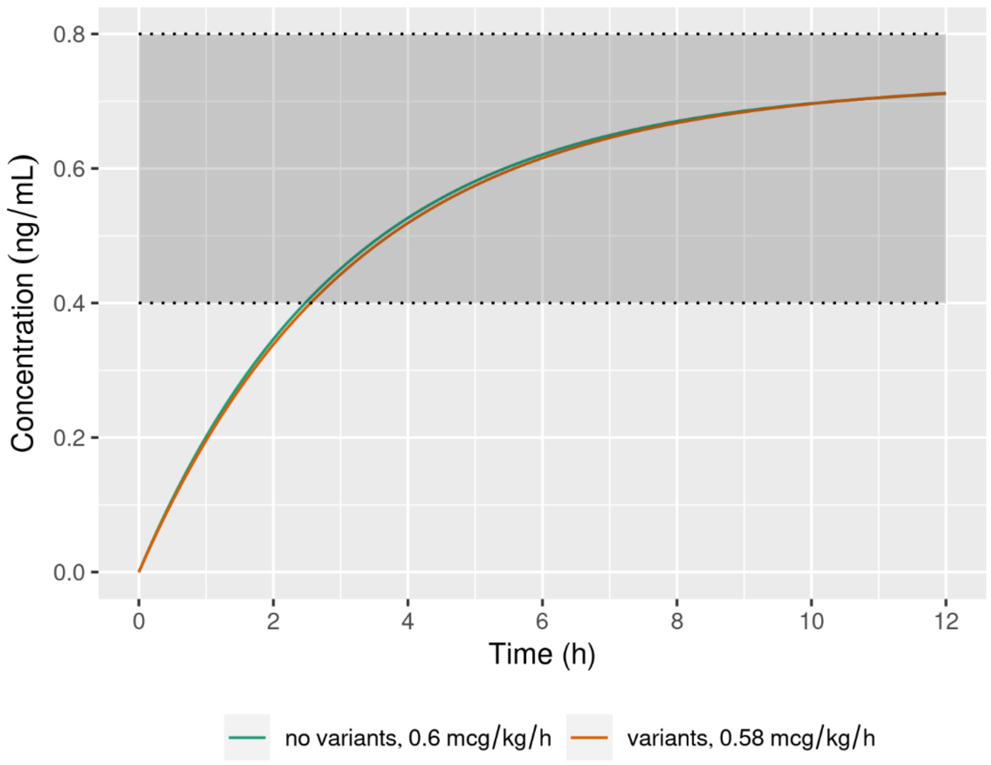
